# The Genetic Determinants of Aortic Distension

**DOI:** 10.1101/2021.10.16.21265089

**Authors:** James P. Pirruccello, Seung Hoan Choi, Mark D. Chaffin, Mahan Nekoui, Elizabeth L. Chou, Sean J. Jurgens, Samuel F. Friedman, Dejan Juric, James R. Stone, Puneet Batra, Kenney Ng, Anthony A. Philippakis, Mark E. Lindsay, Patrick T. Ellinor

## Abstract

As the largest conduit vessel, the aorta is responsible for the conversion of phasic systolic inflow from ventricular ejection into more continuous blood delivery to peripheral arteries. Distension during systole and recoil during diastole conserves ventricular energy and is enabled by the specialized composition of the aortic extracellular matrix. Aortic distensibility decreases with age and prematurely in vascular disease. To discover genetic determinants of aortic distensibility we trained a deep learning model to quantify aortic size throughout the cardiac cycle and calculate aortic distensibility and aortic strain in 42,342 participants in the UK Biobank with available cardiac magnetic resonance imaging. In up to 40,028 participants with genetic data, common variant analysis identified 12 and 26 loci for ascending and 11 and 21 loci for descending aortic distensibility and strain, respectively. Of the newly identified loci, 22 were specific to strain or distensibility and were not identified in a thoracic aortic diameter GWAS within the same samples. Loci associated with both aortic diameter and aortic strain or distensibility demonstrated a consistent, inverse directionality. Transcriptome-wide analyses, rare-variant burden tests, and analyses of gene expression in single nucleus RNA sequencing of human aorta were performed to prioritize genes at individual loci. Loci highlighted multiple genes involved in elastogenesis, matrix degradation, and extracellular polysaccharide generation. Characterization of the genetic determinants of aortic function may provide novel targets for medical intervention in aortic disease.

## Introduction

Pathogenic insult to the thoracic aorta comes from different influences including hypertension, hypercholesterolemia, aging, valvular dysfunction, and genetic perturbations that interfere with aortic homeostasis. Aortic disease may manifest under these influences variably as early or advanced atherosclerotic lesions, intramural hematomas, aneurysms, or even aortic failure in the form of rupture or dissection. One common early manifestation resulting from all of these predisposing factors to aortic disease is a reduction in aortic motion, or “stiffening”, that can be detected via imaging or hemodynamic consequences.

Every cardiac cycle the aorta accepts the output of ventricular systole to transport blood to target tissues, hence its designation as a “conduit vessel”. However the aorta is not a simple conduit but part of an hemodynamic system that conserves ventricular energy and ensures flow in the distal vasculature during diastole. This “windkessel” function of the aorta occurs when aortic strain stores potential energy during systolic ejection which is then utilized during diastole to maintain forward flow. This aortic property can be estimated noninvasively by measurement of aortic distensibility, which indexes aortic strain (the measured relative change in cross-sectional area from diastole to systole) to a non-invasive estimate of central pulse pressure. In addition to impairing distal blood delivery, stiffening of the aorta also has deleterious effects on the heart by increasing ventricular afterload and decreasing the coronary flow reserve ratio thereby impairing endocardial blood flow^1^. Decreased aortic distensibility is seen with normal aging^2^, but also prematurely in multiple cardiovascular diseases including hypertension^3^, atherosclerosis^4^, aneurysmal disease^5–7^, aortic dissection^8^, cognitive impairment and dementia^9^, and is predictive of non-aortic cardiovascular events and all-cause mortality^10^. Despite the importance of aortic distensibility in health and disease there is a limited understanding of its basic biologic mechanisms and their relationship to comorbidities in humans.

In this study we quantify aortic distention from 42,342 participants in the UK Biobank to discover epidemiologic associations with anthropomorphic phenotypes, relationship to other medical disorders, and the underlying human genetic architecture of this fundamental aortic function.

## Results

### Semantic segmentation of aorta with deep learning

A deep learning model based on ResNet34 was trained to annotate the ascending and descending aorta from 501 manually annotated training examples^11,12^. This model had high accuracy: in a held-out test set of 20 images not used for training, the average Dice score (which has a maximum value of 1 for perfect agreement, see **Methods**) was 0.97 for the ascending aorta and 0.96 for the descending aorta (**Supplementary Table 1**). The model was applied to all available aortic distensibility images in 42,342 UK Biobank participants. This allowed measurement of aortic diameter and circumferential aortic strain, which is a dimensionless representation of how much the aortic cross-sectional area changes between its minimum and maximum value during the cardiac cycle (**Figure 1** **Panels A** and **B**). Strain was then combined with a previously described estimate of central pulse pressure to produce estimates of aortic distensibility^13,14^.

**Figure 1.**
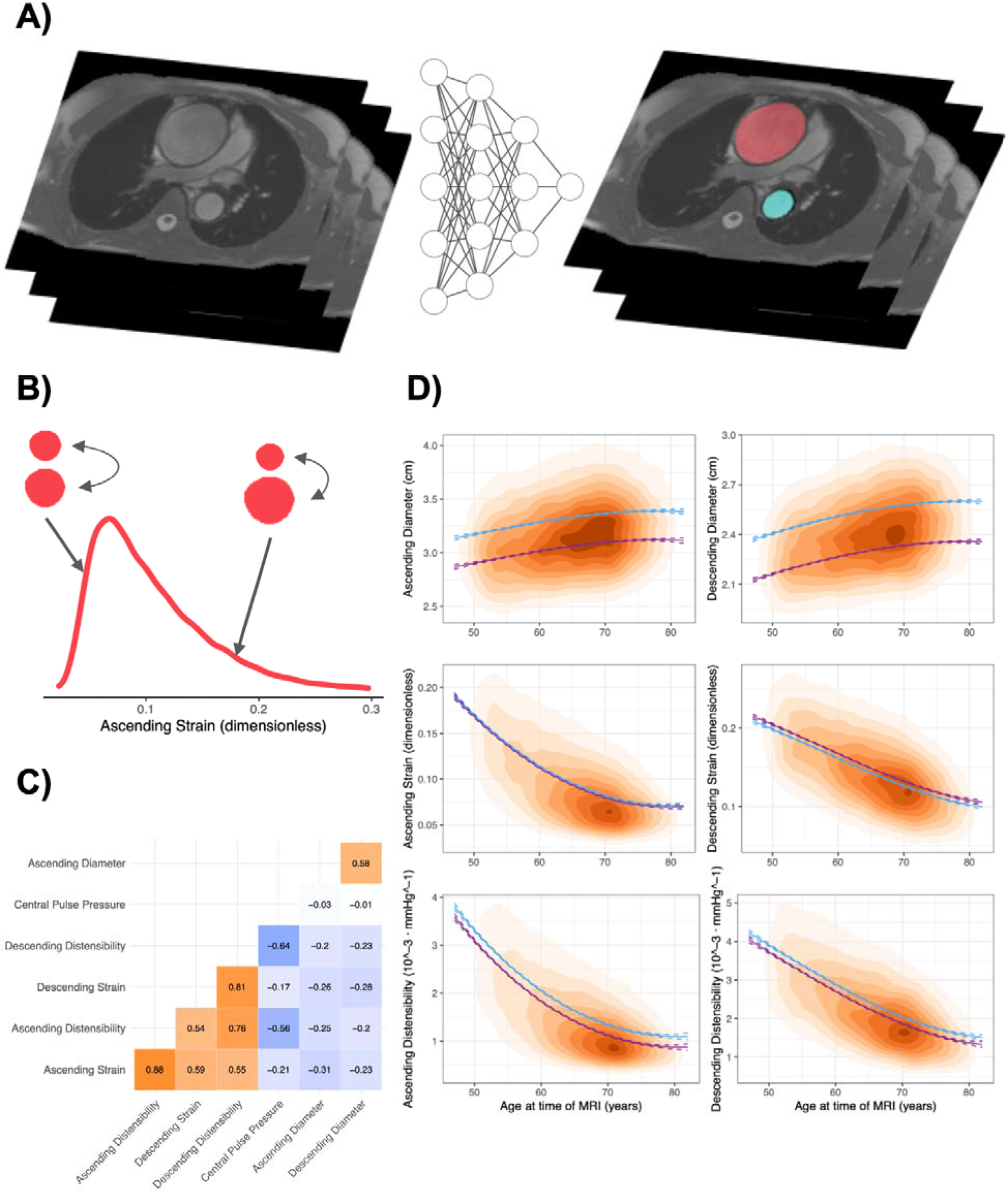
Phenotype derivation and distribution. **Panel A**: Depiction of the derivation of ascending (red) and descending (turquoise) aortic diameter with a deep learning model. **Panel B**: Population distribution of ascending aortic strain, with exaggerated maximum and aortic cross-sections of participants with typical strain (left) and above average strain (right) for illustration purposes. **Panel C:** Cross-correlation between aortic phenotypes and central pulse pressure. **Panel D**: Population density based on age at time of MRI and aortic phenotypic readings. Plots are shaded with increasing color intensity by population density. Modeled mean values by age and sex were estimated from a linear model that included sex and a 3 degree-of-freedom basis spline for age. These estimates are plotted in blue for men and in purple for women.

### Epidemiological characteristics of aortic strain and distensibility in the UK Biobank

For the 40,577 participants who contributed to at least one GWAS (**Supplementary Figure 1**), the distribution of ascending and descending aortic strain and distensibility are shown in **Supplementary Figure 2**. Cross-correlations of traits including diameter, distensibility, strain, and central pulse pressure are shown in **Figure 1** **Panel C**. All data are described after an initial quality control step, described in more detail in the **Supplementary Note**, which resolved a profound right skew of the distensibility estimates due to likely spurious central pulse pressure estimates. Consistent with previous reports, aortic distensibility was inversely associated with age (**Figure 1** **Panel D**; **Supplementary Table 2**)^2,15^. Distensibility and strain were also associated with continuous cardiovascular measurements such as heart rate, blood pressure, physical activity, and fat mass (**Supplementary Figure 3**, **Supplementary Table 3**).

We also assessed the relationships between aortic phenotypes and several cardiovascular diseases (defined in **Supplementary Table 4**) and their risk factors. In linear models adjusted for age, sex, and the first five principal components of ancestry, hypertension diagnosed prior to imaging was associated with lower strain and distensibility (**Figure 2** **left panel**). The presence of coronary artery disease (CAD) was associated with increased strain and distensibility, contrary to expectation^16^. The strongest CAD association was with descending aortic strain, which was 0.24 standard deviation (SD) higher in those with a history of CAD (P = 1.6E-26). Previous reports have described that beta blocker administration can increase aortic distensibility^6^. To investigate whether a medication effect might mediate the unexpected CAD-strain observation, we computed the association between aortic phenotypes and medication classes being taken by study participants as defined by Anatomical Therapeutic Chemical (ATC) codes^17^ (**Supplementary Figure 4**). The 2,265 participants who reported taking beta blockers had strain and distensibility that were as much as 0.25 standard deviation higher than those of participants not reporting use of beta blockers (P = 2.2E-40; **Supplementary Table 5**). After excluding participants taking beta blockers (leaving 842 of the original 1,550 participants with prevalent CAD), the association effect size of the presence of CAD on descending strain diminished from 0.24 to 0.13, and the association P value diminished by 20 orders of magnitude, suggesting that pharmacological therapies may mediate a significant degree of this otherwise counterintuitive observation.

**Figure 2.**
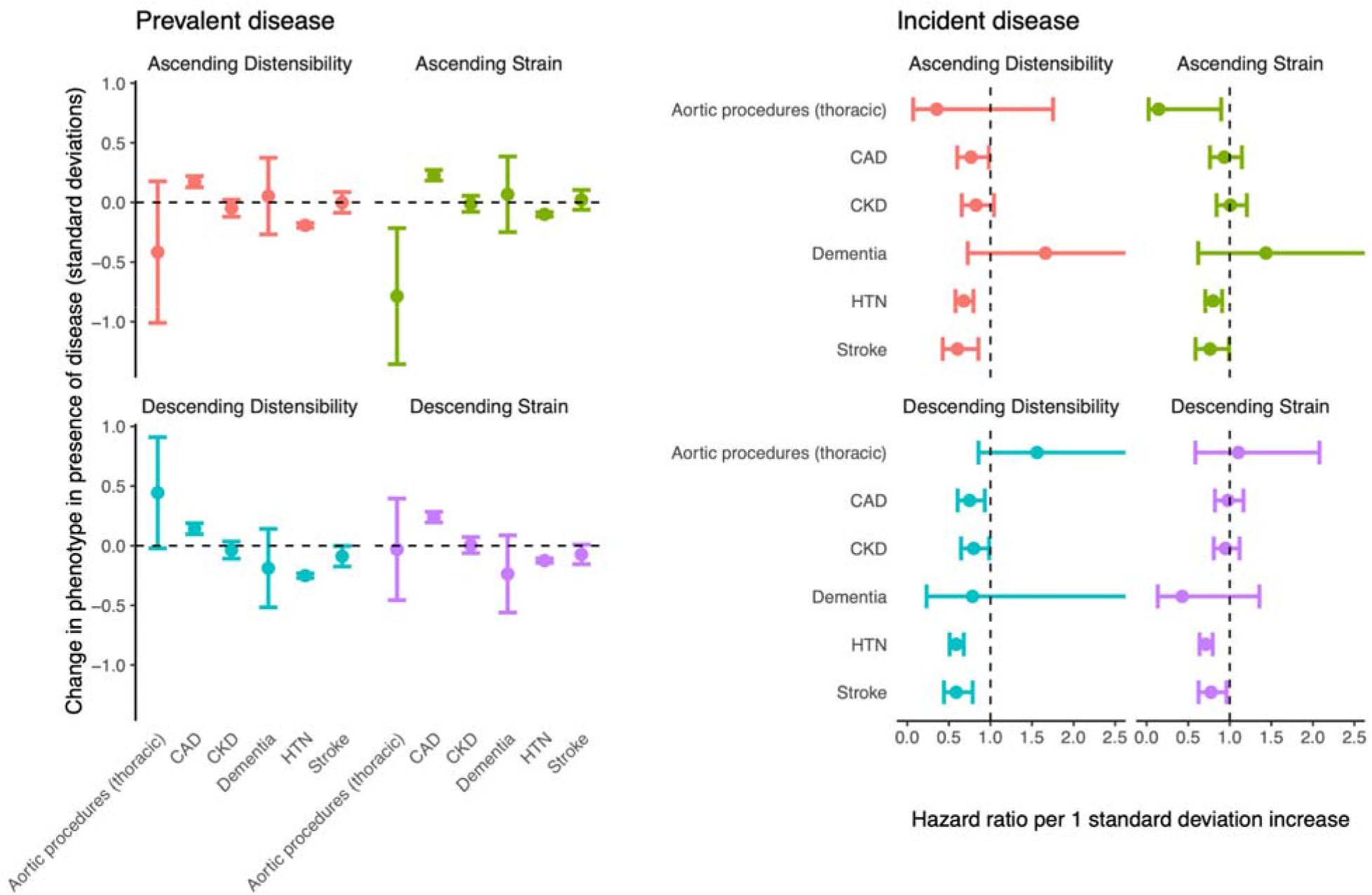
Relationship between strain, distensibility, and disease. **Left panel** (“Prevalent disease”): the difference in aortic phenotypes (**y axis**) between UK Biobank participants with a history of thoracic aortic procedures, CAD, chronic kidney disease (“CKD”), Alzheimer’s disease or dementia (“Dementia”), hypertension (“HTN”), or stroke (“Stroke”) occurring prior to MRI, compared to participants without a history of disease (**x axis**). **Right panel** (“Incident disease”): hazard ratios for disease incidence (**y axis**) occurring after MRI per one standard deviation increase in the aortic phenotypes (**x axis**). Point estimates are represented by a dot; 95% confidence intervals for the effect estimate are represented by error bars.

To expand the analysis of potential pharmacological effects, we produced estimates of the effect of medications on aortic phenotypes: 2,712 participants had undergone two UK Biobank imaging visits; a small number of participants had started taking one or more new medications in between the two visits, allowing an estimate of 43 medication effects on aortic phenotypes at two-tailed P < 0.05 (**Supplementary Table 6**). Although the data did not permit a causal analysis, we conducted PheCode-based association studies before (**Supplementary Table 7**; **Supplementary Figure 5**) and after (**Supplementary Table 8**; **Supplementary Figure 6**) adjusting the aortic phenotypes to account for the additive effect size of medications (see **Methods**)^18^. Accounting for medications eliminated much of the apparent relationship between ischemic heart disease and aortic strain or distensibility.

Finally, we examined incident cardiovascular diseases that occurred after the aortic phenotypes had been measured. We found greater aortic strain to be associated with significantly lower chances of being subsequently diagnosed with hypertension (547 incident cases; HR 0.59 per SD increase in descending aortic strain; P = 3.8E-13) or stroke (146 incident cases; HR 0.59 per SD increase in descending aortic strain; P = 3.1E-04) (**Supplementary Table 9**; **Figure 2** **right panel**).

### Heritability of aortic distensibility

Of the 42,342 participants with aortic measurements, 40,577 had genetic data and contributed to at least one genetic analysis (**Supplementary Figure 1**; **Supplementary Table 10**). The SNP-heritability of aortic distensibility was moderate: heritability estimates using BOLT-REML were of 25% and 22% for ascending and descending aortic distensibility, respectively^19,20^. Aortic strain had a higher SNP-heritability: 33% for the ascending aorta and 30% for the descending aorta by BOLT-REML (**Supplementary Table 11**; cross-trait genetic correlations in **Supplementary Table 12** and **Supplementary Figure 7**).

### Genome wide association study of aortic strain and distensibility

We then conducted a GWAS for each aortic trait with BOLT-LMM^19,20^. At a commonly used significance threshold of 5E-08, we identified 26 loci associated with ascending aortic strain, 21 with descending aortic strain, 12 with ascending aortic distensibility, and 11 with descending aortic distensibility (**Figure 3**; QQ plots in **Supplementary Figure 8**; *ldsc* intercepts in **Supplementary Table 13**). All distensibility-associated loci are shown in **Table 1**, the full list of strain and distensibility loci is in **Supplementary Table 14**, and locus plots are in **Supplementary Figure 9**.

**Figure 3.**
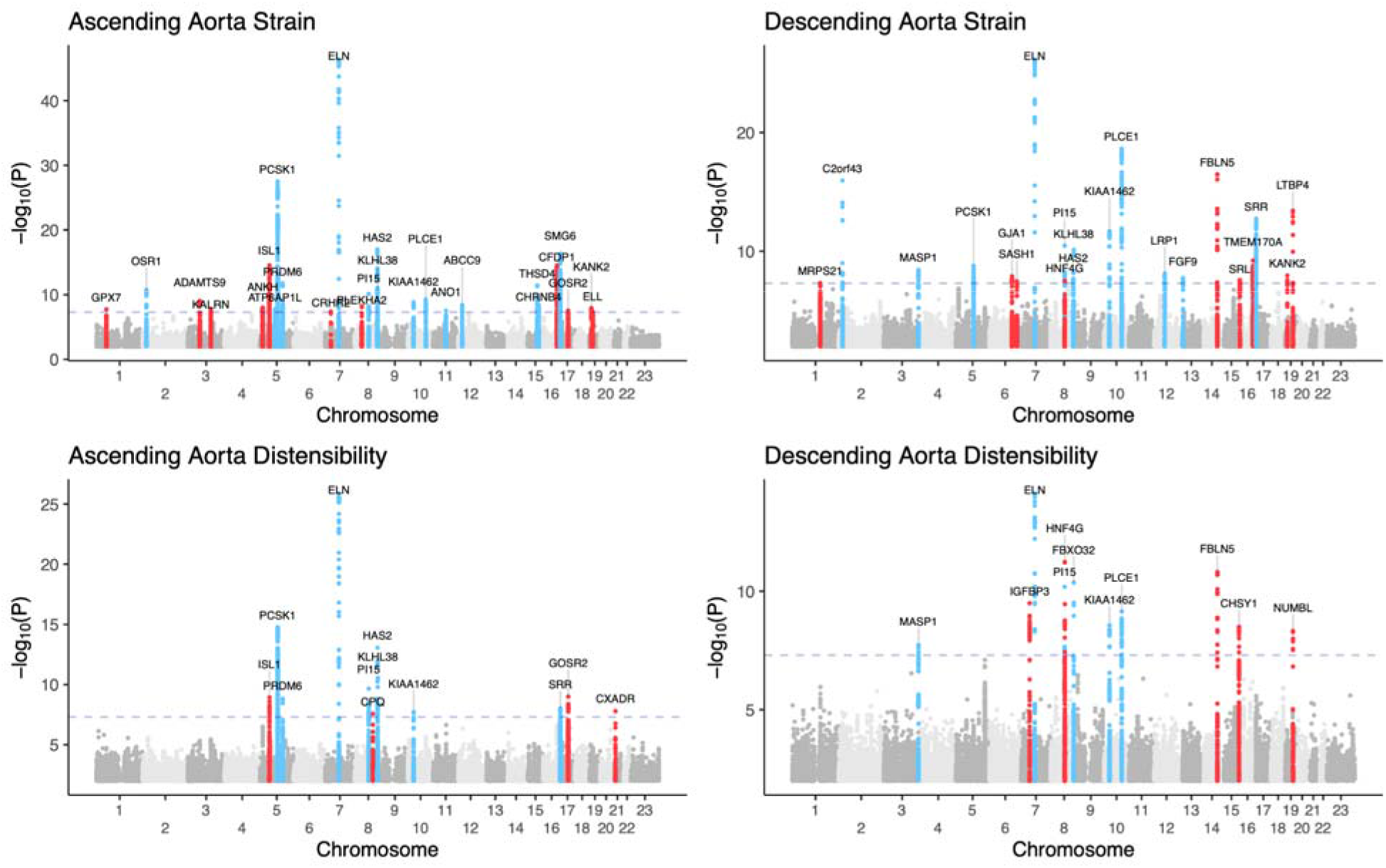
Manhattan plots for genome wide association studies of aortic strain and distensibility. Manhattan plots depicting ascending and descending aortic strain and distensibility. Chromosomal position is on the **x axis**, and -log10(P value) is on the **y axis**. Loci with at least one SNP with P < 5E-08 are depicted in color and labeled with the name of the nearest gene. Loci that were not associated with aortic diameter are highlighted in red.

**Table 1.**
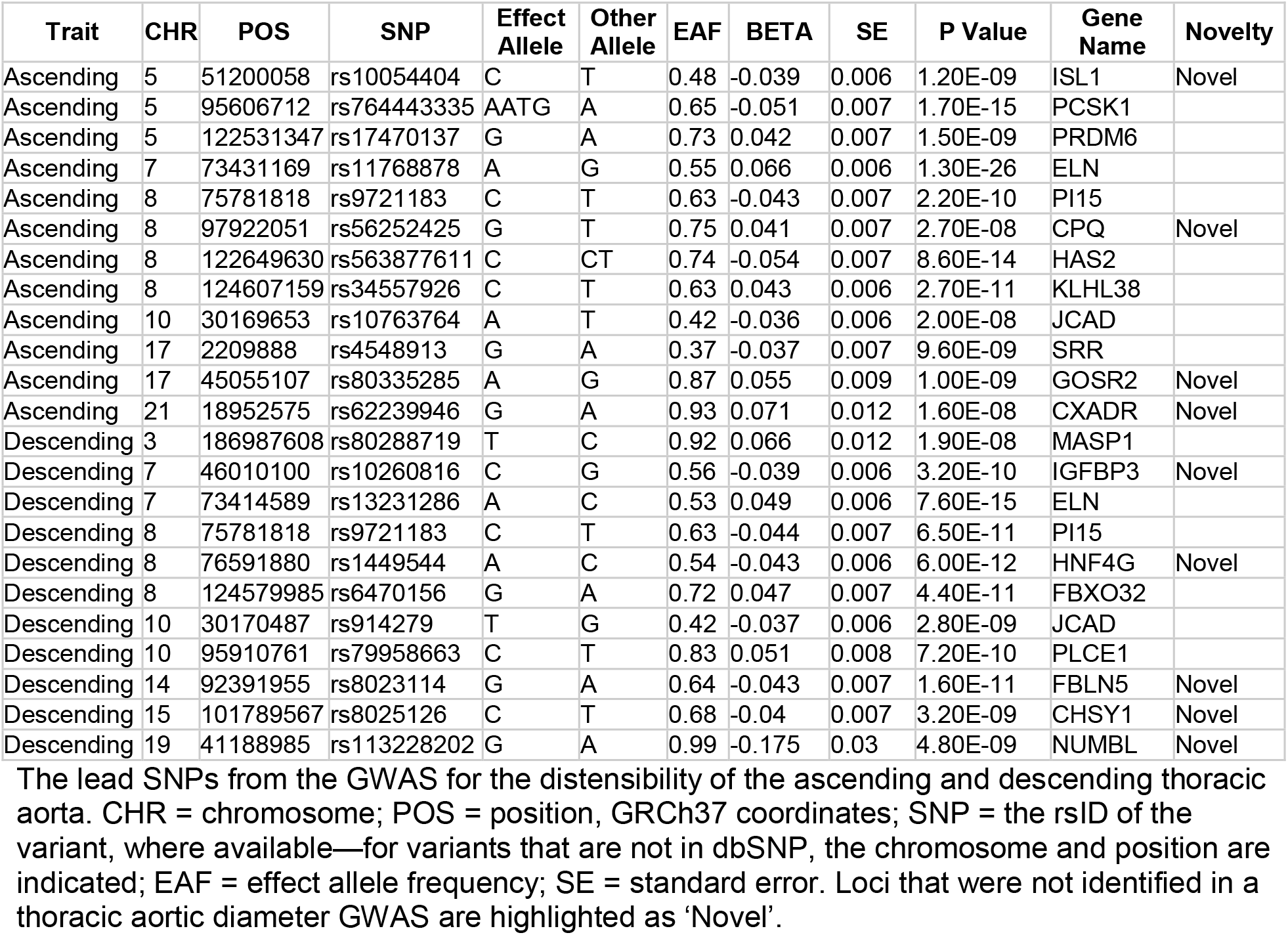
GWAS loci for distensibility of the ascending and descending aorta

| Trait | CHR | POS | SNP | Effect Allele | Other Allele | EAF | BETA | SE | P Value | Gene Name | Novelty |
| --- | --- | --- | --- | --- | --- | --- | --- | --- | --- | --- | --- |
| Ascending | 5 | 51200058 | rs10054404 | C | T | 0.48 | -0.039 | 0.006 | 1.20E-09 | ISL1 | Novel |
| Ascending | 5 | 95606712 | rs764443335 | AATG | A | 0.65 | -0.051 | 0.007 | 1.70E-15 | PCSK1 |  |
| Ascending | 5 | 122531347 | rs17470137 | G | A | 0.73 | 0.042 | 0.007 | 1.50E-09 | PRDM6 |  |
| Ascending | 7 | 73431169 | rs11768878 | A | G | 0.55 | 0.066 | 0.006 | 1.30E-26 | ELN |  |
| Ascending | 8 | 75781818 | rs9721183 | C | T | 0.63 | -0.043 | 0.007 | 2.20E-10 | PI15 |  |
| Ascending | 8 | 97922051 | rs56252425 | G | T | 0.75 | 0.041 | 0.007 | 2.70E-08 | CPQ | Novel |
| Ascending | 8 | 122649630 | rs563877611 | C | CT | 0.74 | -0.054 | 0.007 | 8.60E-14 | HAS2 |  |
| Ascending | 8 | 124607159 | rs34557926 | C | T | 0.63 | 0.043 | 0.006 | 2.70E-11 | KLHL38 |  |
| Ascending | 10 | 30169653 | rs10763764 | A | T | 0.42 | -0.036 | 0.006 | 2.00E-08 | JCAD |  |
| Ascending | 17 | 2209888 | rs4548913 | G | A | 0.37 | -0.037 | 0.007 | 9.60E-09 | SRR |  |
| Ascending | 17 | 45055107 | rs80335285 | A | G | 0.87 | 0.055 | 0.009 | 1.00E-09 | GOSR2 | Novel |
| Ascending | 21 | 18952575 | rs62239946 | G | A | 0.93 | 0.071 | 0.012 | 1.60E-08 | CXADR | Novel |
| Descending | 3 | 186987608 | rs80288719 | T | C | 0.92 | 0.066 | 0.012 | 1.90E-08 | MASP1 |  |
| Descending | 7 | 46010100 | rs10260816 | C | G | 0.56 | -0.039 | 0.006 | 3.20E-10 | IGFBP3 | Novel |
| Descending | 7 | 73414589 | rs13231286 | A | C | 0.53 | 0.049 | 0.006 | 7.60E-15 | ELN |  |
| Descending | 8 | 75781818 | rs9721183 | C | T | 0.63 | -0.044 | 0.007 | 6.50E-11 | PI15 |  |
| Descending | 8 | 76591880 | rs1449544 | A | C | 0.54 | -0.043 | 0.006 | 6.00E-12 | HNF4G | Novel |
| Descending | 8 | 124579985 | rs6470156 | G | A | 0.72 | 0.047 | 0.007 | 4.40E-11 | FBXO32 |  |
| Descending | 10 | 30170487 | rs914279 | T | G | 0.42 | -0.037 | 0.006 | 2.80E-09 | JCAD |  |
| Descending | 10 | 95910761 | rs79958663 | C | T | 0.83 | 0.051 | 0.008 | 7.20E-10 | PLCE1 |  |
| Descending | 14 | 92391955 | rs8023114 | G | A | 0.64 | -0.043 | 0.007 | 1.60E-11 | FBLN5 | Novel |
| Descending | 15 | 101789567 | rs8025126 | C | T | 0.68 | -0.04 | 0.007 | 3.20E-09 | CHSY1 | Novel |
| Descending | 19 | 41188985 | rs113228202 | G | A | 0.99 | -0.175 | 0.03 | 4.80E-09 | NUMBL | Novel |

The lead SNPs from the GWAS for the distensibility of the ascending and descending thoracic aorta. CHR = chromosome; POS = position, GRCh37 coordinates; SNP = the rsID of the variant, where available—for variants that are not in dbSNP, the chromosome and position are indicated; EAF = effect allele frequency; SE = standard error. Loci that were not identified in a thoracic aortic diameter GWAS are highlighted as 'Novel'.

In total, 22 distinct genetic loci were specific to strain and distensibility, after excluding loci that were found among 110 loci that were associated with ascending or descending thoracic aortic diameter within the same sample (**Supplementary Table 15**). These 22 loci included 11 associated with ascending strain, nine with descending strain, four with ascending distensibility, and five with descending distensibility (**Table 2**).

**Table 2.** GWAS loci unique to aortic distensibility and strain

| Trait | CHR | POS | SNP | Effect Allele | Other Allele | EAF | BETA | SE | P Value | Gene Name |
| --- | --- | --- | --- | --- | --- | --- | --- | --- | --- | --- |
| Ascending Aorta Strain | 1 | 53081658 |  | CT | C | 0.44 | 0.036 | 0.006 | 1.80E-08 | GPX7 |
| Ascending Aorta Strain | 3 | 64723072 | rs7638565 | A | G | 0.41 | 0.036 | 0.006 | 9.00E-10 | ADAMTS9 |
| Ascending Aorta Strain | 3 | 124026421 | rs146130720 | A | ATAAGT | 0.35 | -0.037 | 0.006 | 1.60E-08 | KALRN |
| Ascending Aorta Strain | 5 | 15023774 | rs9687138 | C | A | 0.83 | -0.045 | 0.008 | 9.90E-09 | ANKH |
| Ascending Aorta Strain | 5 | 51201361 | rs13158444 | T | C | 0.41 | -0.049 | 0.006 | 3.00E-15 | ISL1 |
| Ascending Aorta Strain | 7 | 30727202 | rs75520340 | T | A | 0.97 | -0.092 | 0.017 | 4.30E-08 | CRHR2 |
| Ascending Aorta Strain | 8 | 38761012 | rs75233095 | G | A | 0.83 | -0.046 | 0.008 | 5.70E-09 | PLEKHA2 |
| Ascending Aorta Strain | 16 | 75413242 | rs150284594 | A | AAG | 0.41 | -0.048 | 0.006 | 3.00E-15 | CFDP1 |
| Ascending Aorta Strain | 17 | 45013271 | rs17608766 | T | C | 0.86 | 0.046 | 0.009 | 2.90E-08 | GOSR2 |
| Ascending Aorta Strain | 19 | 11278681 |  | GT | G | 0.69 | -0.04 | 0.007 | 1.20E-08 | KANK2 |
| Ascending Aorta Strain | 19 | 18567771 | rs796826122 | GA | G | 0.77 | 0.039 | 0.007 | 4.30E-08 | ELL |
| Descending Aorta Strain | 1 | 150267316 | rs143847321 | C | CAAAAAA | 0.49 | -0.032 | 0.006 | 4.80E-08 | MRPS21 |
| Descending Aorta Strain | 6 | 122089704 | rs58730006 | A | AT | 0.9 | 0.056 | 0.01 | 1.30E-08 | GJA1 |
| Descending Aorta Strain | 6 | 148639909 | rs4574664 | G | T | 0.81 | 0.043 | 0.008 | 3.50E-08 | SASH1 |
| Descending Aorta Strain | 8 | 76591880 | rs1449544 | A | C | 0.54 | -0.034 | 0.006 | 3.10E-08 | HNF4G |
| Descending Aorta Strain | 14 | 92393198 | rs8014161 | T | A | 0.64 | -0.051 | 0.006 | 3.30E-17 | FBLN5 |
| Descending Aorta Strain | 16 | 4281391 | rs11864324 | T | G | 0.78 | -0.037 | 0.007 | 2.60E-08 | SRL |
| Descending Aorta Strain | 16 | 75489254 |  | CA | C | 0.49 | 0.038 | 0.006 | 6.00E-10 | TMEM170A |
| Descending Aorta Strain | 19 | 11271714 | rs113783450 | G | A | 0.69 | -0.038 | 0.006 | 1.10E-08 | KANK2 |
| Descending Aorta Strain | 19 | 41117300 | rs34093919 | G | A | 0.99 | -0.203 | 0.027 | 3.90E-14 | LTBP4 |
| Ascending Aorta Distensibility | 5 | 51200058 | rs10054404 | C | T | 0.48 | -0.039 | 0.006 | 1.20E-09 | ISL1 |
| Ascending Aorta Distensibility | 8 | 97922051 | rs56252425 | G | T | 0.75 | 0.041 | 0.007 | 2.70E-08 | CPQ |
| Ascending Aorta Distensibility | 17 | 45055107 | rs80335285 | A | G | 0.87 | 0.055 | 0.009 | 1.00E-09 | GOSR2 |
| Ascending Aorta Distensibility | 21 | 18952575 | rs62239946 | G | A | 0.93 | 0.071 | 0.012 | 1.60E-08 | CXADR |
| Descending Aorta Distensibility | 7 | 46010100 | rs10260816 | C | G | 0.56 | -0.039 | 0.006 | 3.20E-10 | IGFBP3 |
| Descending Aorta Distensibility | 8 | 76591880 | rs1449544 | A | C | 0.54 | -0.043 | 0.006 | 6.00E-12 | HNF4G |
| Descending Aorta Distensibility | 14 | 92391955 | rs8023114 | G | A | 0.64 | -0.043 | 0.007 | 1.60E-11 | FBLN5 |
| Descending Aorta Distensibility | 15 | 101789567 | rs8025126 | C | T | 0.68 | -0.04 | 0.007 | 3.20E-09 | CHSY1 |
| Descending Aorta Distensibility | 19 | 41188985 | rs113228202 | G | A | 0.99 | -0.175 | 0.03 | 4.80E-09 | NUMBL |

The lead SNPs from the GWAS of the strain and distensibility of the ascending and descending thoracic aorta which were not present in GWAS of thoracic aortic diameter. CHR = chromosome; POS = position, GRCh37 coordinates; SNP = the rsID of the variant, where available—for variants that are not in dbSNP, the chromosome and position are indicated; EAF = effect allele frequency; SE = standard error.

For all lead SNPs that were associated with either ascending aortic strain or distensibility, we observed near-perfect correlation between the strain- or distensibility-increasing allele and decreased aortic diameter (**Figure 4**).

**Figure 4.**
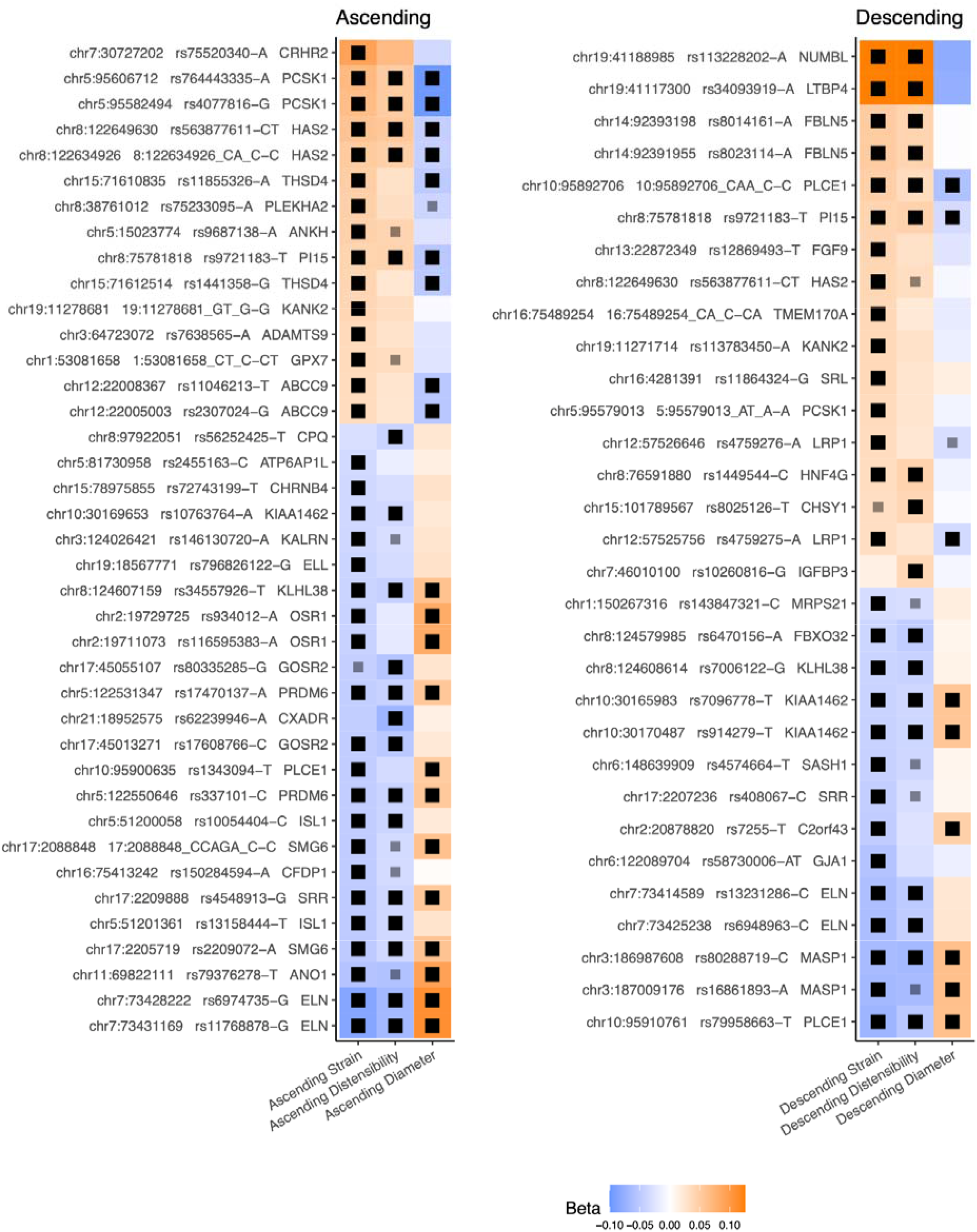
Inverse relation between the genetic determinants of aortic strain or distensibility and aortic diameter. Lead SNPs for ascending (**left panel**) and descending (**right panel**) aortic strain and distensibility are plotted adjacent to the corresponding effect size for aortic diameter. Multiple SNPs at each genomic locus may be displayed when the lead SNP differs across phenotypes. For each SNP, the effects have been aligned such that the minor allele within the study population is modeled. The label indicates the chromosomal position, the SNP and modeled (minor) allele, and the name of the nearest gene transcript. Effect size is represented by color. Black squares indicate P < 5E-08; gray squares indicate P < 5E-06. Effect size range is clamped between -0.1 and 0.125.

### Gene prioritization with MAGMA and TWAS

To prioritize genes for each aortic phenotype, we applied multi-marker analysis of genomic annotation (“MAGMA”)^21^. The gene-based MAGMA analyses of the strain and distensibility GWAS results prioritized fewer than 10 genes in each distensibility GWAS, and approximately twice as many genes in each strain GWAS (**Figure 5**). The most strongly prioritized genes included *SMG6*, *PLCE1*, *PCSK1*, and *PI15*.

**Figure 5.**
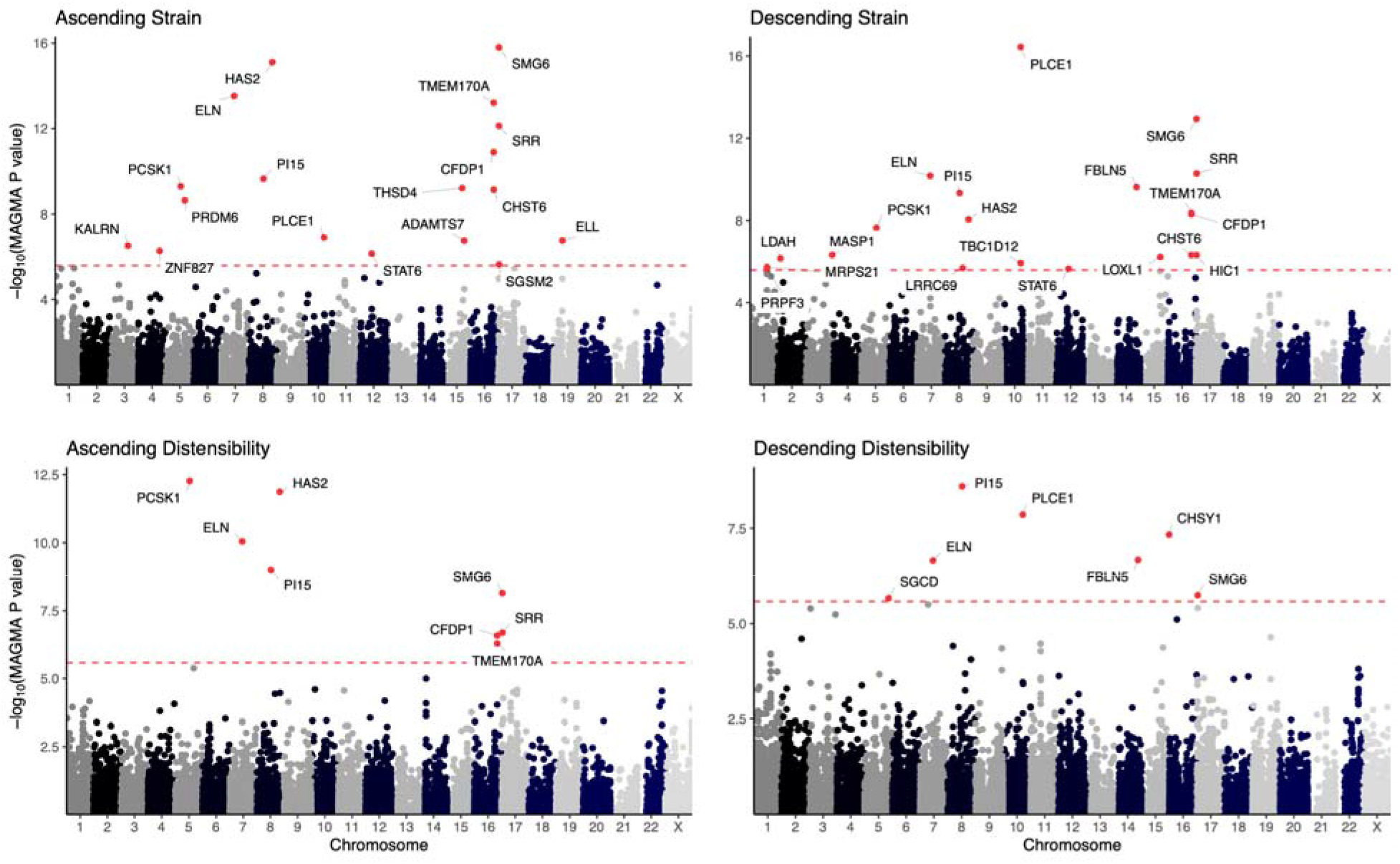
MAGMA-based gene prioritization of aortic strain and distensibility. For each trait, the genes’ chromosomal positions are on the **x axis** and -log10 of the MAGMA P value are on the **y axis**. The horizontal dashed line indicates the per-trait Bonferroni significance.

An alternative approach for gene prioritization is transcriptome wide association study (TWAS), linking predicted gene expression in aorta (based on GTEx v8) with aortic strain and distensibility (**Figure 6**)^22–25^. We identified 23 transcripts that were significantly associated with strain or distensibility at a Bonferroni P < 1.7E-06 (**Supplementary Table 16**). The strongest evidence for association was found between ascending aortic strain and genetically predicted *BCAR1* expression, which has previously been linked to carotid intima-media thickness^26^.

**Figure 6.**
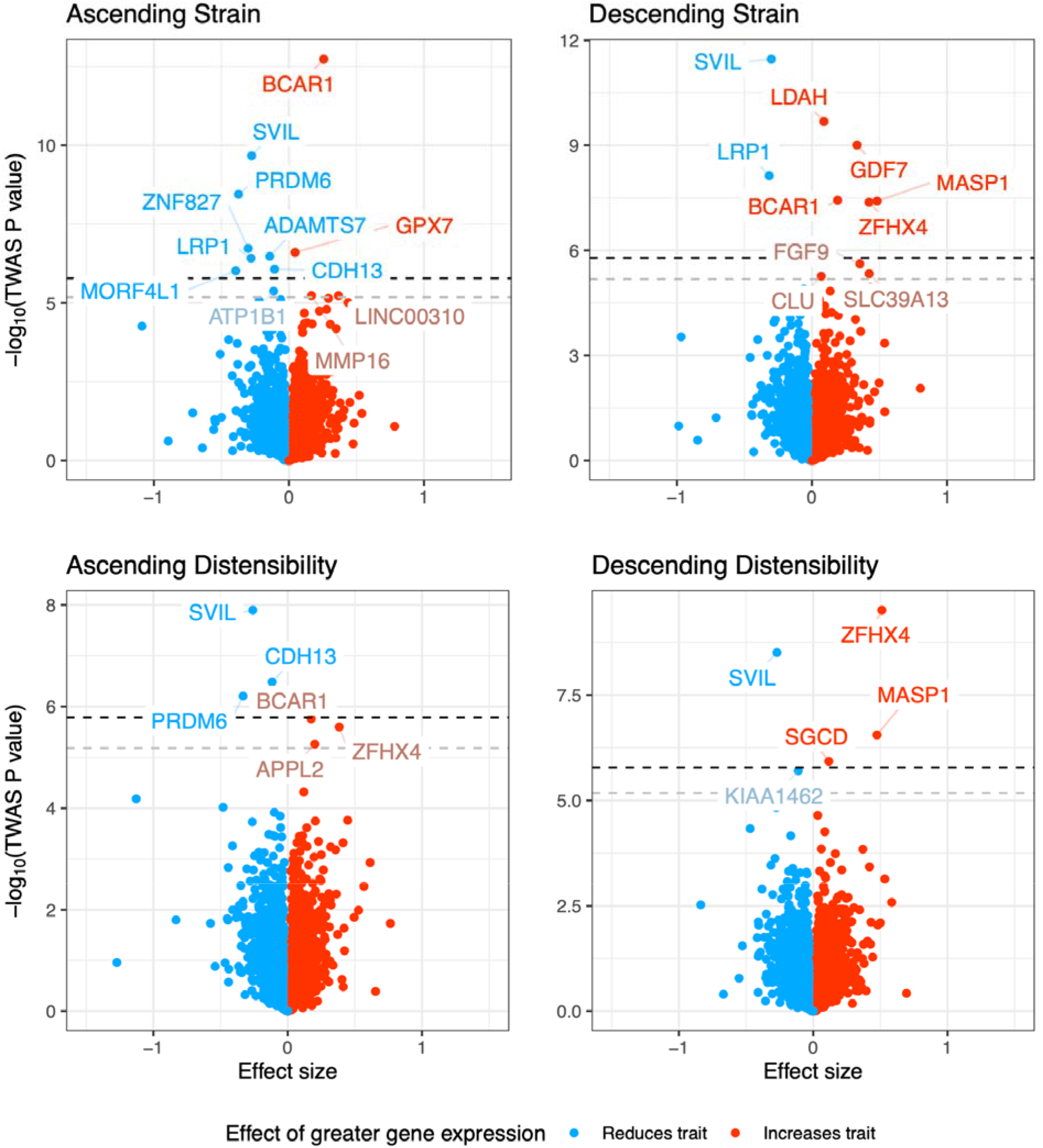
Transcriptome wide analyses of aortic strain and distensibility. TWAS results for ascending and descending aortic distensibility and strain. The **x axis** represents the TWAS effect size, whose units are the predicted standard deviation change in the aortic phenotype per unit change in predicted gene expression; outliers beyond ±1.5 units have been elided. The **y axis** represents the -log_10_ of the TWAS P value. The dark horizontal dashed line represents study-wide Bonferroni significance (adjusted for the total number of tests across all four phenotypes). The light horizontal dashed line represents Bonferroni significance accounting only for the tests performed for that trait. Genes with study-wide significance are labeled with blue (trait-decreasing) or red (trait-increasing) text. Genes with trait-wide significance are labeled with muted blues and reds.

Using single-nucleus RNA sequencing (snRNA-seq) data from the ascending and descending aorta from 3 individuals, we examined the expression of candidate genes linked with aortic distensibility through our TWAS or GWAS analyses^27^. Cell type-specific expression was analyzed for genes that were prioritized by the TWAS or due to being the nearest gene to a GWAS locus (**Supplementary Figure 10**). On average, the greatest expression of these prioritized genes was found in the vascular smooth muscle cells.

### Rare variant association testing

In a rare variant analysis of 22,487 participants with both aortic measurements and exome sequencing data, loss-of-function (LoF) variants in one gene, *PLCE1*, achieved exome-wide significance for association with descending aortic diameter. 23 participants had LoF variants in *PLCE1*, with an estimated effect size of -0.81 standard deviation in distensibility for the LoF carriers (P = 4.2E-06; **Supplementary Figure 11**).

### Polygenic score analyses

We constructed ∼1.1 million SNP polygenic scores using PRScs-auto, and surveyed the connection between those scores and cardiovascular diseases in UK Biobank participants using accelerated failure time models^28,29^. In analyses adjusted for age, sex, and genetic principal components, the polygenic scores were associated with hypertension, CAD, chronic kidney disease (CKD), stroke, and thoracic aortic surgical procedures (**Supplementary Figure 12**; **Supplementary Table 17**). The largest effect size was found between the ascending aortic diameter polygenic score and risk for thoracic aortic procedures: a 1 standard deviation (SD) increase in the score was associated with a 32.5% acceleration in disease onset, P = 8.4E-12. Otherwise, the effect sizes were small: equivalent to delaying or accelerating disease onset by 3-5% per standard deviation change in polygenic score. After adjustment for the aortic diameter polygenic scores, the polygenic prediction of descending aortic distensibility remained statistically significantly associated with risk for hypertension, CAD, CKD, and stroke, with small effect sizes (**Supplementary Figure 18**).

## Discussion

The primary function of large conduit arteries such as the aorta is to convey blood flow through the body. However, beyond this conduit function, the aorta decreases cardiac work through the storage of energy in systole as aortic strain and recovery of this energy in diastole with a loss of only 15-20%^30^. The extracellular matrix of the aorta is responsible for providing the reversible elasticity necessary for this function. Aortic distensibility ensures optimal coronary artery perfusion and buffers pulsatility delivered to distal capillary beds, associated with endothelial and end-organ dysfunction^31^. Multiple cardiovascular morbidities have been associated with abnormalities of aortic distensibility, most prominently atherosclerosis, aneurysms, hypertension, and aging. Despite the importance of this function in the cardiovascular system, few biologic determinants of aortic strain and distensibility in humans have been identified to date. We and others have previously analyzed the thoracic aortic dimension at large scale in the UK Biobank^15,27,32^. In this study, we extended this work by measuring thoracic aortic strain and distensibility, then analyzing their epidemiological properties and genetic architecture.

The deep learning model that we developed in order to measure the ascending and descending thoracic aortic diameter and cross-sectional area achieved high accuracy, which allowed for the quantification of aortic strain and distensibility in 42,342 participants in the UK Biobank. Downstream analyses recapitulated well established observations from the literature, particularly that of the inverse relationship between age and distensibility. Because reduced strain corresponds with advanced age, the observation that prevalent CAD was associated with increased strain was surprising. However, the expected inverse association between CAD and distensibility was recovered with three separate approaches: one, by demonstrating an inverse association between distensibility assessed prior to the diagnosis of CAD; two, by conducting polygenic analyses of strain and distensibility, which confirmed that genetic predictions of increased strain and distensibility were associated with a lower incidence of CAD; and three, by estimating and accounting for the effect of medications on aortic strain and distensibility. Our estimates of medication effect size were not causal, because they were confounded by indication. However, the analyses did suggest that the effects of some medications may be perceptible by measuring aortic strain and distensibility after even a short period of therapy during which small changes in aortic diameter would be difficult to detect (average follow-up time between UK Biobank MRI visits: 2.2 years). This extends prior observations about beta blockade to include estimates for several additional medication classes (**Supplementary Table 6**)^6^. The consistent inverse relationship between aortic distensibility and diameter across our epidemiological and genetic analyses (**Figure 1D**; **Supplementary Figure 7**) raises the possibility of using aortic distensibility as an intermediate phenotype for changes in aortic diameter.

Although diameter and distensibility were consistently inversely correlated with one another, aortic strain and distensibility were not merely redundant with respect to diameter. The GWAS for distensibility found associations at 12 loci for the ascending aorta and 11 loci for the descending aorta, and a greater number were identified for strain (26 for the ascending aorta and 21 for the descending aorta). 22 of these loci were specific to strain or distensibility, and were not identified at genome-wide significance in a thoracic aortic diameter GWAS within the same samples. Examination of single nucleus sequencing data from aortic tissue revealed enrichment of genes near GWAS loci in vascular smooth muscle cells, the primary cell type responsible for homeostasis of extracellular matrix function in the aorta. Multiple genes implicated by this methodology are in fact regulators or constituents of the extracellular matrix, responsible for the elastic properties of the aorta. Additionally, while the polygenic scores had very modest effect sizes in predicting outcomes such as coronary artery disease, chronic kidney disease, and stroke, they remained statistically significant predictors even after accounting for the aortic diameter polygenic scores. These findings underscore the notion that strain and distensibility add information beyond that which is obtained by assessing only aortic diameter.

In a separate UK Biobank aortic diameter and distensibility GWAS that initially found 10 genetic loci associated with ascending aortic distensibility and 7 with descending aortic distensibility, the investigators applied a multi-trait meta-analytic approach called MTAG^33^, gaining 22 additional genetic loci for distensibility^34^. In the present study, we did not report MTAG-enhanced results because we found MTAG to have a high maximum false discovery rate (maxFDR) for our aortic measurements (maxFDR of 15% for ascending and 25% for descending distensibility; **Supplementary Note**), which is more likely to occur in scenarios where the analyzed traits have a power imbalance^33^. Because of the modest heritability of distensibility, we also evaluated aortic strain, which had a higher heritability and which would not be affected by the limitations with central pulse pressure estimates that we attempted to address with our quality control approach.

Interesting genetic variants associated with ascending aortic strain included rs9687138, which is an expression quantitative trait locus (eQTL) for *ANKH*: the A allele was associated with increased *ANKH* expression in GTEx v8, as well as with increased aortic strain in our analysis, consistent with previously established biology wherein the *ANKH* protein product inhibits vascular calcification^25,35^. Other strain-associated loci not found in the diameter GWAS included those near *ISL1*, which plays a role in the development of the secondary heart field and aortic arch formation^36^; and *ADAMTS9*, whose protein product is also expressed in tissues of the second heart field^37^. The TWAS of aortic strain highlighted another protease, *ADAMTS7*, which is a known mediator of atherogenesis and VSMC behavior^38,39^. Finally, four aortic strain loci had previously been associated with aortic root diameter in an echocardiography-based GWAS (near *CFDP1*, *GOSR2*, *PRDM6*, and *SMG6*). For each of the four lead SNPs near these genes, the allele associated with a greater aortic root diameter also corresponded to a lesser ascending aortic strain (**Supplementary Table 19**)^40^.

Several additional loci that were significantly associated with strain and distensibility in the present study were noteworthy. Approximately ∼40% of the dry weight of the aorta is comprised of a single protein, elastin^41^, the product of the *ELN* gene on chromosome 7. Monomeric tropoelastin protein spontaneously assembles into large macromolecular fibers and sheets in a phase transition process called ‘coacervation’, which is rendered irreversible through the activity of multiple cross linking enzymes^42^. The macromolecular product of this process, termed ‘amorphous elastin’, is ubiquitous in the body and responsible for the resiliency of nearly all tissues that undergo repetitive biomechanical deformation, notably the aorta, skin, and lung. Perhaps unsurprisingly, the gene *ELN* was implicated in this GWAS as a determinant of ascending and descending aortic strain and distensibility. In both animal^43^ and *ex vivo* human models^44^, elastin has been shown to be the primary mediator of resiliency to aortic deformation at physiologic blood pressures. Similarly the gene *FBLN5*, the introns of which contain lead SNPs for descending strain and distensibility, encodes fibulin-5, a mediator of elastogenesis which recruits tropoelastin and elastin cross linking enzymes such as lysyl oxidase like 1 (LOXL1) to microfibrils to direct elastin fiber assembly and maturation^45–47^. Homozygous mutations in *FBLN5* cause autosomal recessive cutis laxa in humans, a disease of failed elastogenesis^48^. *Fbln5*^-/-^ mice have been proposed as a model of vascular aging secondary to the phenotype of early onset aortic stiffness and increased pulse wave velocity. Our findings lend support to this proposition in humans. A locus near *LTBP4*, which is an *FBLN5* binding partner that also binds to TGF-β and participates in elastogenesis^49,50^, was also associated with descending aortic strain.

Elastogenesis is completed in adolescence and amorphous elastin has half-life of 70 years, suggesting that some loci found in our study (ie. *ELN* and *FBLN5*) likely affect aortic distension in a congenital manner. However, other loci harbor genes encoding protein products whose known function suggests that they may act on shorter timescales, which may be relevant to an acquired loss of aortic distensibility. *PI15* is a strong candidate for a disease causing gene near a lead SNP on chromosome 8, as loss of this gene product induces aortic elastin defects and ruptures in rodents^51^, and is downregulated in experimental hypertension^52^. *PI15* encodes an extracellular peptidase inhibitor with limited activity against trypsin, although the actual target of its inhibitory activity in the aortic wall is currently unknown and may be an important target. Additionally, *HAS2* near rs11992999 on chromosome 8 encodes a hyaluronic synthetase producing the mucopolysaccharide hyaluronic acid, known to affect the proliferation of VSMCs^53,54^ and modulate the mechanical properties of tissues^55^. A lead SNP for descending aortic distensibility was also on the transcript of another gene encoding an enzyme involved in polysaccharide production (*CHSY1*, encoding chondroitin sulfate synthetase 1), expression of which is induced in aortic smooth muscle cells by TGF-β^56^.

Finally, several lines of evidence converged on the locus near *PLCE1*. SNPs at the locus were found to be associated with aortic diameter, strain, and distensibility (**Supplementary Figure 13**). MAGMA highlighted both *PLCE1* at the locus for diameter, strain, and distensibility, and *NOC3L* at the locus for diameter (**Figure 5**, and **Supplementary Figure 14**). The association pattern was complex, with two conditionally independent signals driving the diameter association, only one of which (a block that included rs1343094, an eQTL for *PLCE1* in the tibial artery) was also significantly associated with strain and distensibility. The diameter-specific signal, near rs35247409, was an eQTL for the nearby *NOC3L*; a genetic prediction of decreased expression of *NOC3L* was associated with an increase in aortic diameter. This signal was also in linkage disequilibrium with the CAD risk signal seen by Aragam, *et al*^57^, oriented such that the CAD risk-decreasing allele was aligned with the rs35247409 aortic diameter-increasing allele (**Supplementary Figure 15**). For the second signal at the locus, a genetic prediction of decreased expression of *PLCE1* was associated with an increased aortic diameter and decreased strain and distensibility (**Supplementary Figure 15**). In alignment with this expression-based expectation, LoF variants in *PLCE1* were significantly associated with decreased aortic distensibility in the exome-wide analysis. *PLCE1* encodes phospholipase C ε which generates second messengers diacylglycerol and inositol 1,4,5 triphosphate^58^. It interacts with advillin and plays a role in the regulation of actin cytoskeleton^59,60^. In aortic single nucleus sequencing data, it is expressed in smooth muscle and endothelial cells, among other cell types (**Supplementary Figure 10**). However, loss of function mutations in *PLCE1* are known to cause steroid-resistant nephrotic syndrome in humans^61^, and therefore the effect on aortic distensibility may or may not be mediated locally.

In conclusion, we have measured ascending and descending thoracic aortic strain and distensibility in up to 42,342 UK Biobank participants, investigated their epidemiology, and identified 41 significantly associated loci, many of which have established connections to TGF-β signaling, elastogenesis, or polysaccharide regulation.

### Limitations

This study has several limitations. All aortic measurements were derived from deep learning models of cardiovascular MRI at the level of the right pulmonary artery. The deep learning models have not been tested outside of the specific devices and imaging protocols used by the UK Biobank and are unlikely to generalize to other data sets without fine tuning. The study population was largely composed of people of European ancestries, limiting generalizability of the findings to global populations. The participants who underwent MRI in the UK Biobank tended to be healthier than the remainder of the UK Biobank population, which itself is likely to be healthier than the general population. CPP, used with strain to compute distensibility, was estimated by a validated device and not directly measured; the quality control procedures that we developed for CPP were heuristic. The effect of medication classes on aortic phenotypes was estimated based on a biased sample, because these medications were initiated due to a medical indication—not at random. Currently, there is little follow-up time subsequent to the initial MRI visit for most UK Biobank participants, limiting our ability to reliably assess the relationship between cardiovascular measurements and disease-based outcomes. Confirmation of the rare variant association between *PLCE1* loss-of-function and decreased descending aortic strain will require external replication.

## Methods

### Study design

All population-level analyses were conducted in the UK Biobank, which is a richly phenotyped, prospective, population-based cohort that recruited 500,000 participants aged 40-69 years in the UK via mailer from 2006-2010^62^. We analyzed 42,342 participants with imaging data who had not withdrawn consent as of February 2020. Access was provided under application #7089 and approved by the Partners HealthCare institutional review board (protocol 2003P001563).

### Statistical analysis

Statistical analyses were conducted with R 3.6.3 (R Foundation for Statistical Computing, Vienna, Austria). Survival analyses and accelerated failure time analyses were conducted with Cox proportional hazards models in R ^63^.

### Cardiovascular magnetic resonance imaging protocols

The UK Biobank is conducting an imaging substudy on 100,000 participants which is currently underway^14,64^. Cardiac magnetic resonance imaging was performed with 1.5 Tesla scanners (MAGNETOM Aera, Siemens Healthcare), using electrocardiographic gating for cardiac synchronization^14^. A balanced steady-state free precession cine, consisting of a series of exactly 100 images throughout the cardiac cycle, was acquired for each participant at the level of the right pulmonary artery^14^.

### Semantic segmentation and quality control

Segmentation maps were traced for the ascending and descending thoracic aorta manually by a cardiologist (JP). To produce the final model used in this manuscript, 501 samples were chosen, manually segmented, and used to train a deep learning model with fastai v1.0.59^12^. The model consisted of a U-Net-derived architecture, where the encoder was a resnet34 model pre-trained on ImageNet^11,12, 65–67^. 80% of the samples were used to train the model, and 20% were used for validation.

During training, all images were resized to be 160 pixels in width by 132 pixels in height for the first half of training (‘small image training’), and then 240 pixels in width by 196 pixels in height, which is the native size of these images, for the second half (‘large image training’), detailed below. The Adam optimizer was used, and the model was trained with a minibatch size of 4 (when training with small images) or 2 (when training with large images)^68^. Rather than using extensive hyperparameter tuning with a grid search, the model was instead trained using a cyclic learning rate training policy, which alternately decreases and increases the learning rate during training^69^.

The maximum learning rate (the step size during gradient descent) was chosen with the learning rate finder from the FastAI library ^12^. Weight decay was set to 0.001 (whereas the FastAI default is 0.01). Small image training was performed for 15 epochs, with a maximum learning rate of 2E-03, and with 20% of the iterations permitted to have an increasing learning rate during each. Training was performed while keeping all ImageNet-pretrained layers fixed, so that only the final layer was fine-tuned. Then all layers were unfrozen and the model was trained for an additional 15 epochs with a maximum learning rate that varied during training from 2E-05 to 2E-03, with 40% of the iterations permitted to have an increasing learning rate during each epoch. For large image training, the same model was then updated using full-dimension images, and with the maximum learning rate set to 2E-04, with 30% of the iterations permitted to have an increasing learning rate over 8 epochs. Finally, all layers were unfrozen and the model was trained for an additional 15 epochs with a maximum learning rate ranging from 2E-05 to 2E-04.

Throughout training, augmentations (random perturbations of the images) were applied as a regularization technique. These augmentations included affine rotation, zooming, and modification of the brightness and contrast. We did not permit mirroring transformations.

This model was then used to infer segmentation of the ascending and descending aorta on all available “CINE_segmented_Ao_dist” images in the UK Biobank. During inference, adaptive pooling was used to permit arbitrary image sizes^70^, which allows for the production of output that matches the input size, preserving the number of millimeters per pixel as reported in the metadata.

Segmentation accuracy was assessed for the ascending and descending aortic annotations. The Sørensen-Dice coefficient (ranging from 0 for perfect disagreement to 1 for perfect agreement) was used to compare the pixel-wise output of the deep learning model to manual annotations within held-out test images that were not used in training or validation^71,72^. This coefficient was computed for each of the held-out test images, and the mean coefficient was computed from these results.

### Diameter, strain, and distensibility

The ascending and descending thoracic aortic cross-sectional areas were converted from pixel counts (which were produced by the deep learning model) to absolute areas (cm^2^) by using the metadata accompanying each image that defined the width and height of each pixel. To measure aortic strain, the greatest and least areas throughout the cardiac cycle were taken. Strain was defined as the change in area (greatest area minus least area) divided by the least area. This process was repeated for ascending and descending thoracic aortic luminal areas. To produce estimates of distensibility, the ascending and descending thoracic aortic strain values were divided by the Vicorder-based central pulse pressure estimates, which were derived from brachial blood pressure measurements during the MRI visit^2,13,14^. The central pulse pressure values required quality control steps, detailed in the **Supplementary Note**.

We also produced an updated set of ascending and descending aortic diameter measurements based on the same deep learning model, for the purpose of conducting a secondary GWAS to allow clarification of which strain and distensibility GWAS loci would not have been discovered in a set of diameter GWASes within the same sample. We used the same procedure to estimate diameter as in our previously reported diameter GWAS^27^: major and minor axes were computed from the deep learning-derived segmentation output by using classical image moment algorithms^73^. The length of the minor elliptical axis (i.e., the diameter, measured in pixels) was ascertained at the point in the cardiac cycle when the aorta was at its maximum size. This length of the minor axis was converted to an absolute length in centimeters by using the metadata accompanying each image.

### Definitions of curated cardiovascular diseases

We defined a set of curated cardiovascular diseases for epidemiological analyses and for exclusion criteria based on self report, ICD codes, and procedural codes (**Supplementary Table 4**). The data were obtained from the UK Biobank in June 2020, at which time the recommended phenotype censoring date was March 31, 2020. The UK Biobank defines that date as the last day of the month for which the number of records is greater than 90% of the mean of the number of records for the previous three months (https://biobank.ndph.ox.ac.uk/ukb/exinfo.cgi?src=Data_providers_and_dates).

### Disease-based epidemiological analyses

Association analyses between cardiovascular diseases (as a binary independent variable) and the aortic phenotypes (as the dependent variable) were performed using linear models that also accounted for the MRI serial number, sex, the first five principal components of ancestry, age at enrollment, age and age^2^ at the time of MRI, and the genotyping array.

Incident disease analyses were performed with Cox models that evaluated survival to censoring or first diagnosis of a disease with the R *survival* package^63^. The model was run separately for each disease, repeated to include each aortic phenotype in the model for each disease. The model was adjusted for the MRI serial number, sex, the first five principal components of ancestry, age and age^2^ at the time of MRI, and the genotyping array. P values for violations of the proportional hazards assumptions are provided for the model globally and for the aortic phenotype term within the model^74^.

### Medication adjustment

Medications taken by UK Biobank participants were previously mapped to ATC codes^17,75^. We grouped medications by the third level of their ATC code, which approximately groups medications by therapeutic type (e.g., “beta blocking agents”).

To produce estimates of the relationships between these third-level medication groupings and aortic phenotypes, we analyzed the 2,712 UK Biobank participants who had undergone a second imaging visit, which was on average 2.2 years subsequent to the first. For each class, all participants who were not on that medication class during the first imaging visit were analyzed, and the linear model predicted the aortic phenotype at the second visit as a function of the aortic phenotype at the first visit, plus age at the first visit, sex, the time elapsed between the first and second visits, and a binary flag indicating whether or not the participant started a medication in the ATC class between the two visits. The coefficient of the binary flag represents an additive effect size estimate of the medication on the aortic phenotype, although this is not a causal estimate.

### PheCode analyses

We tested for association between the aortic phenotypes and all available PheCode-based disease labels, which were derived from ICD-10 codes and OPCS-4 codes^18^. The model was formulated as a linear model with the aortic phenotype scaled so that its mean was zero with a standard deviation of one as the outcome variable, to allow for comparability of disease effect across phenotypes. Independent variables included the presence or absence of disease as a binary variable, and clinical covariates that included MRI serial number, sex, the first five principal components of ancestry, age at enrollment, the cubic natural spline of age at the time of MRI, and the genotyping array. This test was repeated for each disease and for each phenotype.

A second PheCode-based analysis was conducted after adjusting each aortic phenotype for all medication classes that achieved P < 0.05 in the univariate analyses described in the **‘Medication adjustment’** section above. For example, if a medication had an estimated effect size on ascending aortic diameter of -0.05cm, then for this analysis, participants taking this medication would have their aortic diameter adjusted by adding 0.05cm.

### Genotyping, imputation, and genetic quality control

UK Biobank samples were genotyped on either the UK BiLEVE or UK Biobank Axiom arrays and imputed into the Haplotype Reference Consortium panel and the UK10K+1000 Genomes panel^76^. Variant positions were keyed to the GRCh37 human genome reference. Genotyped variants with genotyping call rate < 0.95 and imputed variants with INFO score < 0.3 or minor allele frequency <= 0.005 in the analyzed samples were excluded. After variant-level quality control, 11,622,901 imputed variants remained for analysis.

Participants without imputed genetic data, or with a genotyping call rate < 0.98, mismatch between self-reported sex and sex chromosome count, sex chromosome aneuploidy, excessive third-degree relatives, or outliers for heterozygosity were excluded from genetic analysis^76^. Participants were also excluded from genetic analysis if they had a history of aortic aneurysm, dissection, rupture, or procedures documented prior to the time they underwent cardiovascular magnetic resonance imaging at a UK Biobank assessment center. Our definitions of these diseases in the UK Biobank are provided in **Supplementary Table 4**. Participants were also excluded if they had a measured aortic diameter > 5cm.

### Heritability and genome-wide association studies

For aortic diameter, strain, and distensibility, a rank-based inverse normal transformation was applied^77^. All GWAS were adjusted for sex, age at enrollment, age and age^2^ at the time of MRI, the first 10 principal components of ancestry, the genotyping array, and the MRI scanner’s unique identifier.

BOLT-REML v2.3.4 was used to assess the SNP-heritability of the phenotypes, as well as their genetic correlation with one another using the directly genotyped variants in the UK Biobank^19^.

Genome-wide association studies for each phenotype were conducted using BOLT-LMM version 2.3.4 to account for cryptic population structure and sample relatedness^19,20^. We used an autosomal panel of 714,477 directly genotyped SNPs that passed quality control to construct the genetic relationship matrix (GRM), with covariate adjustment as noted above. Associations on the X chromosome were also analyzed, using all autosomal SNPs and X chromosomal SNPs to construct the GRM (N=732,116 SNPs), with the same covariate adjustments and significance threshold as in the autosomal analysis. In this analysis mode, BOLT treats individuals with one X chromosome as having an allelic dosage of 0/2 and those with two X chromosomes as having an allelic dosage of 0/1/2. Variants with association P < 5E-8, a commonly used threshold, were considered to be genome-wide significant.

Linkage disequilibrium score regression was conducted on the summary statistics for each trait using *ldsc* version 1.0.1 with its default settings^78^. This yielded the *ldsc* intercept and attenuation ratio, which help distinguish polygenicity from confounding biases, as well as a second estimate of heritability for each trait.

We identified lead SNPs for each trait. Linkage disequilibrium (LD) clumping was performed with PLINK-1.9^79^ using the same participants used for the GWAS. We outlined a 5-megabase window (--clump-kb 5000) and used a stringent LD threshold (--r2 0.001) in order to account for long LD blocks. With the independently significant clumped SNPs, distinct genomic loci were then defined by starting with the SNP with the strongest P value, excluding other SNPs within 500kb, and iterating until no SNPs remained. Independently significant SNPs that defined each genomic locus were termed the lead SNPs. To allow for identification of the same locus shared across traits, even when the traits’ lead SNPs at the same locus were different, we labeled the GWAS loci in the supplementary tables with a “Global Locus ID”.

No aortic strain or distensibility lead SNP deviated from Hardy-Weinberg equilibrium (HWE) at a threshold of P < 1E-06^79^.

### TWAS

For each strain and distensibility GWAS, we performed a TWAS to identify genes whose imputed cis-regulated gene expression corrrelates with aortic strain and distensibility^23,24,80,81^. We used PrediXcan with aortic eQTL data from GTEx v8^22,25^. Genes were considered significant based on a per-trait Bonferroni significance threshold.

### MAGMA

We used MAGMA version 1.09b, which addresses limitations in the statistical assumptions made in versions prior to 1.08^21,82,83^. MAGMA was run with its default settings. We used the default gene and SNP annotations keyed to GRCh37, based on a European ancestry panel from the 1000 Genomes Project, which were precomputed by the MAGMA authors. Genes were considered significant based on a per-trait Bonferroni significance threshold.

### Single nucleus gene expression lookups

We conducted lookups of gene expression within 12 cell-type clusters defined with single nucleus sequencing that was previously performed using samples from ascending and descending aorta from 3 humans which were publicly available (Gene Expression Omnibus Database #GSE165824 and Broad Institute Single Cell Portal #SCP1265)^27^. Those 12 cell populations included vascular smooth muscle cells, fibroblasts, three distinct types of endothelial cells, as well as macrophages and lymphocytes. The #SCP1265 database includes, for each human gene, the pre-computed log fold-change and P value for the gene in each cell type compared to all other cell types using the limma-voom framework, adjusting for sample of origin^84^. In the present work, we selected genes associated with aortic distensibility (either via the TWAS or due to being the nearest gene to a GWAS lead SNP) and looked up their log fold-change and P value for each cell type.

### Exome-based rare variant association test

We used whole exome sequencing (WES) on 200,000 UK Biobank participants, updated from the initial 50,000-participant release^85^. As detailed in prior work, samples were restricted to those who also had high-quality genotyping array data available. Additional filters were applied to study samples and autosomal exome sequence variants: sample call rate (<90%), genotype call rate (<90%), Hardy-Weinberg equilibrium test (P< 1E-14 among unrelated individuals) and minor allele count. Of the 200,643 individuals in the UK Biobank with WES who passed the internal quality-control, we excluded additional samples due to withdrawn consent, duplicates, sex-mismatch, low-callrate (<90%), and outliers (in the transition/transversion ratio, heterozygous/homozygous ratio, SNP/indel ratio, or number of singletons), leaving 200,337 individuals. Of these, up to 22,487 also had aortic imaging data.

The protein consequences of variants were annotated using dbNSFP4.1a and the Loss-of-Function Transcript Effect Estimator (LOFTEE) plug-in implemented in the Variant Effect Predictor (VEP) (https://github.com/konradik/loftee)^86,87^. We removed any loss-of-function variants (LOFs) flagged by LOFTEE as dubious, as described in prior work^88^.

We conducted gene-based LOF burden testing using a linear mixed-effects model that was adjusted for the fixed effects of age at MRI, sex, principal components of ancestry, sequencing batch, and MRI serial number, as well as the mixed effect of the empirical kinship matrix using the R package *GENESIS*^89^. We aggregated LOF variants in each gene, and tested exome-wide associations between carriers and aortic phenotypes with a score test. If the cumulative minor allele count was less than 10 for a gene, the gene was excluded. The Bonferroni multiple testing threshold was applied to assess significance.

### Polygenic score analyses

We computed polygenic scores using the software program PRScs (version 43128be7fc9ca16ad8b85d8754c538bcfb7ec7b4) with a UK Biobank European ancestry linkage disequilibrium panel made publicly available by the software authors^28^. The PRScs method applies a continuous shrinkage prior to the SNP weights. PRScs was run in ‘auto’ mode on a per-chromosome basis. This mode places a standard half-Cauchy prior on the global shrinkage parameter and learns the global scaling parameter from the data; as a consequence, PRScs-auto does not require a validation data set for tuning. Based on the software default settings, only the 1,117,425 SNPs found at HapMap3 sites that were also present in the UK Biobank were permitted to contribute to the score. These scores were applied to the entire UK Biobank.

The aortic strain and distensibility scores were tested for association with the curated cardiovascular diseases described above. This was first attempted using Cox proportional hazards models as implemented by the R *survival* package^63^. However, significant violations of the proportional hazards assumptions were present for some of the most common cardiovascular diseases (such as hypertension and CAD) even before the addition of the polygenic scores to the Cox models. Therefore, we pursued accelerated failure time analyses, which make the assumption that the effect of a covariate, such as a polygenic score, is to accelerate or decelerate progression towards a disease^29^. Accelerated failure time analysis was pursued with the *survreg* function within the same R *survival* package, using the log-logistic distribution for all models.

Participants related within 3 degrees of kinship to those who had undergone MRI, based on the precomputed relatedness matrix from the UK Biobank, were excluded from analysis^76^. We conducted these analyses in individuals who were genetic inliers for European ancestries based on the first three pairs of genetic principal components (PC1&2, PC3&4, PC5&6) by using the aberrant package as described previously^76,90^. We also excluded individuals with disease that was diagnosed prior to enrollment in the UK Biobank. We counted survival as the number of years between enrollment and disease diagnosis (for those who developed disease) or death, loss to follow-up, or end of follow-up time (for those who did not develop disease). We adjusted for covariates including sex, the cubic basis spline of age at enrollment, the interaction between the cubic basis spline of age at enrollment and sex, the genotyping array, and the first five principal components of ancestry. For models testing the effect of the ascending aortic strain or distensibility polygenic score on disease risk, we also repeated the analysis after adjustment for the ascending aortic diameter polygenic score. A parallel adjustment was conducted for the descending thoracic aorta polygenic scores.

## Appendices

### Data availability

UK Biobank data are made available to researchers from research institutions with genuine research inquiries, following IRB and UK Biobank approval. GWAS summary statistics and polygenic score weights will be available upon publication at the Broad Institute Cardiovascular Disease Knowledge Portal (http://www.broadcvdi.org). Single nucleus sequencing data were retrieved from the Broad Institute Single Cell Portal #SCP1265. Aortic diameter, strain, and distensibility measurements will be returned to the UK Biobank for use by any approved researcher. All other data are contained within the article and its supplementary information.

### Author contributions

PTE, MEL, and JPP conceived of the study. JPP and MN annotated images. JPP trained the deep learning models. JPP, SHC, and MDC conducted bioinformatic analyses. JPP, MDC, SHC, MEL, and PTE wrote the paper. All other authors contributed to the analysis plan or provided critical revisions.

### Sources of funding

Dr. Pirruccello was supported by a Sarnoff Scholar Award and National Institutes of Health (NIH) grant K08HL159346. Dr. Lindsay was supported by the Fredman Fellowship for Aortic Disease, the Toomey Fund for Aortic Dissection Research, and NIH R01HL130113. Dr. Ellinor was supported by the Fondation Leducq (14CVD01), a grant from the American Heart Association Strategically Focused Research Networks, MAESTRIA (965286), and by grants from the NIH (R01HL092577, R01HL128914, K24HL105780). Dr. Chou was funded by NIH #T32HL007208. This work was funded by a collaboration between the Broad Institute and IBM Research.

### Disclosures

Dr. Pirruccello has served as a consultant for Maze Therapeutics and has received research support from IBM Research. Dr. Juric is supported by grants from Genentech, Eisai, EMD Serono, Takeda, Amgen, Celgene, Placon Therapeutics, Syros, Petra Pharma, InventisBio, Infinity Pharmaceuticals, and Novartis. Dr. Juric has also received personal fees from Genentech, Eisai, EMD Serono, Ipsen, Syros, Relay Therapeutics, MapKure, Vibliome, Petra Pharma, and Novartis. Dr. Batra receives sponsored research support from Bayer AG and IBM, and has consulted for Novartis and Prometheus Biosciences. Dr. Ellinor is supported by a grant from Bayer AG to the Broad Institute focused on the genetics and therapeutics of cardiovascular diseases. Dr. Ellinor has also served on advisory boards or consulted for Bayer AG, Quest Diagnostics, MyoKardia and Novartis. The Broad Institute has filed for a patent on an invention from Drs. Ellinor, Lindsay, and Pirruccello related to a genetic risk predictor for aortic disease. The remaining authors report no disclosures.

## Data Availability

GWAS summary statistics and polygenic score weights will be available upon publication at the Broad Institute Cardiovascular Disease Knowledge Portal (http://www.broadcvdi.org). Aortic diameter, strain, and distensibility measurements will be returned to the UK Biobank for use by any approved researcher. All other data are contained within the article and its supplementary information.

https://singlecell.broadinstitute.org/single_cell/study/SCP1265/deep-learning-enables-genetic-analysis-of-the-human-thoracic-aorta

