## Supplementary Note and Figures for "The Genetic Determinants of Aortic Distension"

### Supplementary Material

**Supplementary Material**	**1**

Supplementary Note	3

Central pulse pressure and distensibility quality control	3

Multi-trait analysis of GWAS (MTAG)	4

Supplementary References	5

Supplementary Figures	6

Supplementary Figure 1: Sample flow diagram	7

Supplementary Figure 2: Strain and distensibility distributions	8

Supplementary Figure 3: Continuous phenotypes associated with aortic distensibility and strain	9

Supplementary Figure 4: Association between ATC medication classes and aortic phenotypes	10

Supplementary Figure 5: PheCodes prior to adjusting for medications	11

Supplementary Figure 6: PheCodes after adjusting for medications	12

Supplementary Figure 7: Cross-trait genetic correlation	13

Supplementary Figure 8: GWAS QQ plots	14

Supplementary Figure 9: GWAS locus plots	15

Supplementary Figure 10: Single nucleus sequencing	56

Supplementary Figure 11: Rare variant association testing	57

Supplementary Figure 12: Polygenic scores	58

Supplementary Figure 13: PLCE1 ascending aorta locus plots	59

Supplementary Figure 14: Gene-based MAGMA for aortic diameter	60

Supplementary Figure 15: PLCE1 locus, common genetic variant data	61

Supplementary Figure 16: Distensibility without QC	62

Supplementary Figure 17: Strain without QC	63

Supplementary Figure 18: Central pulse pressure without QC	64

Supplementary Figure 19: Central pulse pressure by SBP/ESP	65

Supplementary Figure 20: Central pulse pressure by SBP/ESP over time	66

Supplementary Figure 21: Distensibility after QC	67

#### Supplementary Note

##### Central pulse pressure and distensibility quality control

Aortic distensibility is aortic strain divided by central pulse pressure. Without quality control, the distensibility estimates are heavily right-skewed (**Supplementary Figure 16**). In contrast, the aortic strain estimates are only modestly right-skewed (**Supplementary Figure 17**), pointing to the centrally provided central pulse pressure (CPP) measurements as a potential explanation. The distribution of the CPP measurements is notable for a second mode below approximately 22.5mmHg (**Supplementary Figure 18**), which would not be expected physiologically. Plotting the CPP as a function of central systolic blood pressure divided by central end systolic pressure yielded revealed what appeared to be a mixture of three different distributions of points, the most extreme of which had negative estimates for cardiac output from the Vicorder recording, which are not physically possible and were therefore set to missing values (**Supplementary Figure 19**). After excluding those values, the remaining data still appeared to represent a mixture of two distributions of points. Notably, when plotted over the course of time, the right-shoulder distribution did not exist in the earliest imaging studies obtained in the UK Biobank (**Supplementary Figure 20**). We speculated that this right shoulder might be a systematic misreading by the device of the central pulse pressure, or possibly a second device that is calibrated differently, although we were not able to conclusively determine the explanation for the second shoulder.

Based on the above, we developed the following heuristic CPP inclusion criteria from the above observations:

1. CPP > 22.5mmHg
2. Central systolic blood pressure > 85mmHg
3. Cardiac output estimate > 0 and must be non-missing
4. CPP >= 200 * (central systolic blood pressure / central end systolic pressure) - 200

Based on the application of those inclusion criteria, approximately 10% of distensibility values were rejected, and the distribution of aortic distensibility, without any additional filtering, took on a much less extreme skew (**Supplementary Figure 21**).

##### Multi-trait analysis of GWAS (MTAG)

MTAG is an analytic method that conducts meta-analysis of genome-wide association study (GWAS) summary statistics across multiple correlated traits, yielding updated summary statistics for each trait^1^. The MTAG software provides the ability to compute a maximum false discovery rate (maxFDR), which is an estimate of the upper bound for the FDR under several of the software’s assumptions which include, among others, that (a) effect sizes follow a spike-and-slab distribution, and (b) 10% of single nucleotide polymorphisms are non-null for each trait^1^.

We applied MTAG 1.0.8 to the BOLT-LMM results for the three ascending aortic traits (diameter, strain, and distensibility), and separately for the three descending aortic traits. The --fdr flag was passed, causing the software to compute the maxFDR values for each trait. Additionally, the --no-allele-flipping and --incld-ambig-snps flags were passed, because all of the summary statistics came from the same software and the same population. Otherwise, the default settings were used.

For the ascending aortic traits, MTAG yielded a maxFDR of 1.0% for diameter, 7.6% for strain, and 14.7% for distensibility. For the descending aortic traits, the maxFDR was 1.9% of diameter, 9.2% for strain, and 25.5% for distensibility. The MTAG authors described maxFDR levels above 15% as “concerning.”^1^

#### Supplementary Figures

##### Supplementary Figure 1: Sample flow diagram

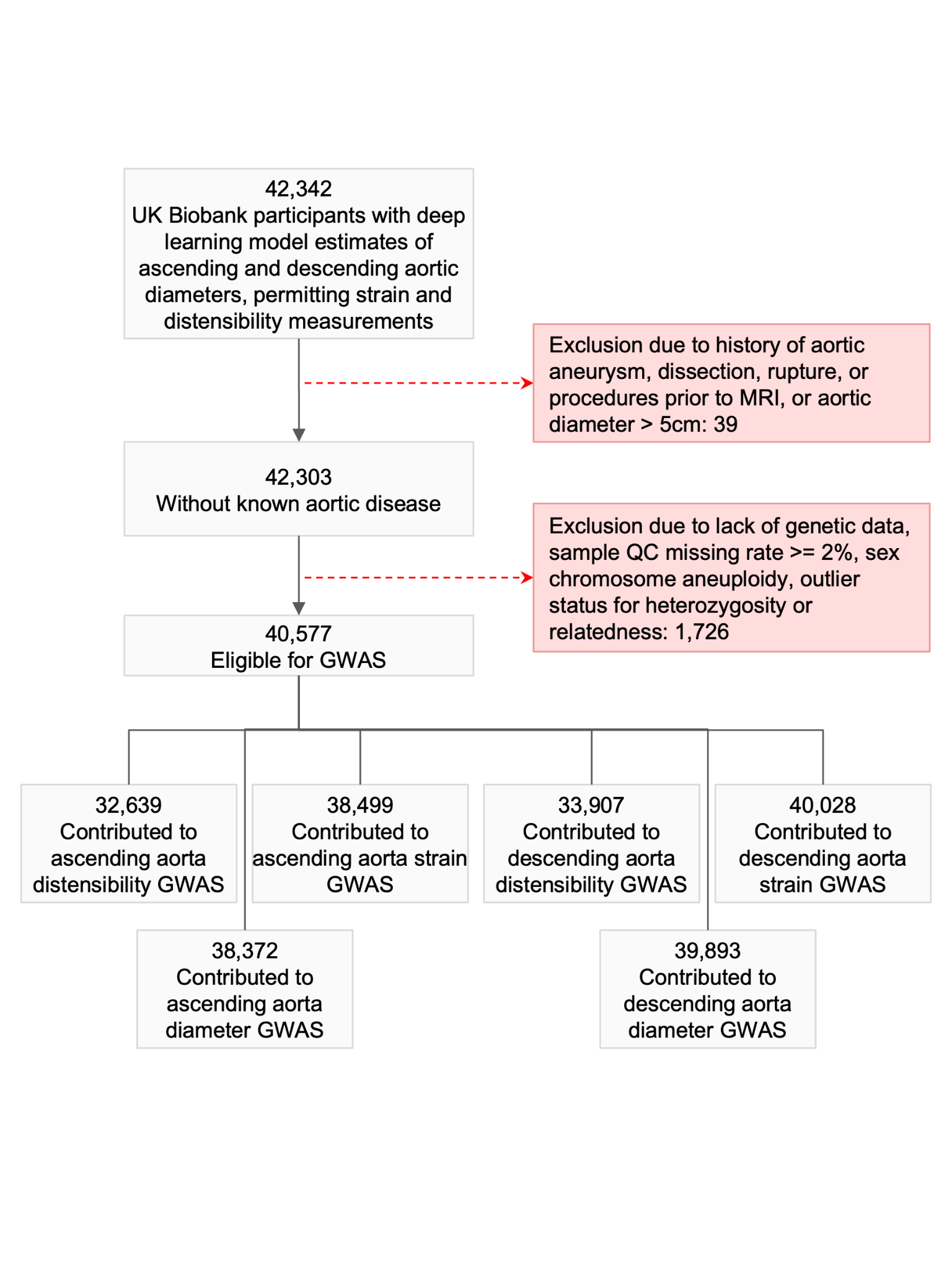

##### Supplementary Figure 2: Strain and distensibility distributions

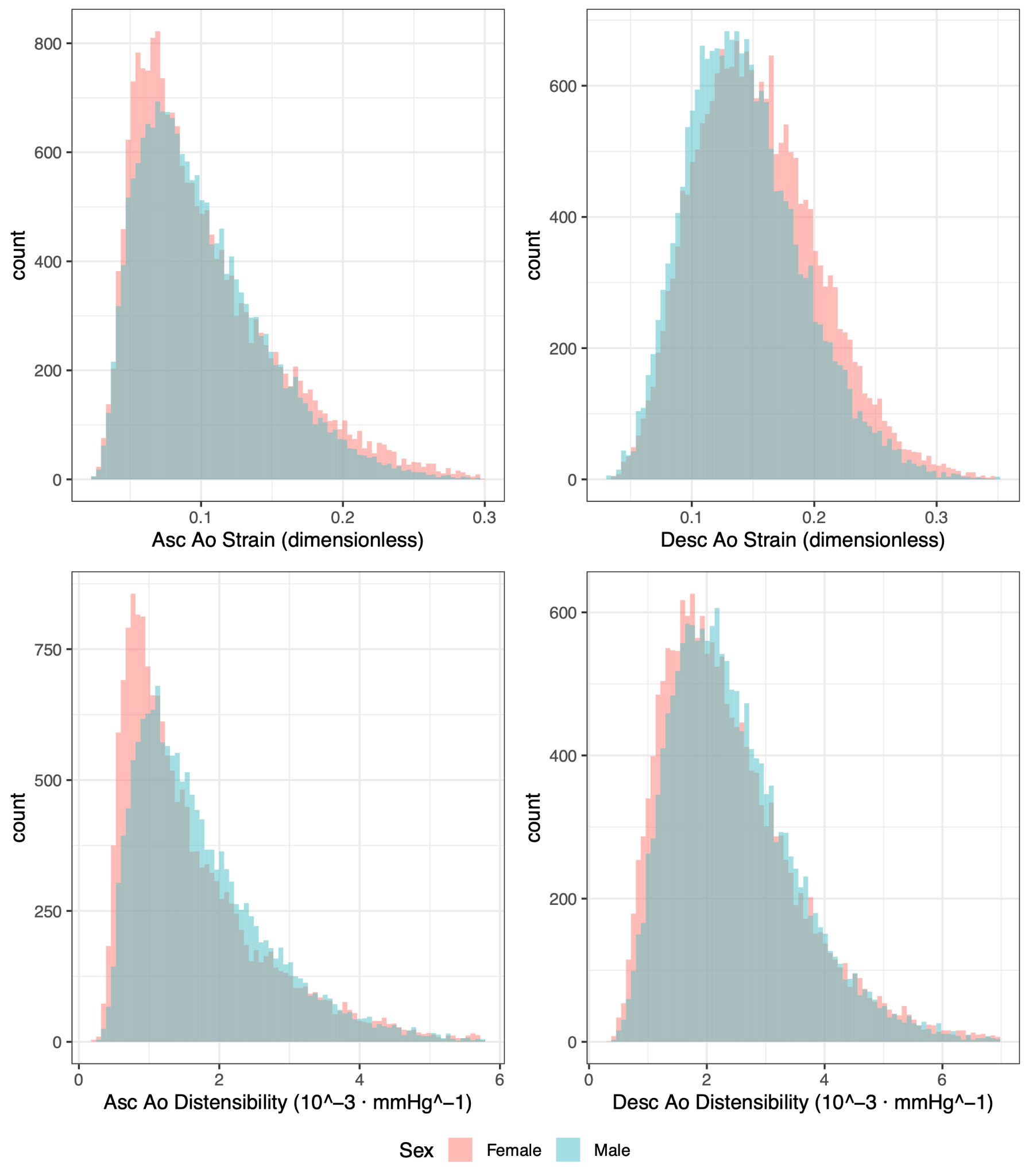

##### Supplementary Figure 3: Continuous phenotypes associated with aortic distensibility and strain

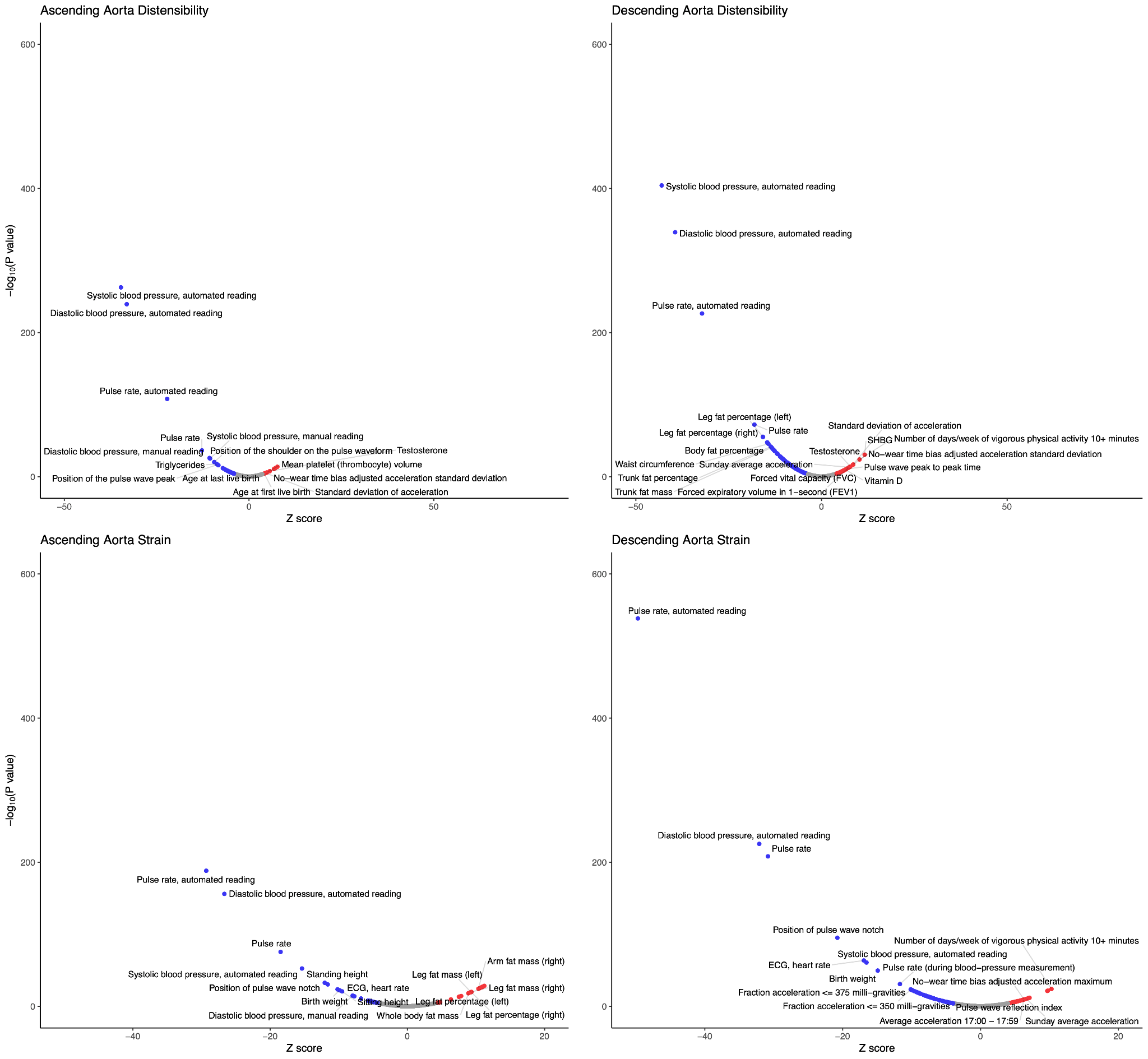

##### Supplementary Figure 4: Association between ATC medication classes and aortic phenotypes

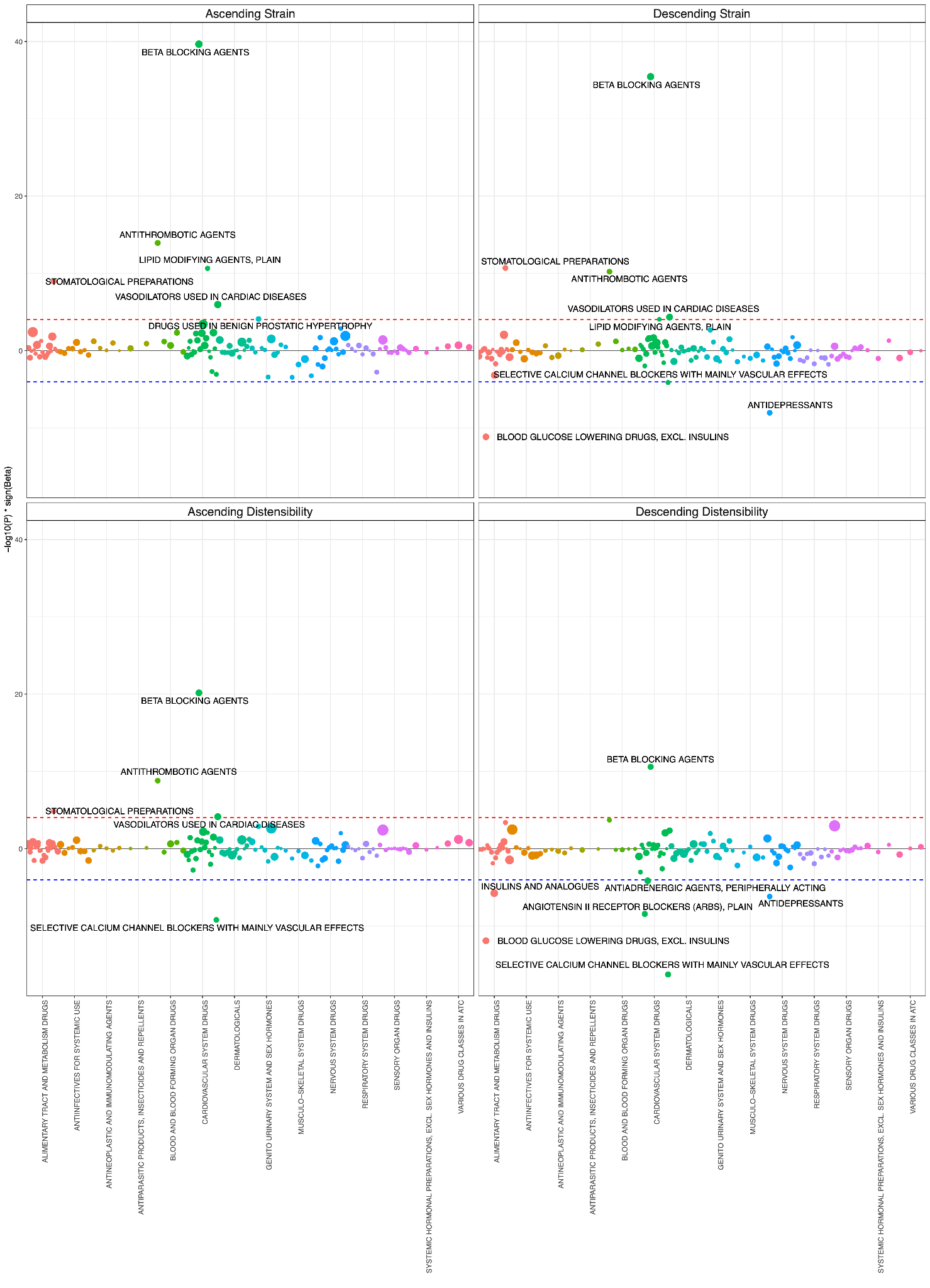

##### Supplementary Figure 5: PheCodes prior to adjusting for medications

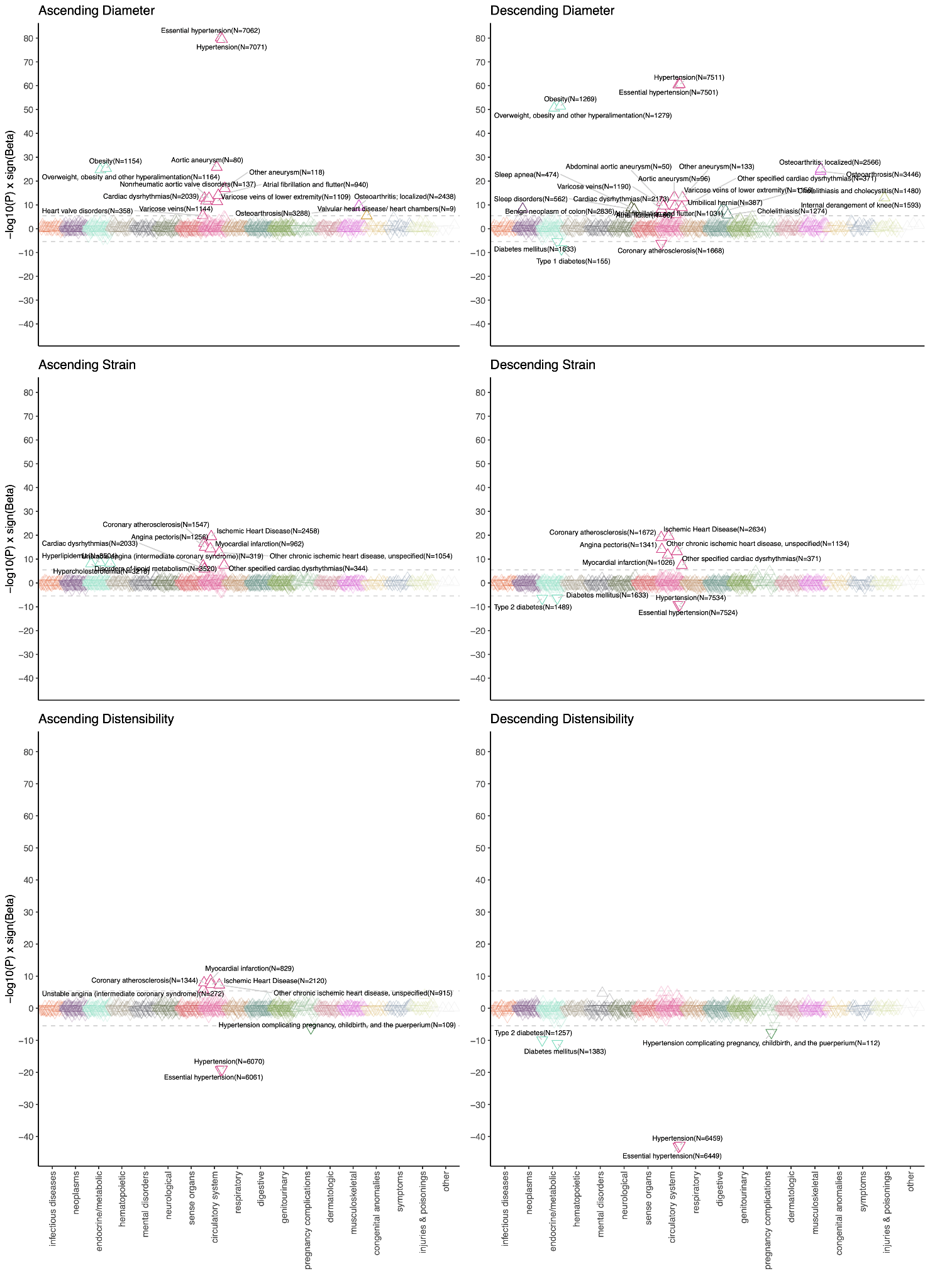

##### Supplementary Figure 6: PheCodes after adjusting for medications

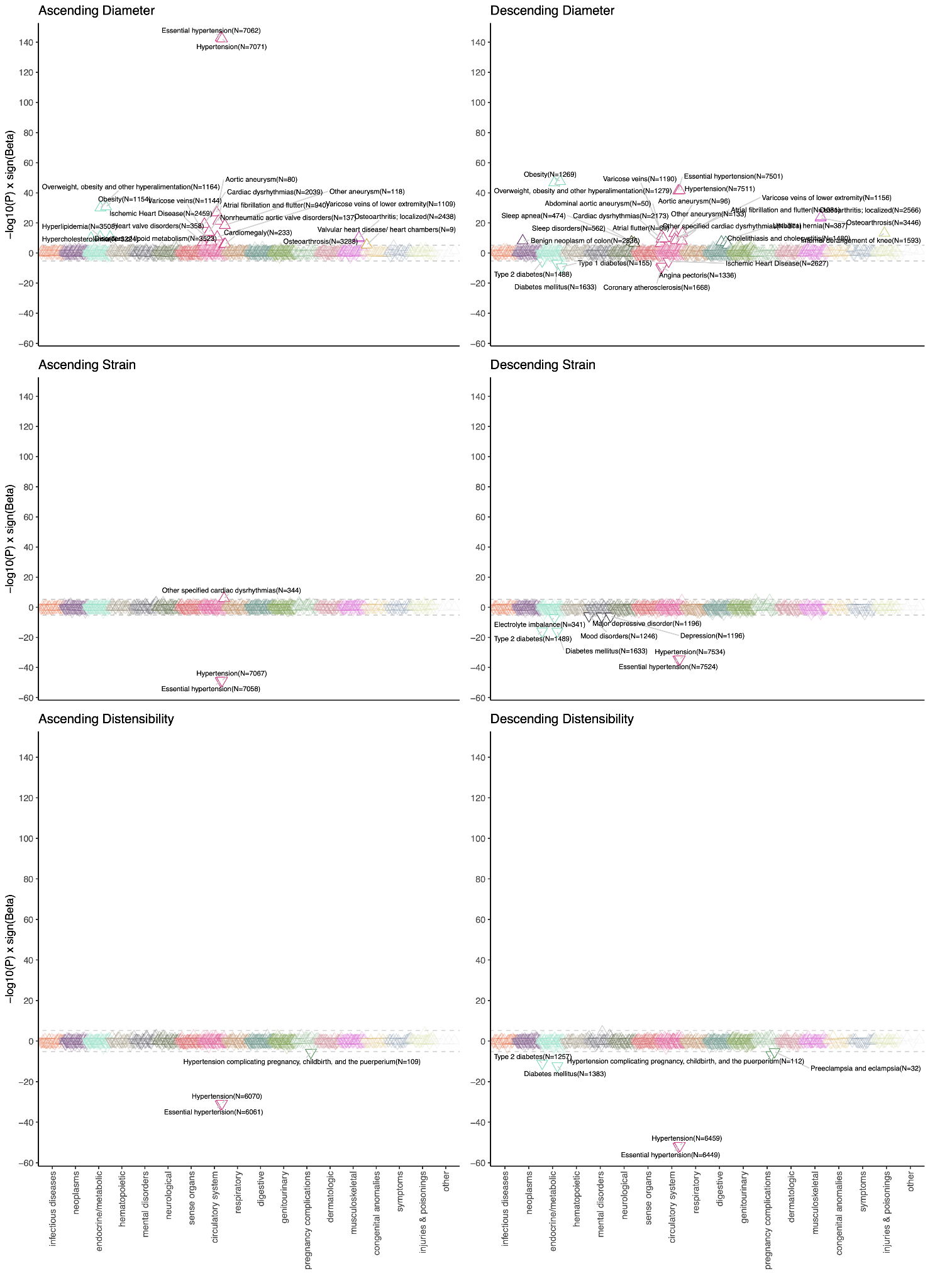

##### Supplementary Figure 7: Cross-trait genetic correlation

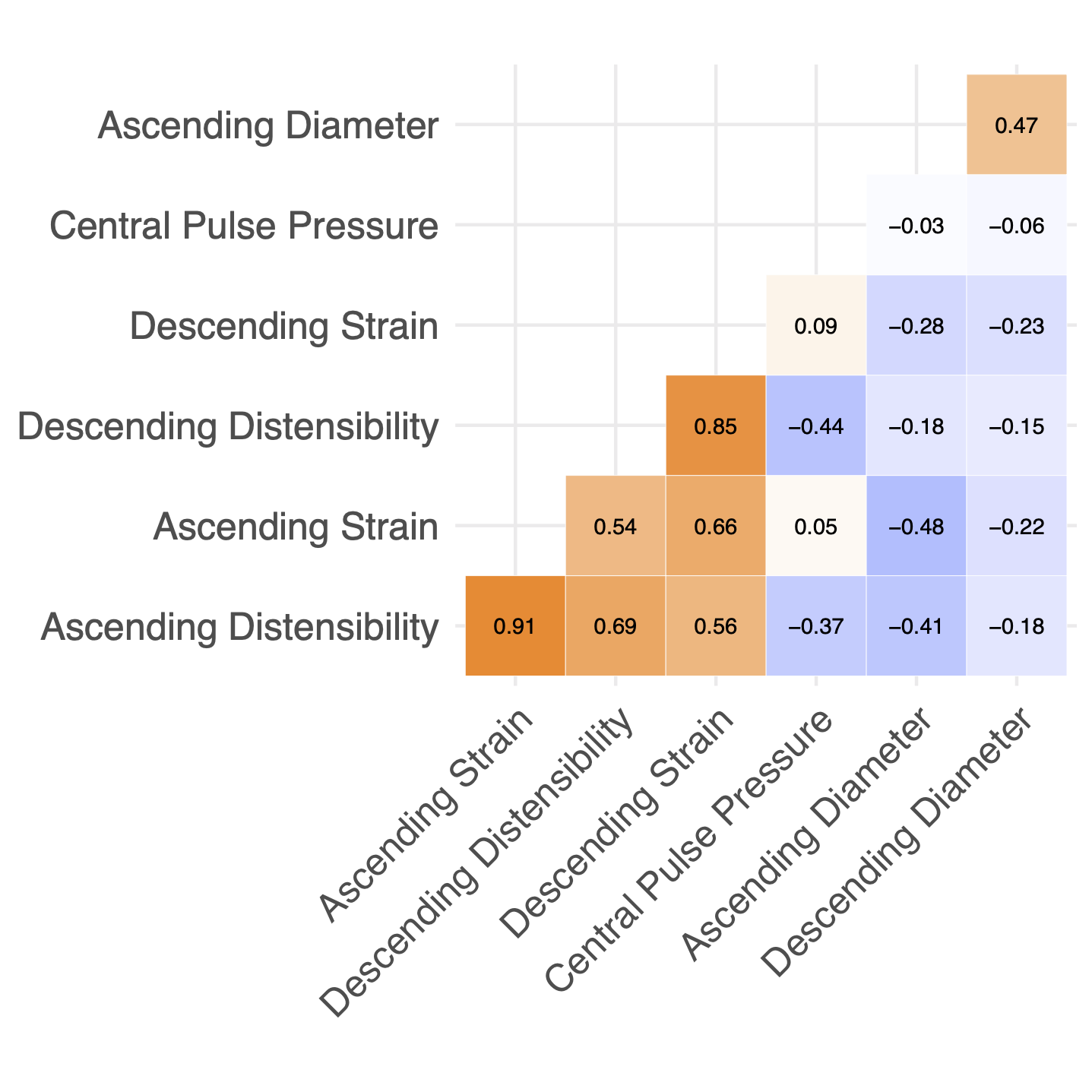

##### Supplementary Figure 8: GWAS QQ plots

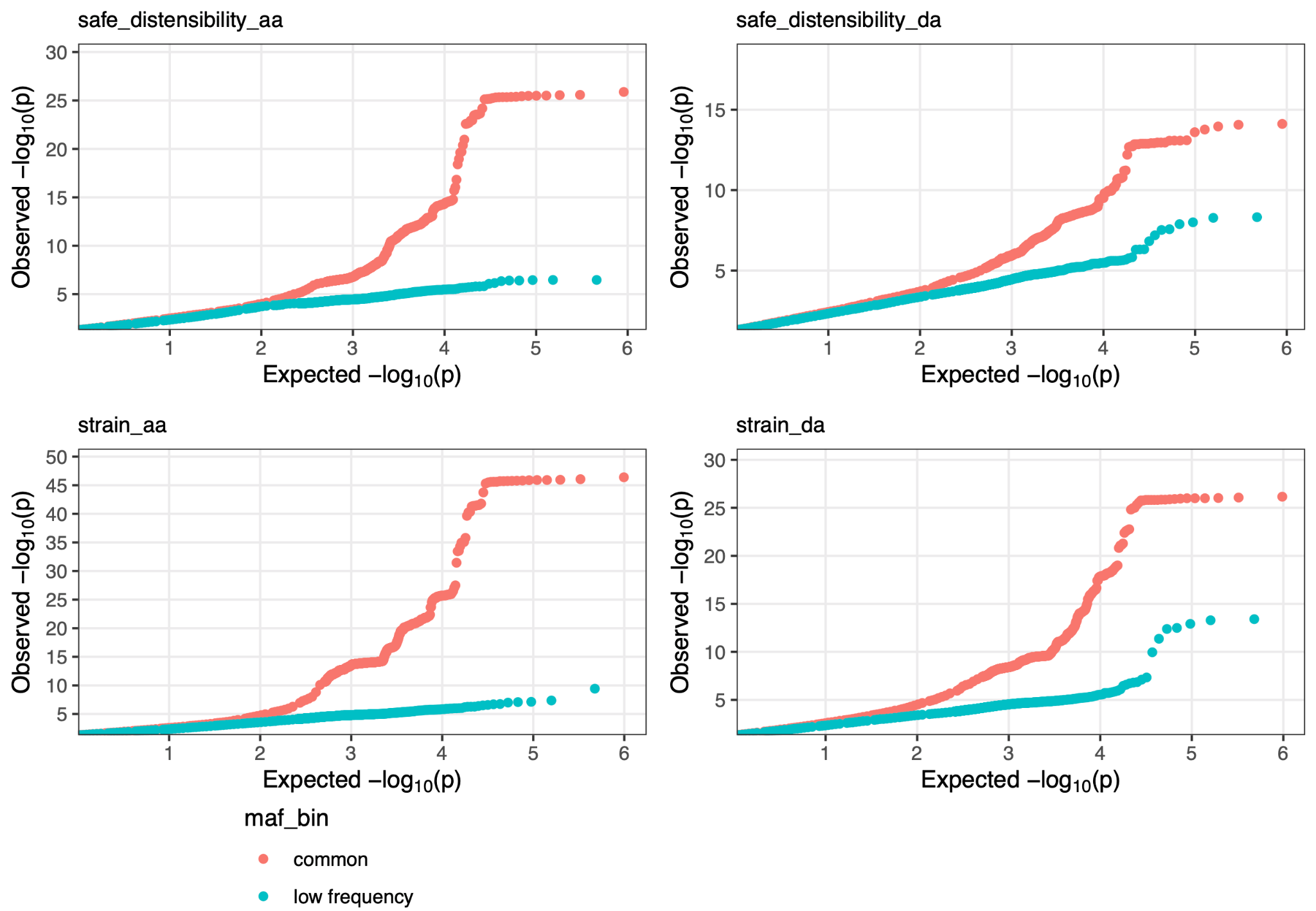

##### Supplementary Figure 9: GWAS locus plots

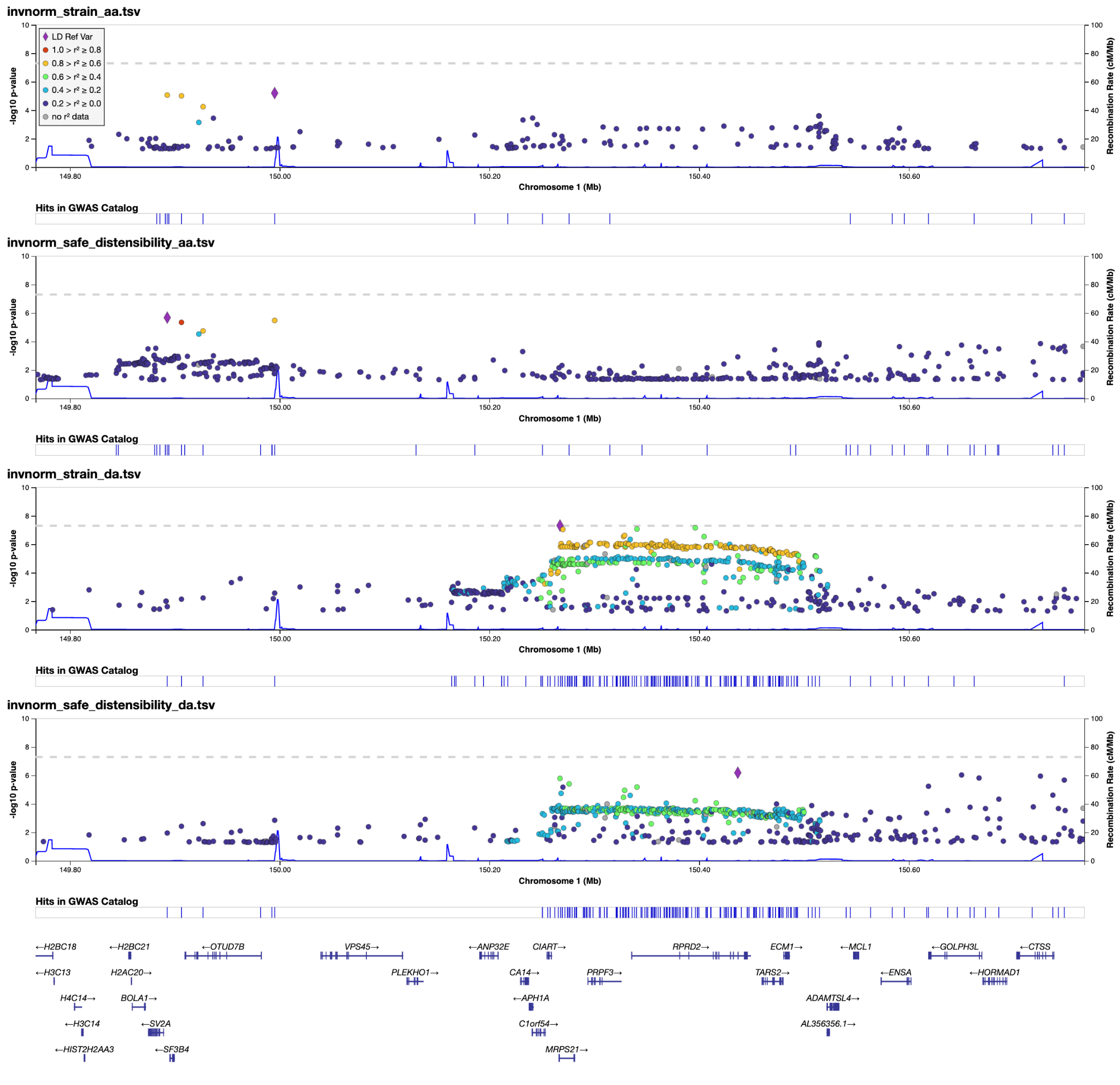

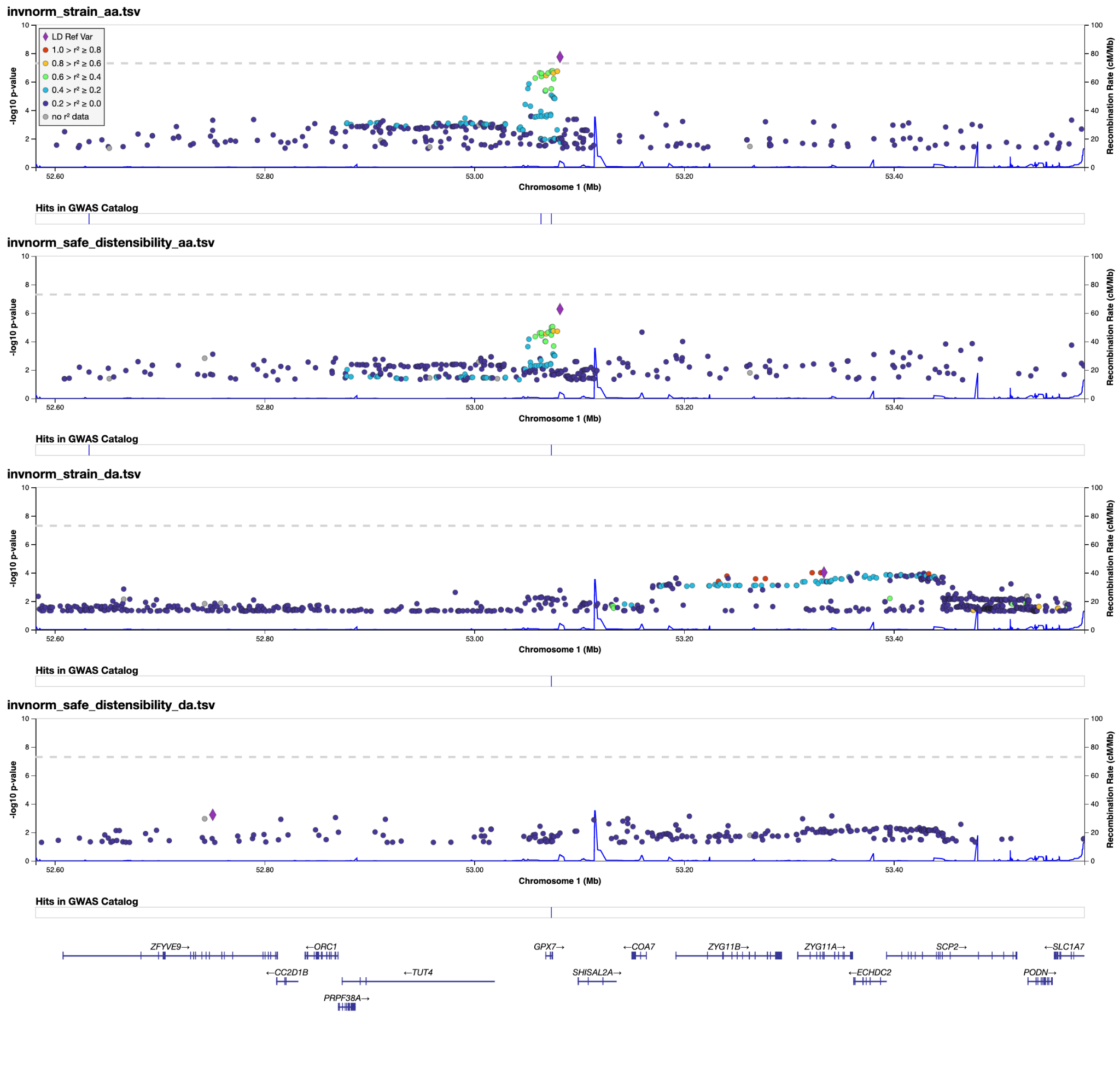

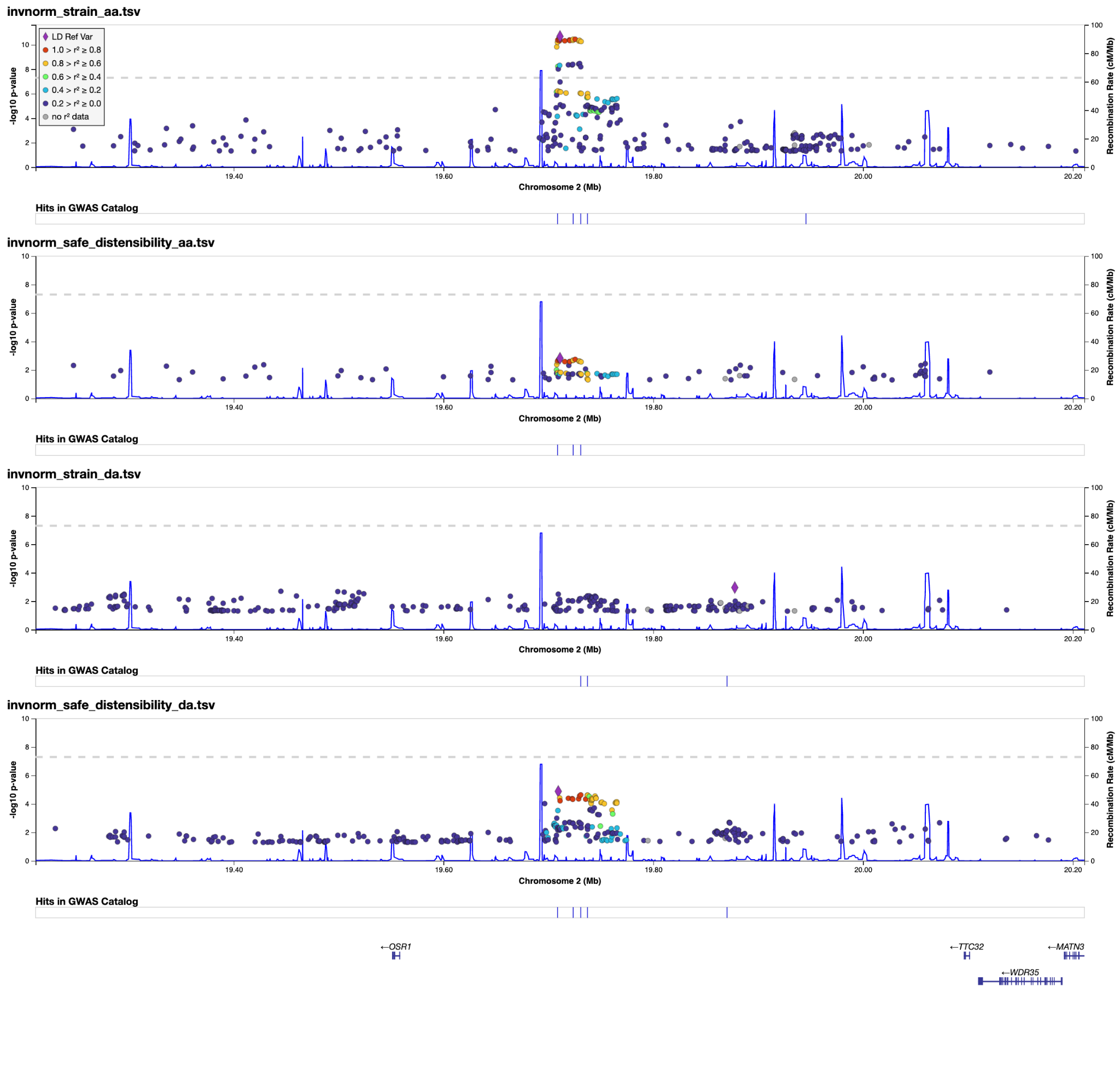

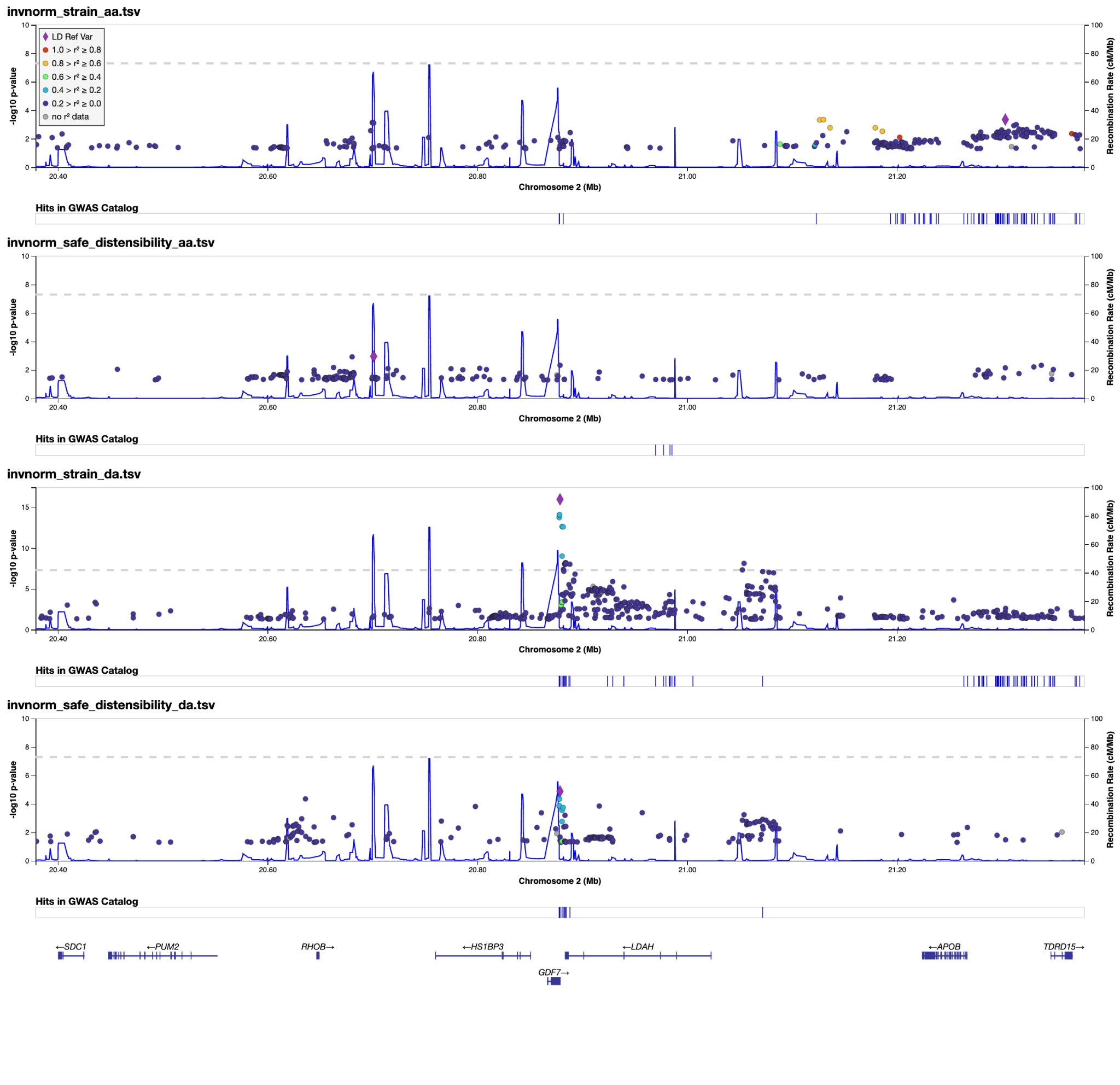

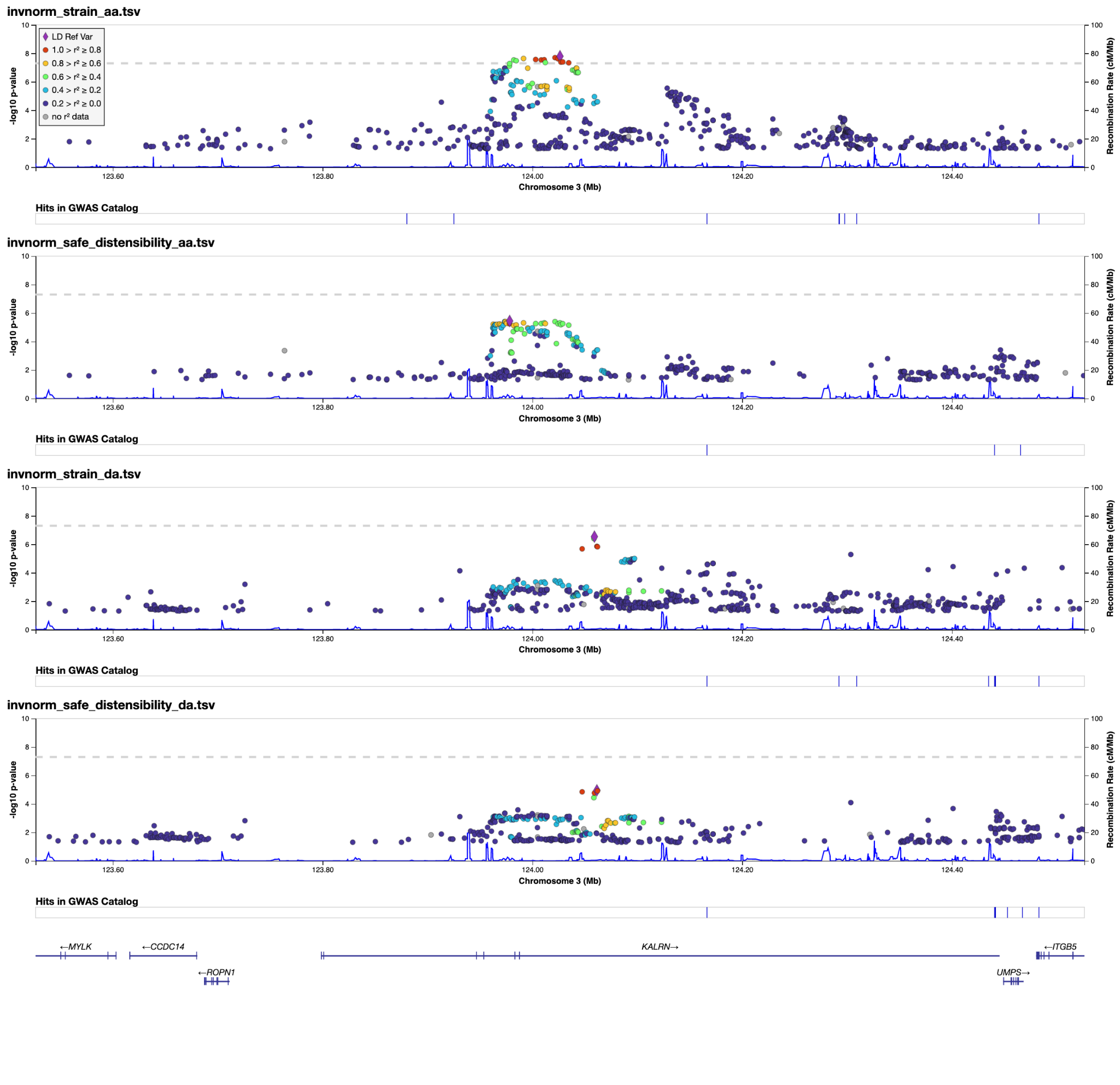

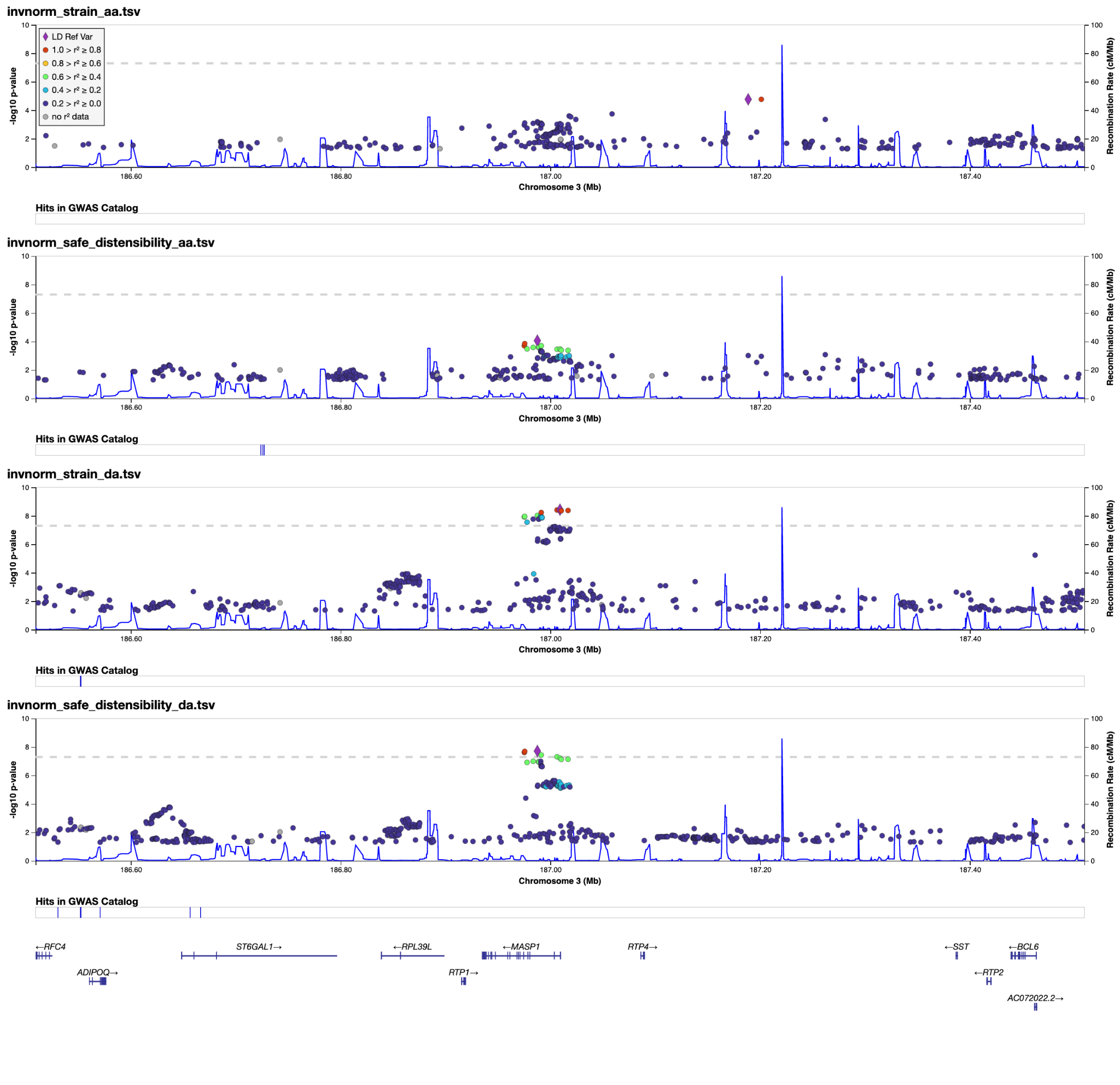

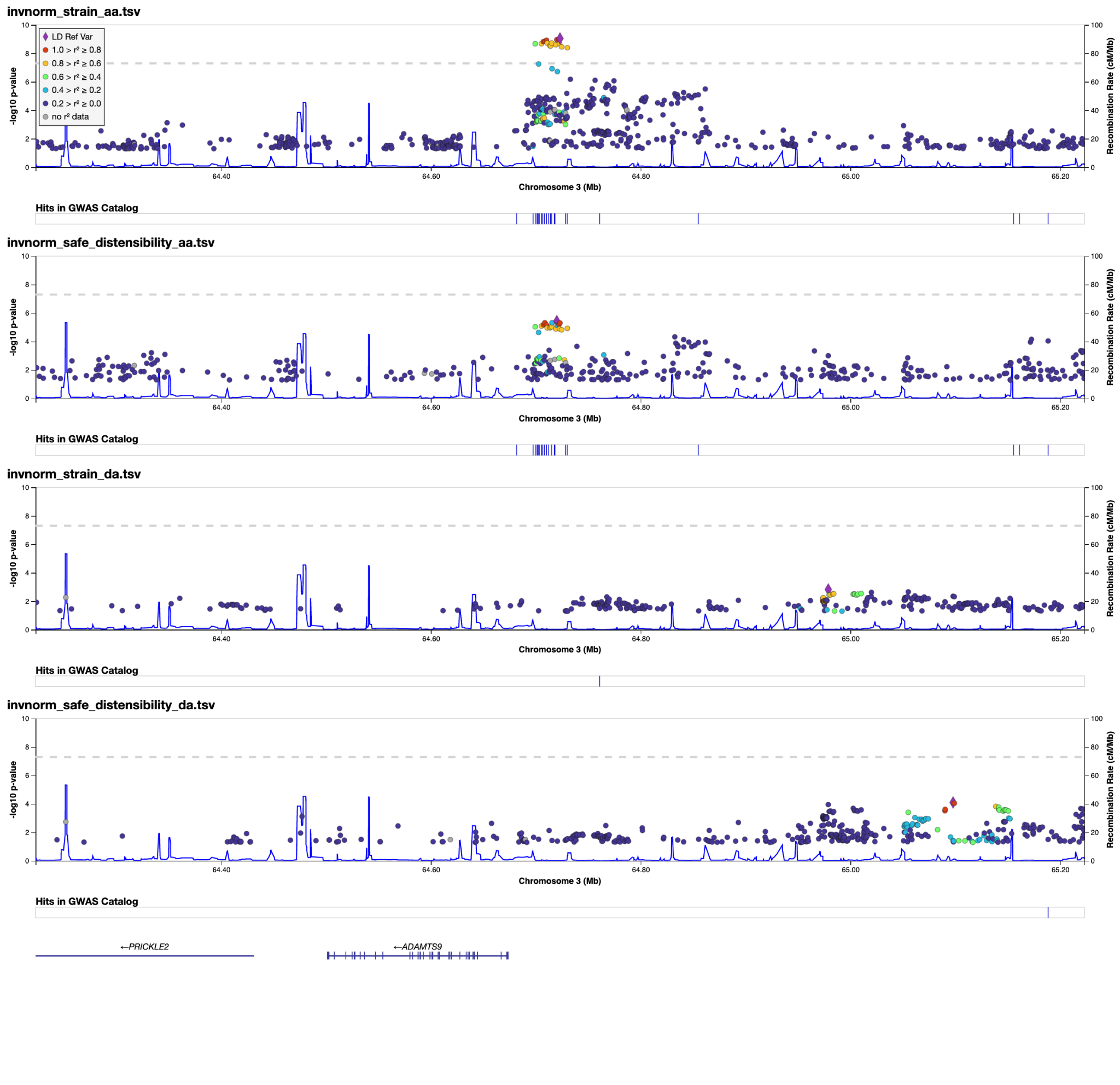

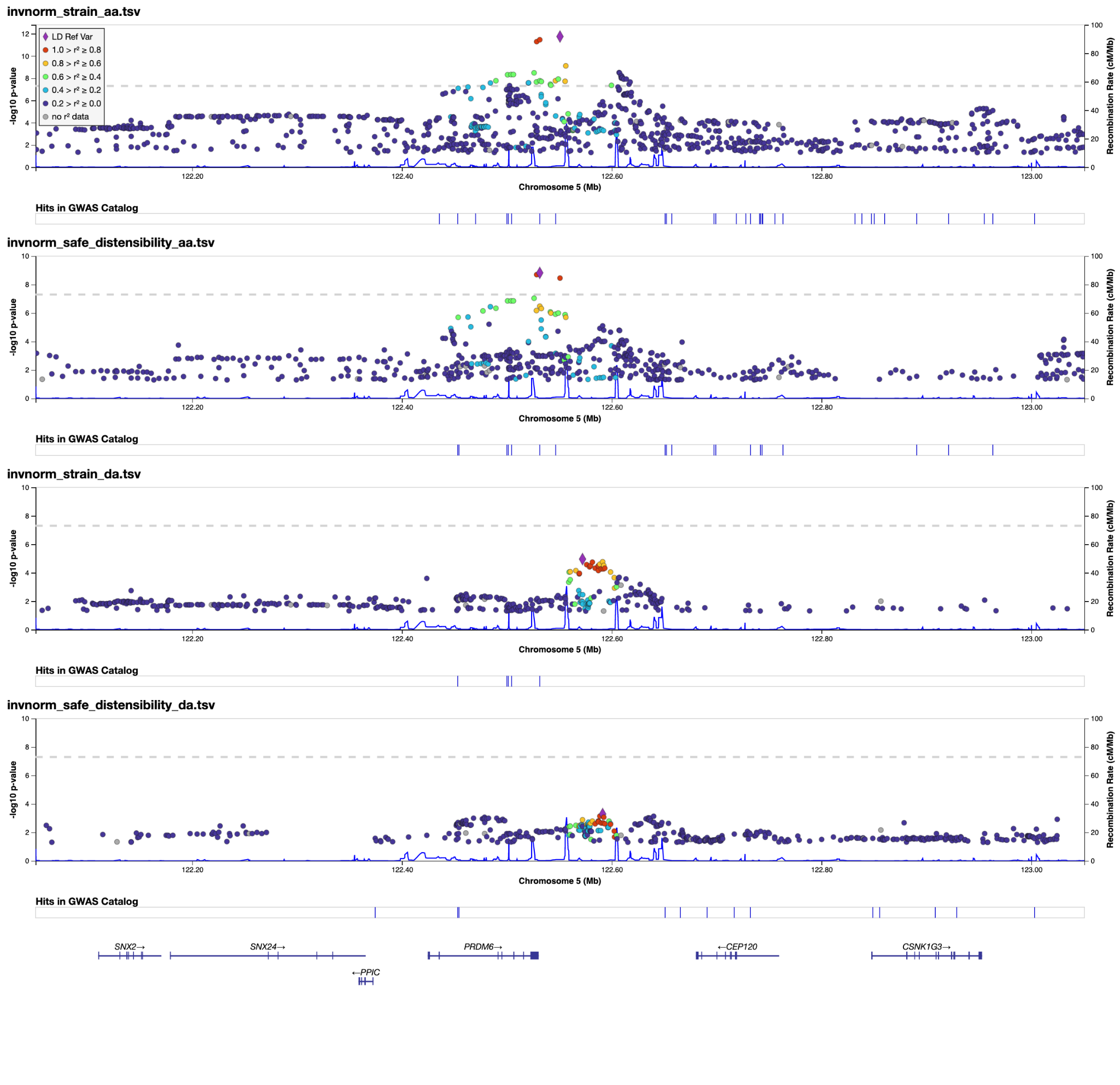

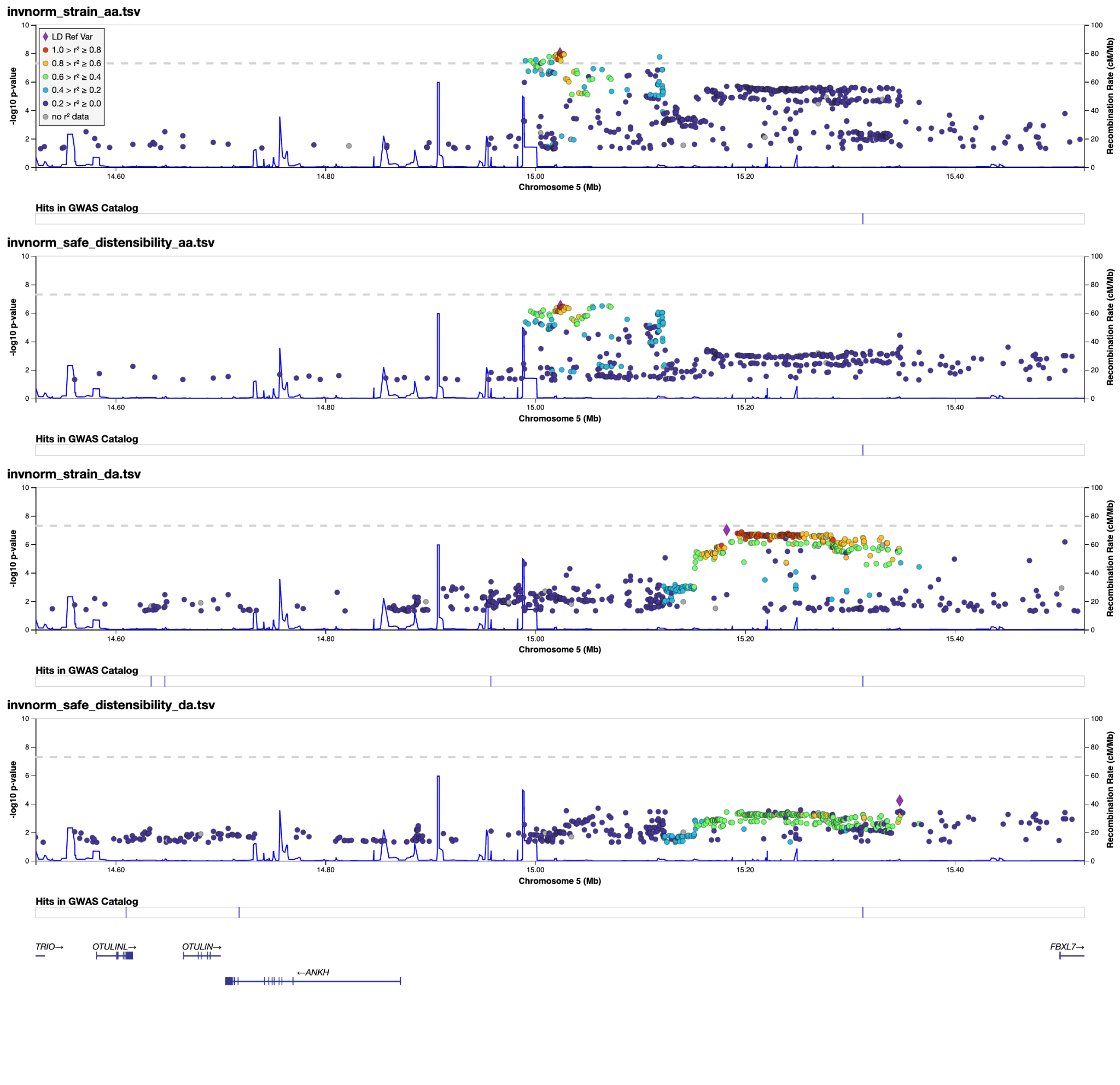

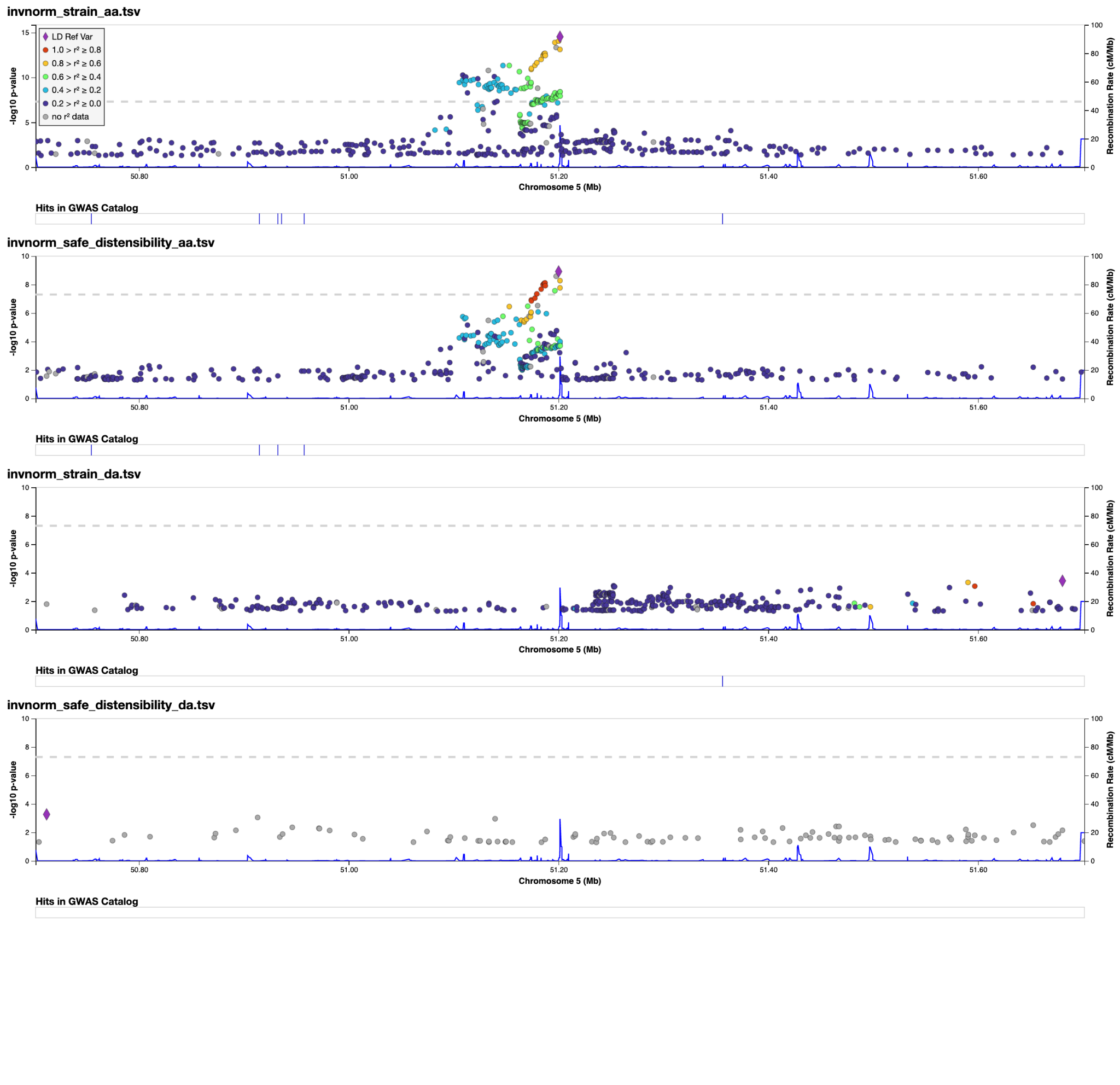

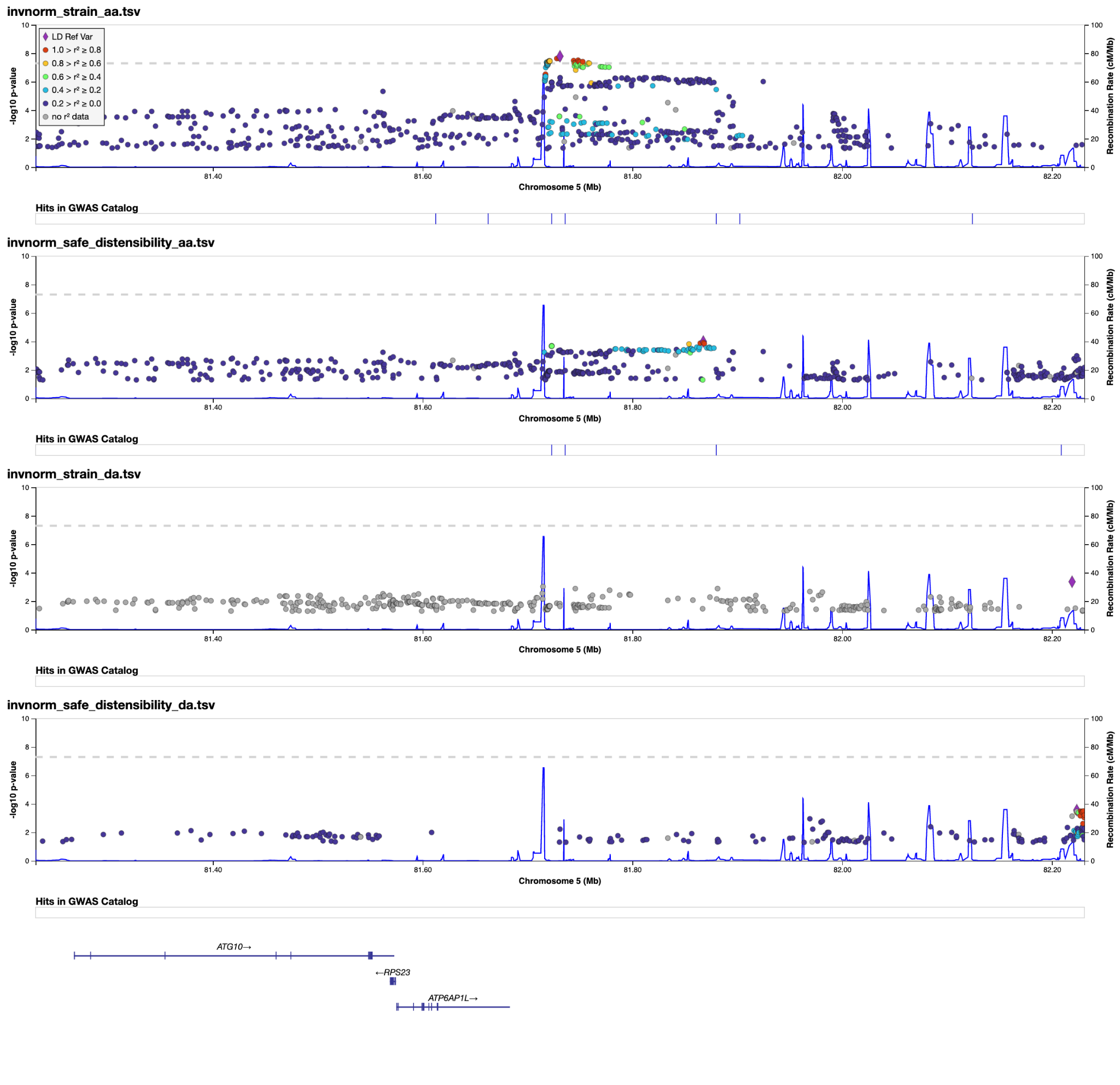

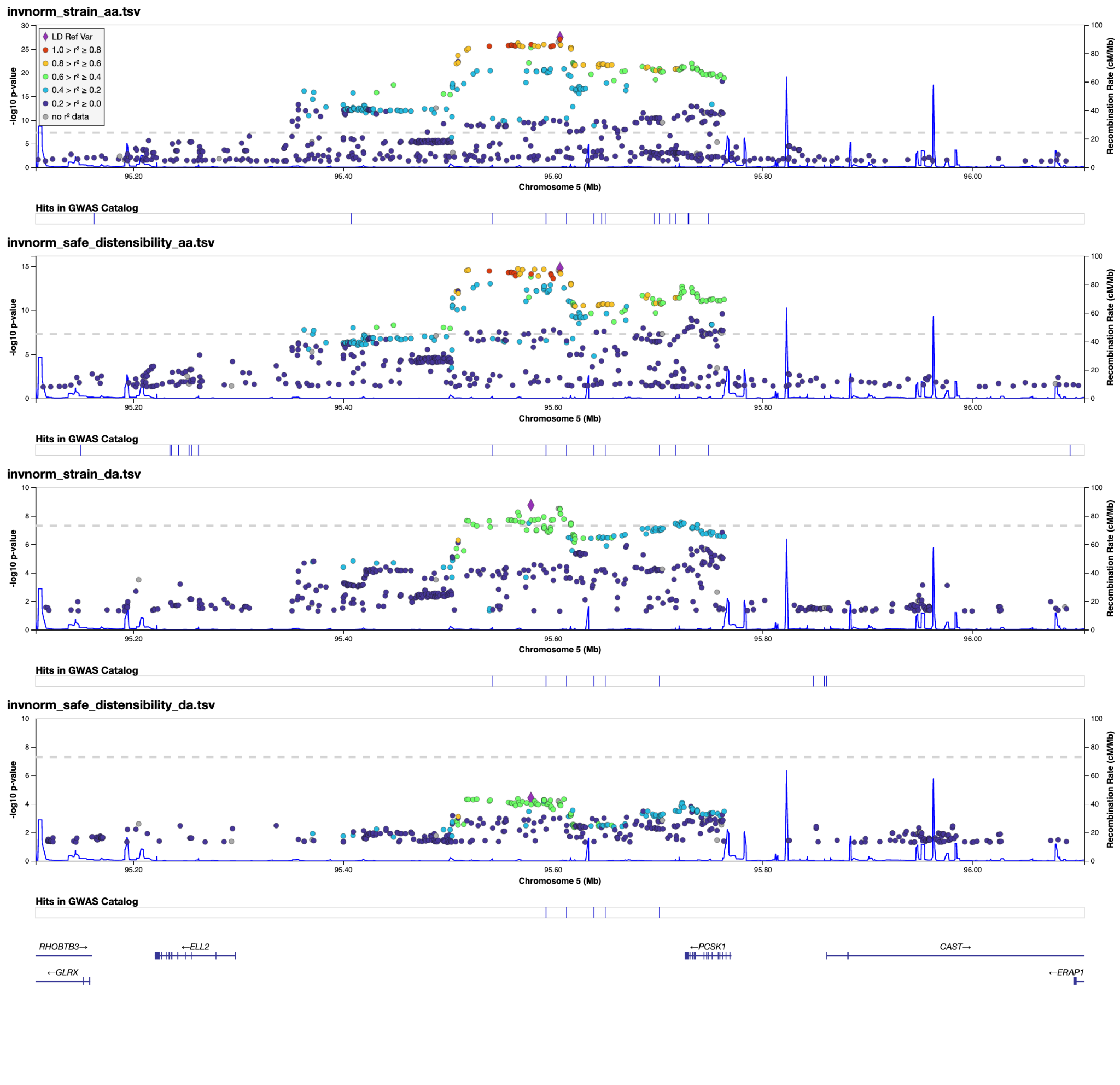

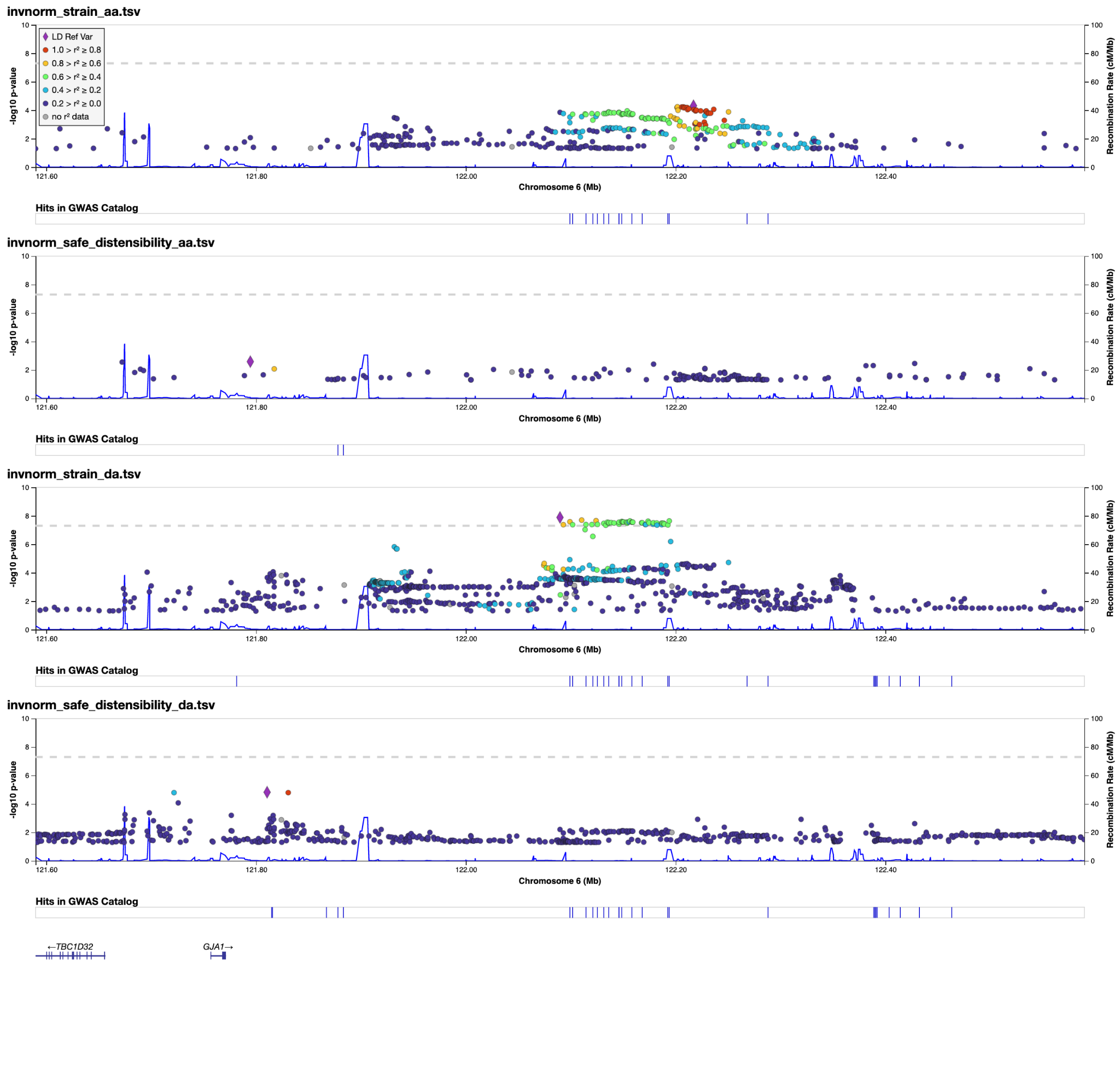

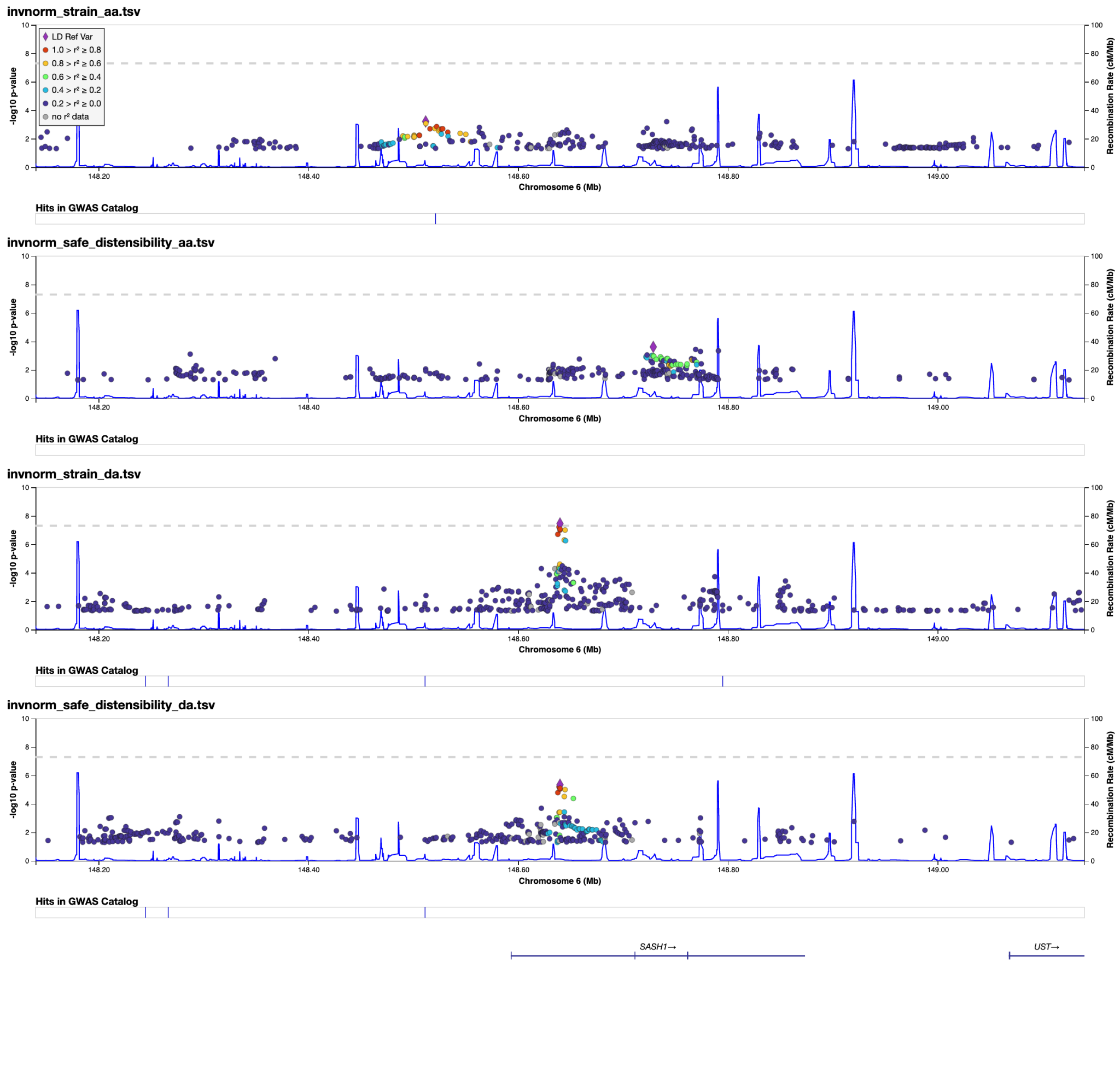

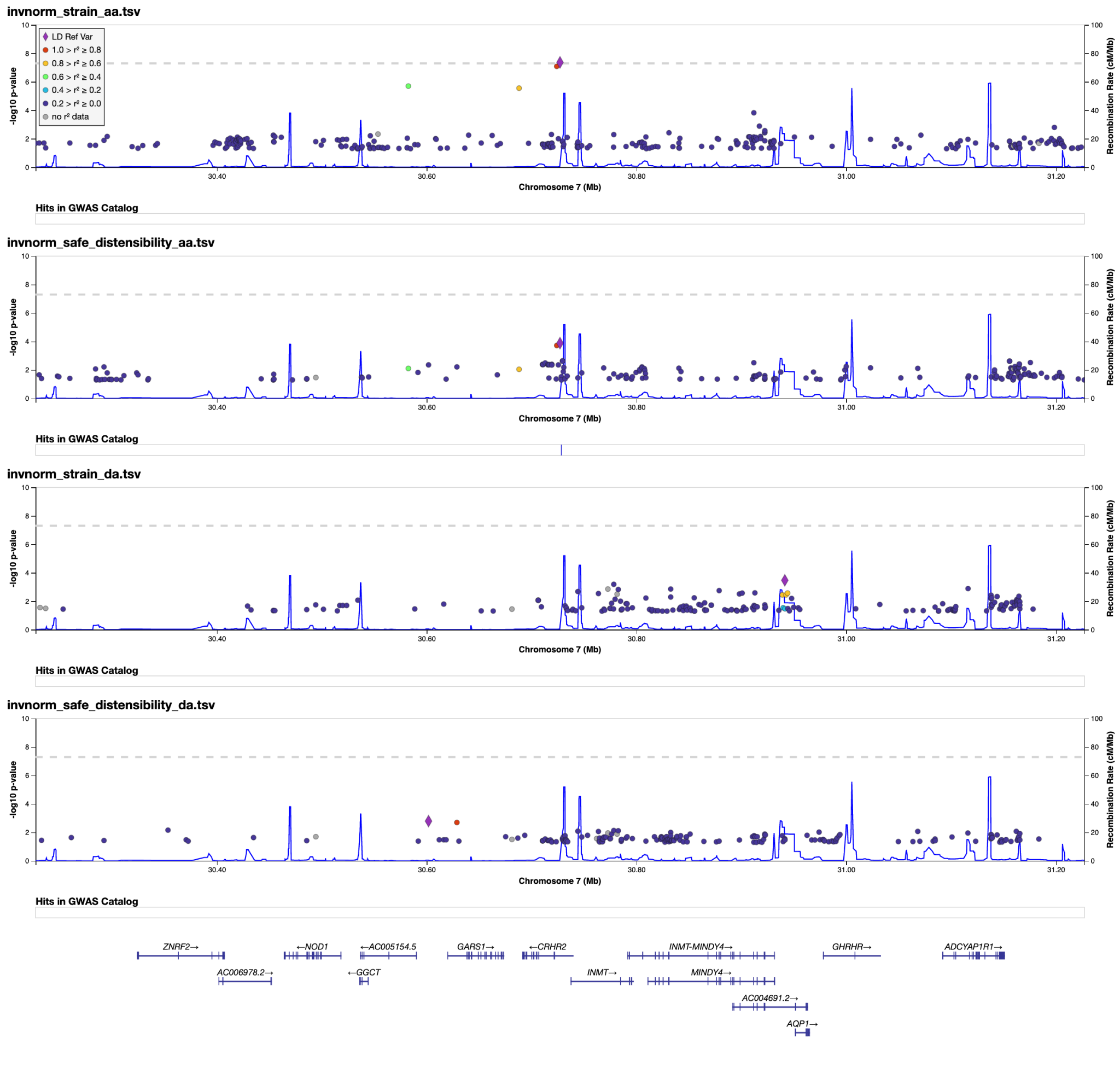

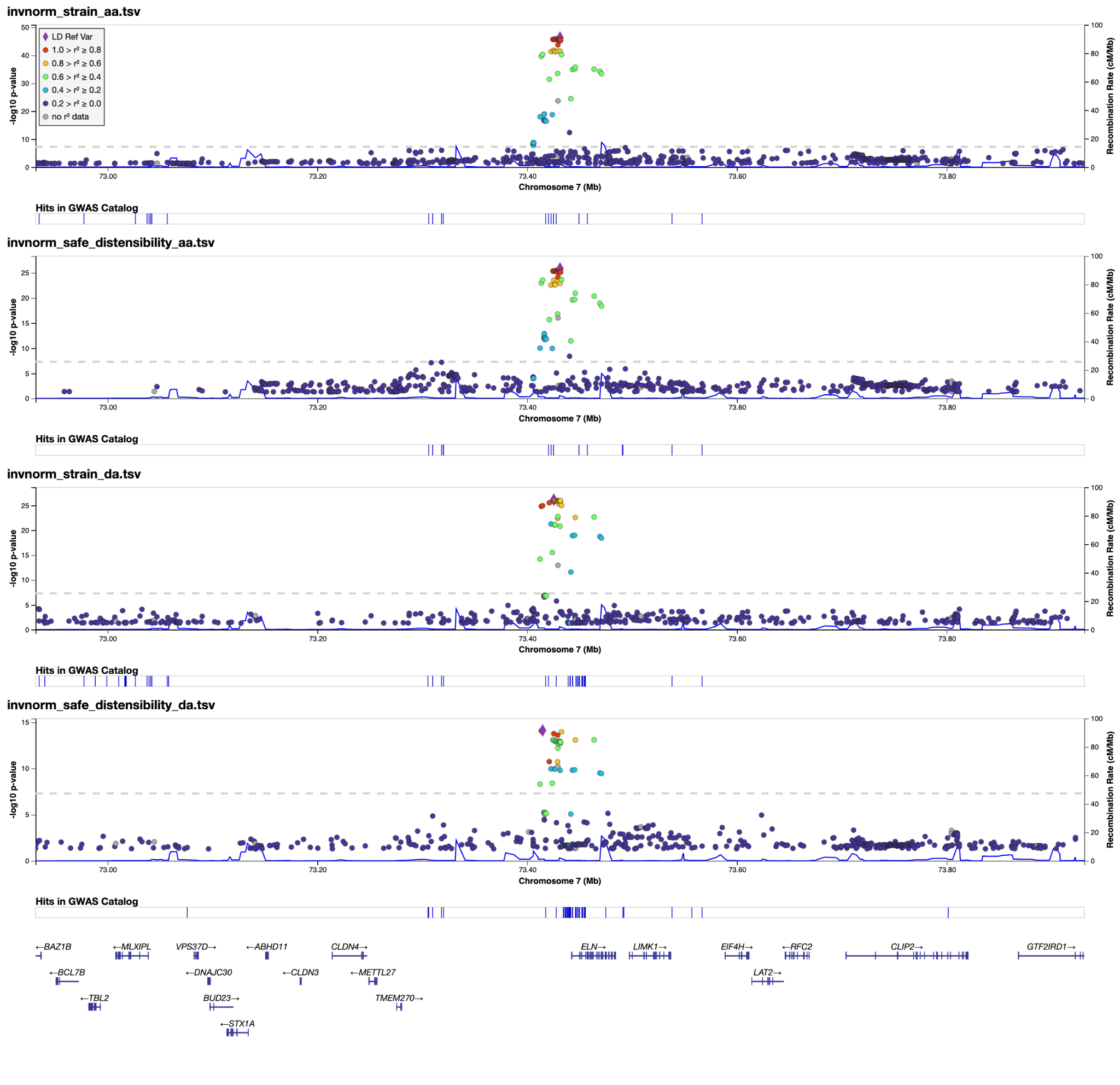

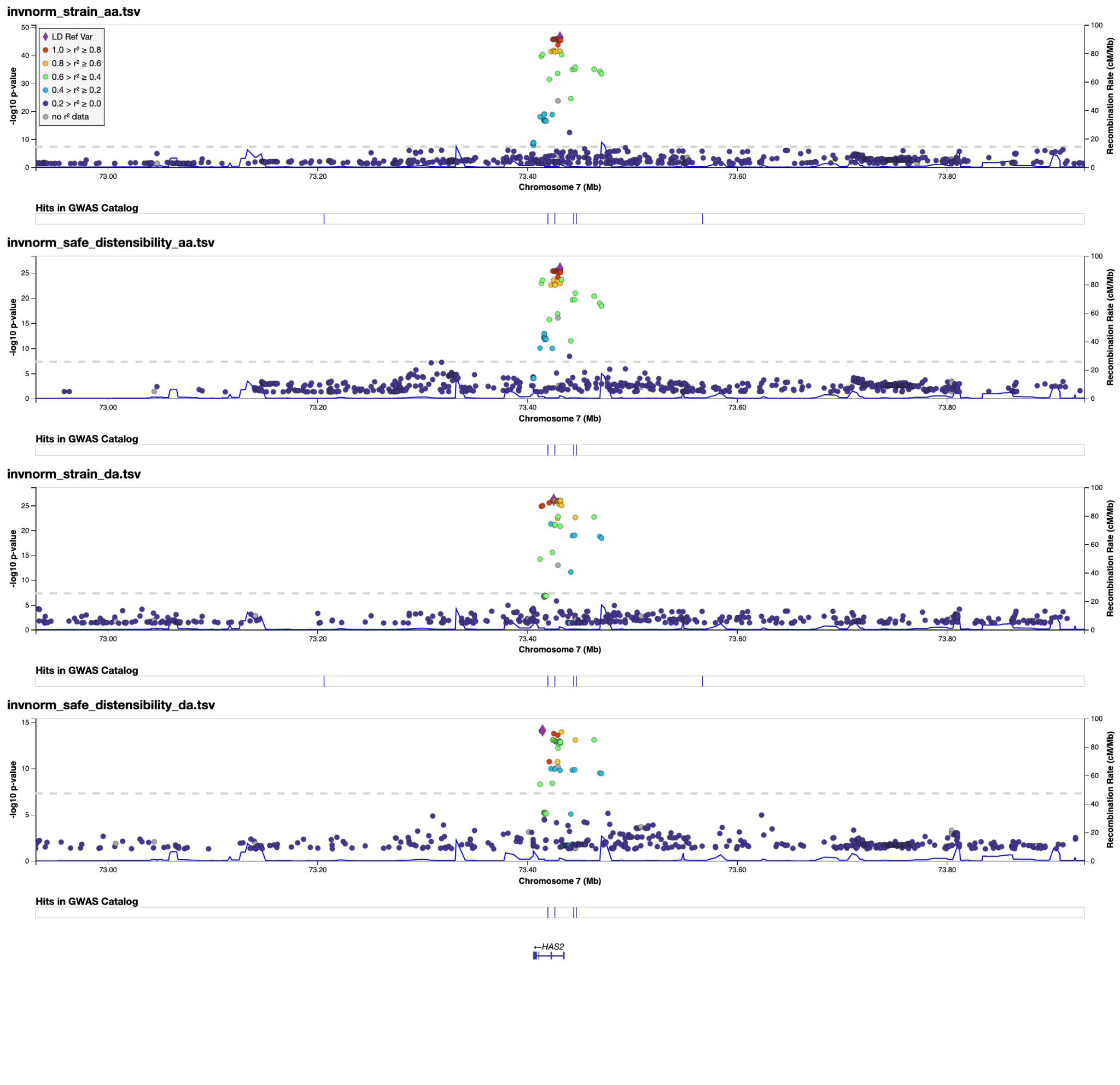

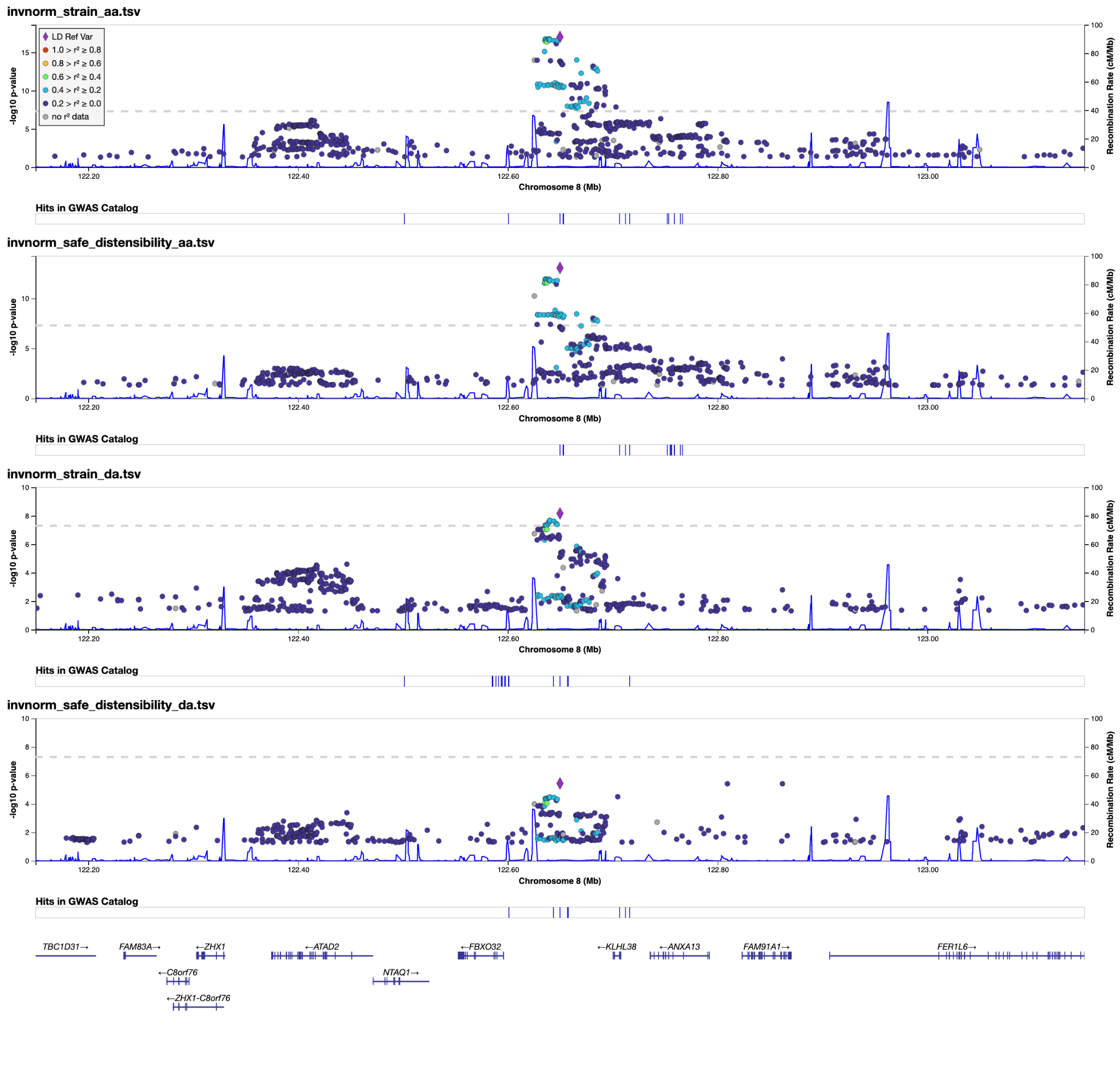

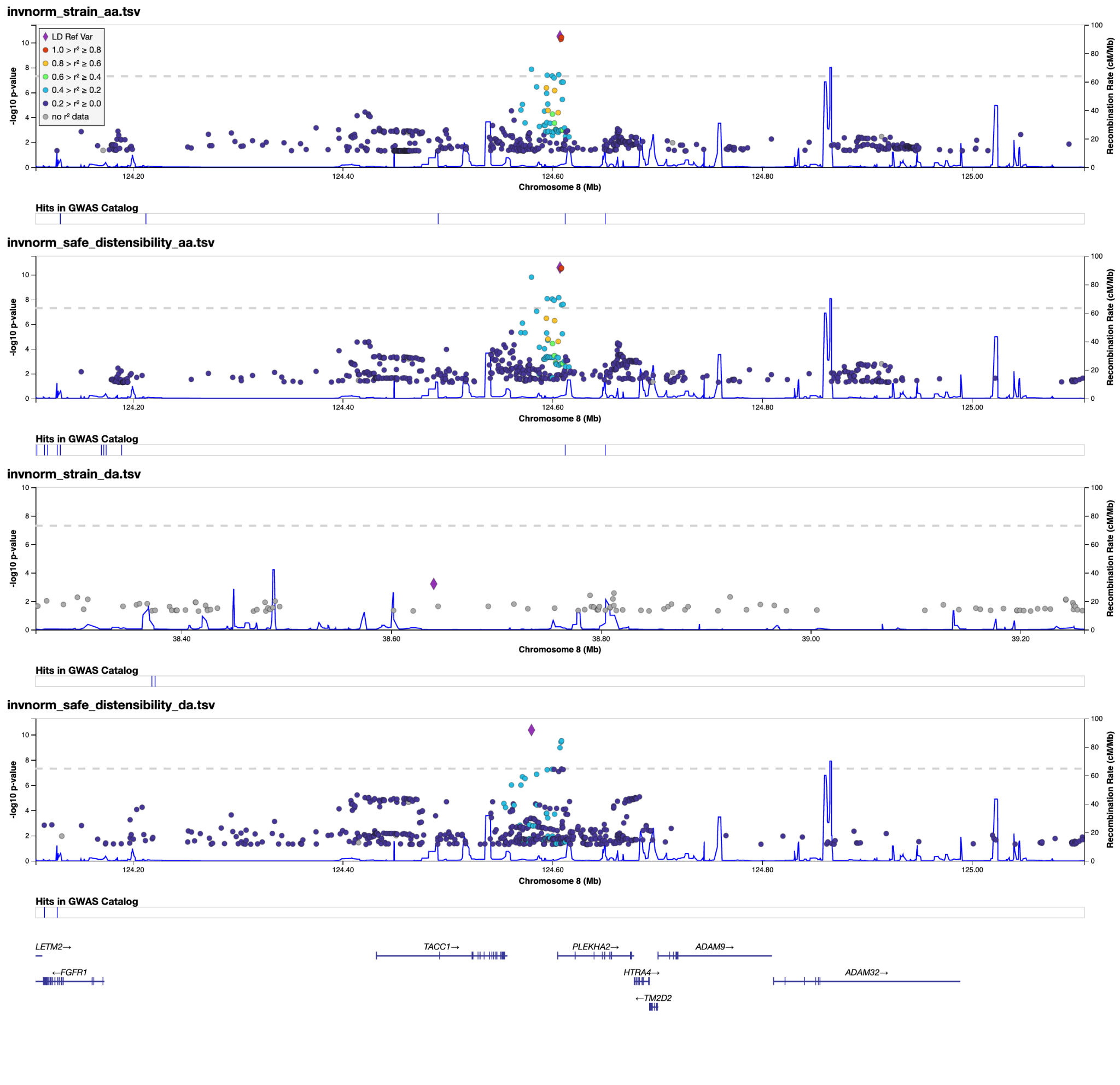

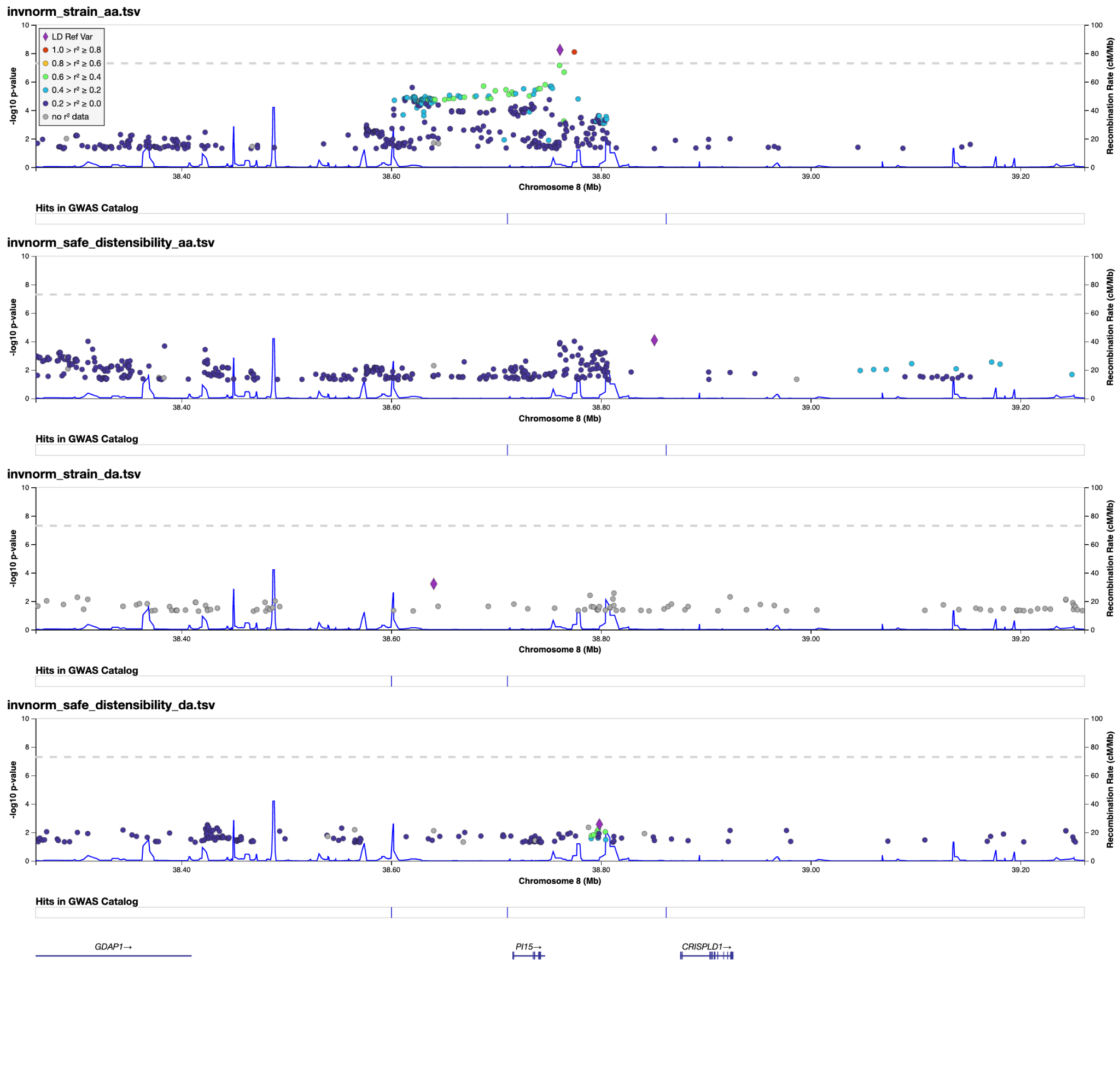

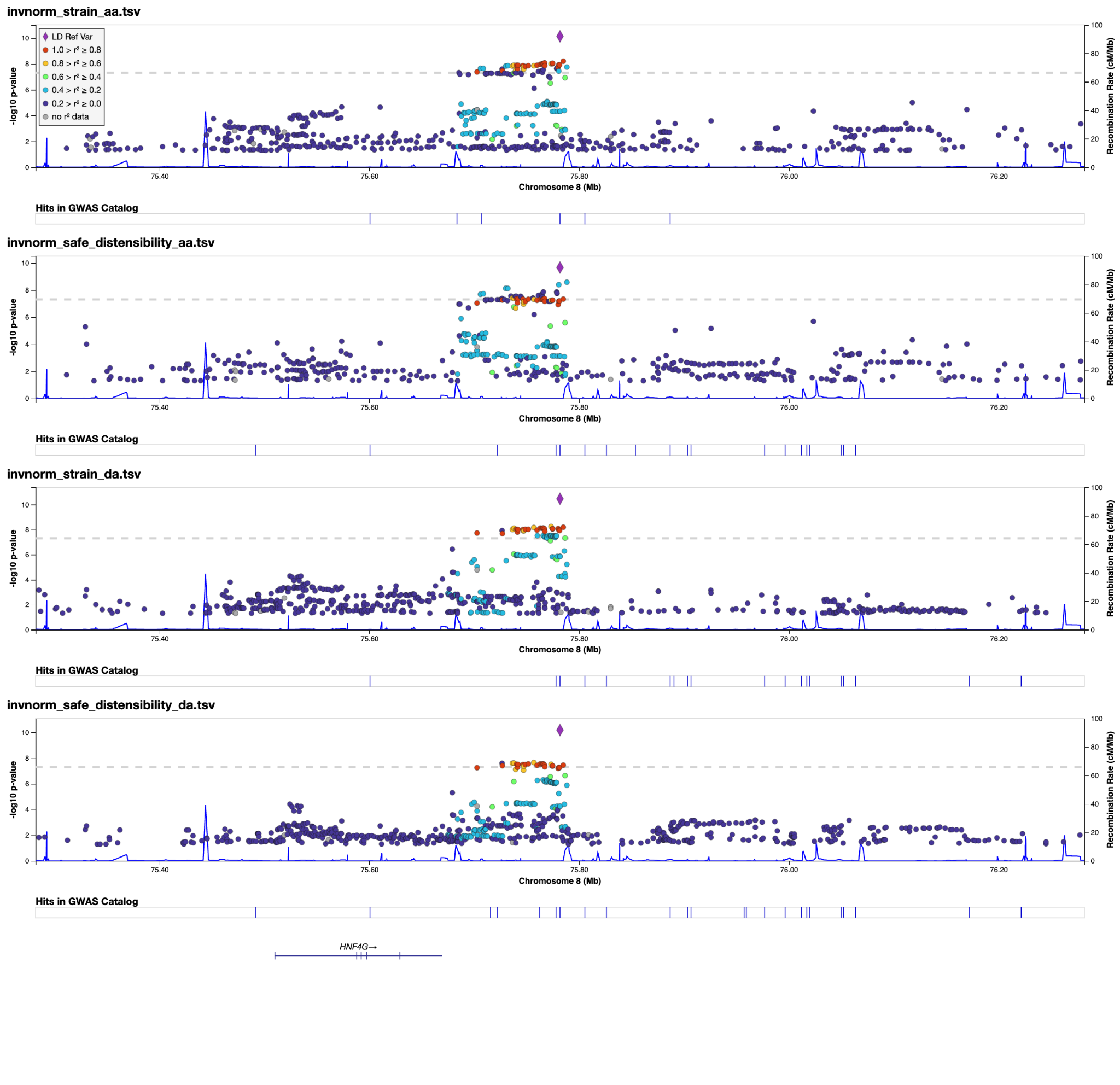

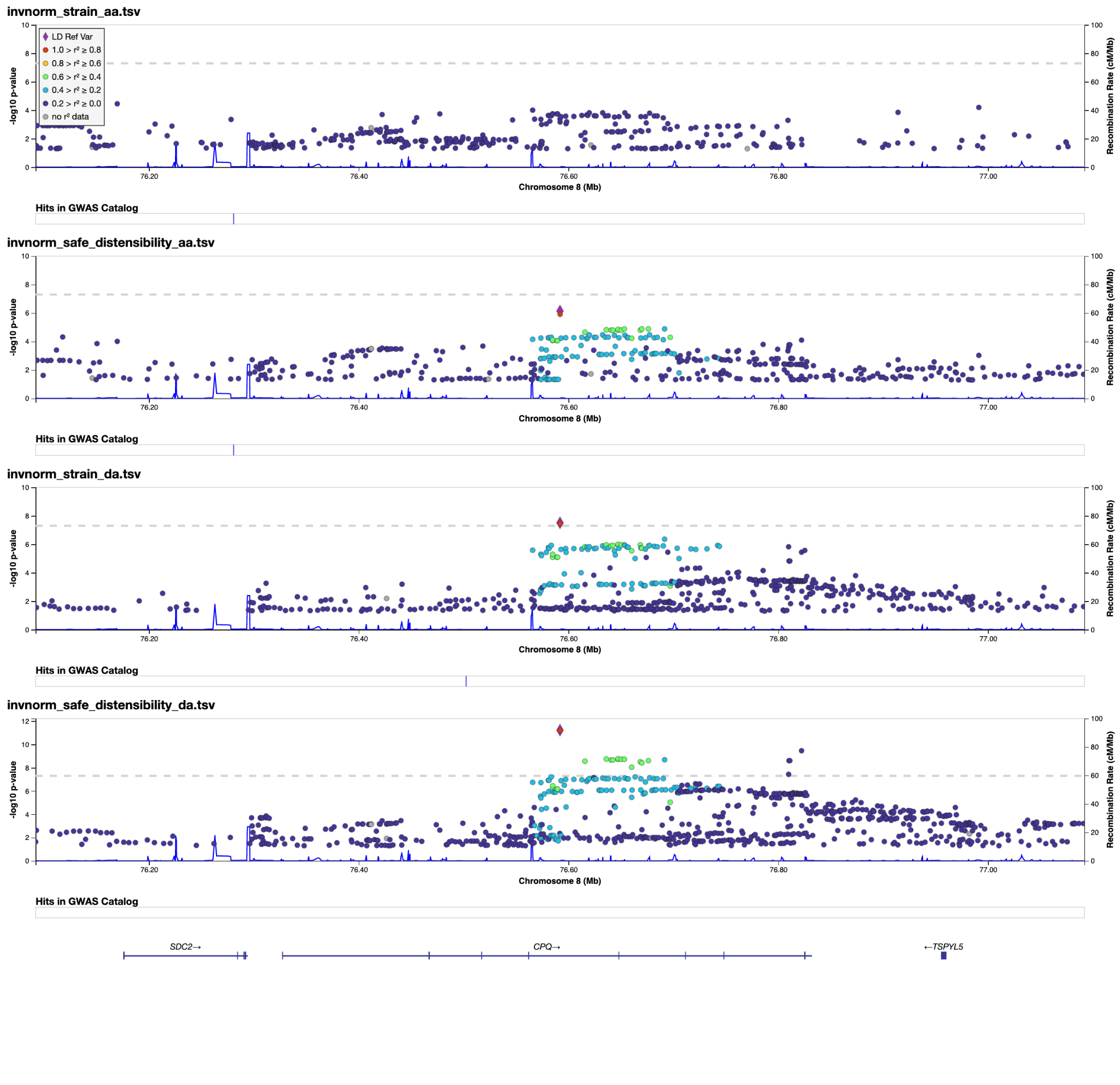

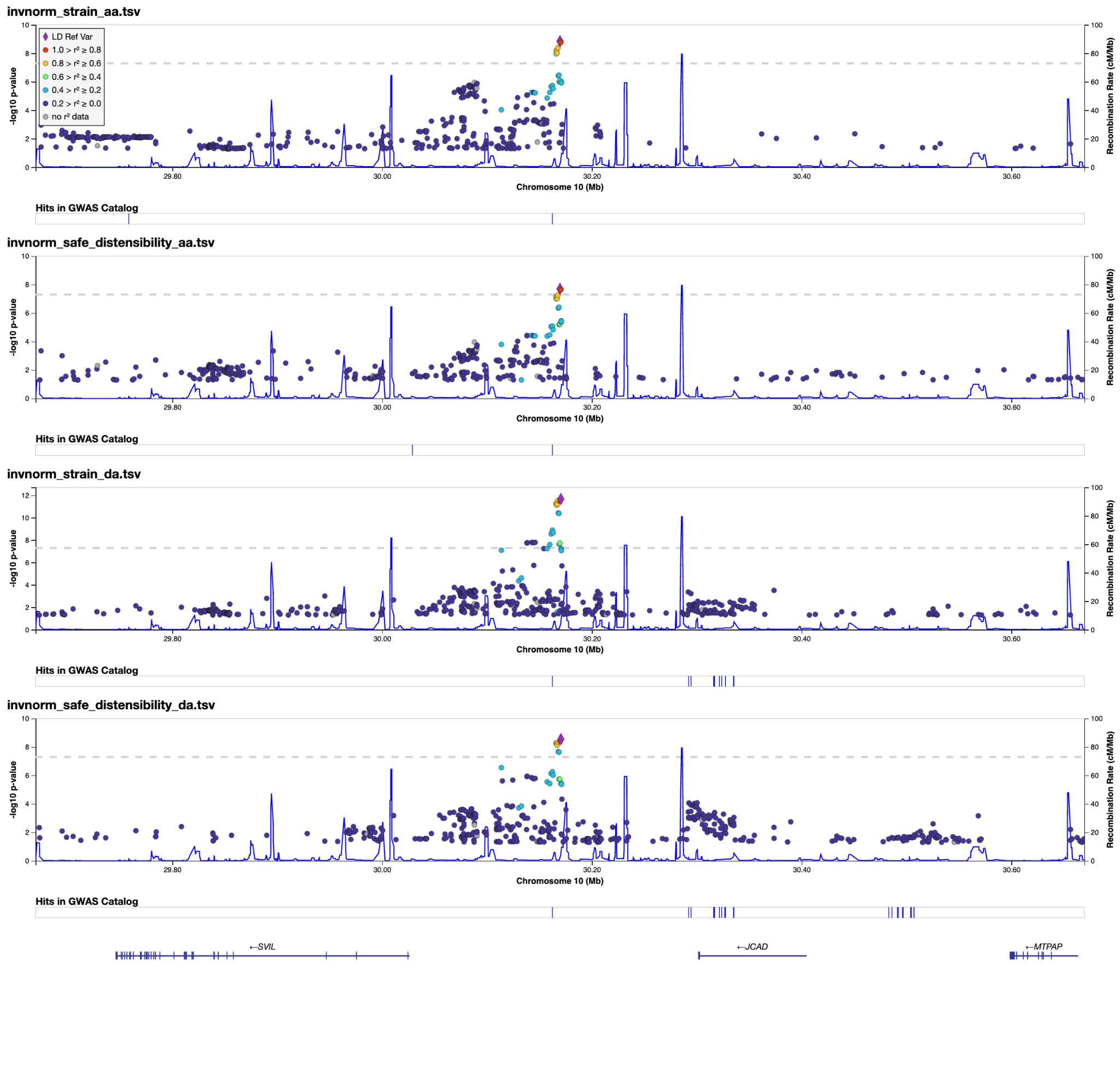

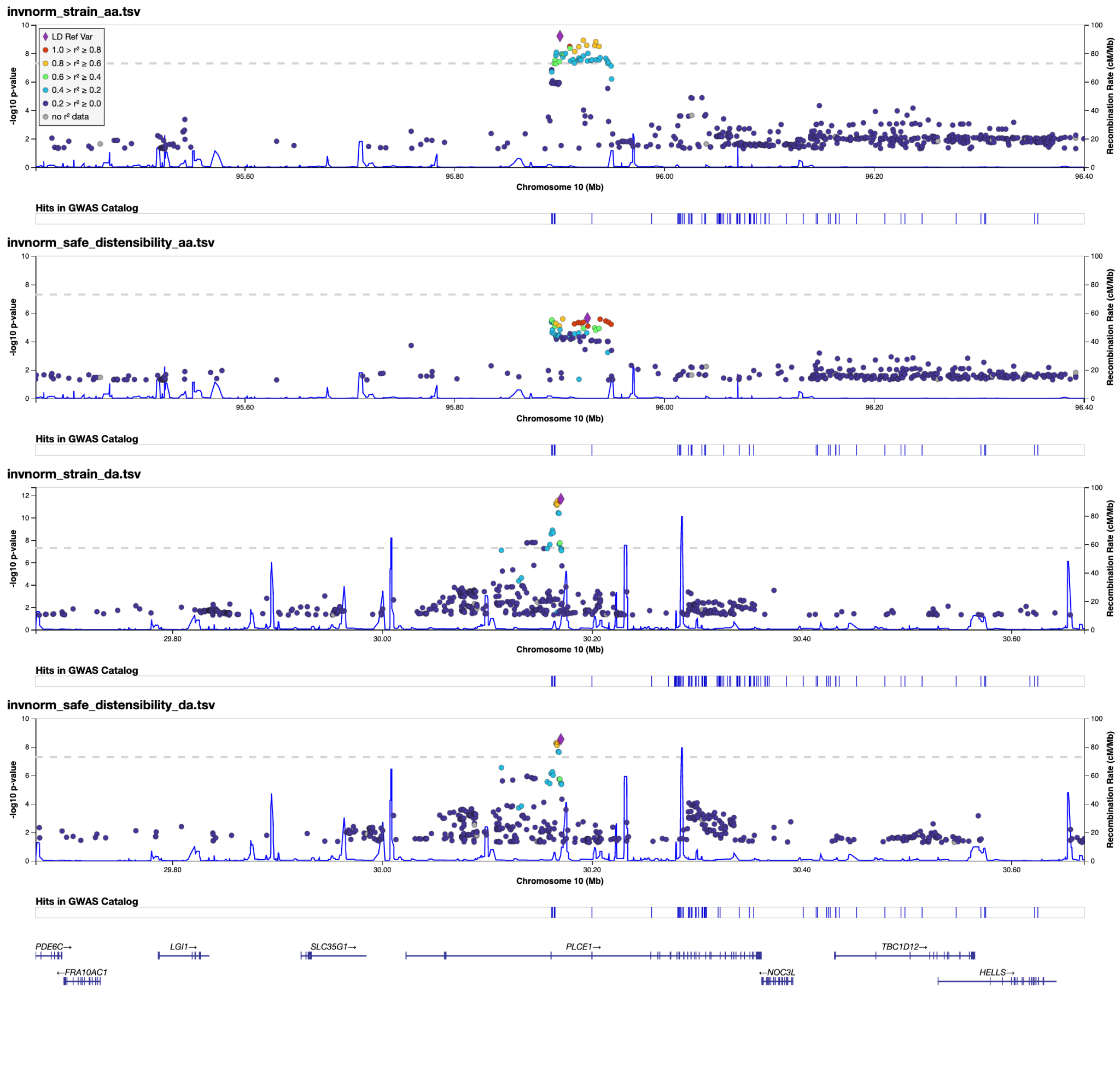

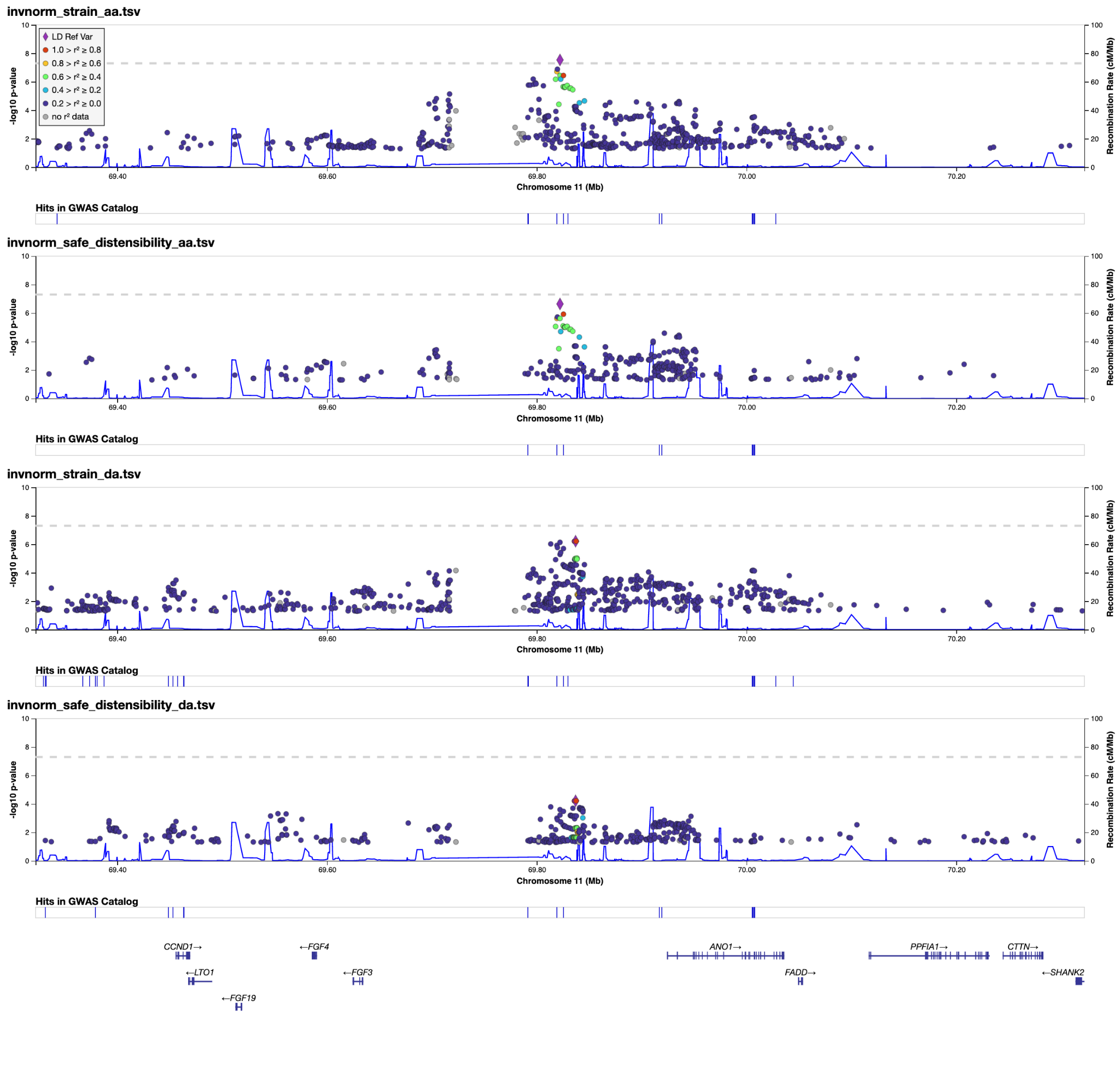

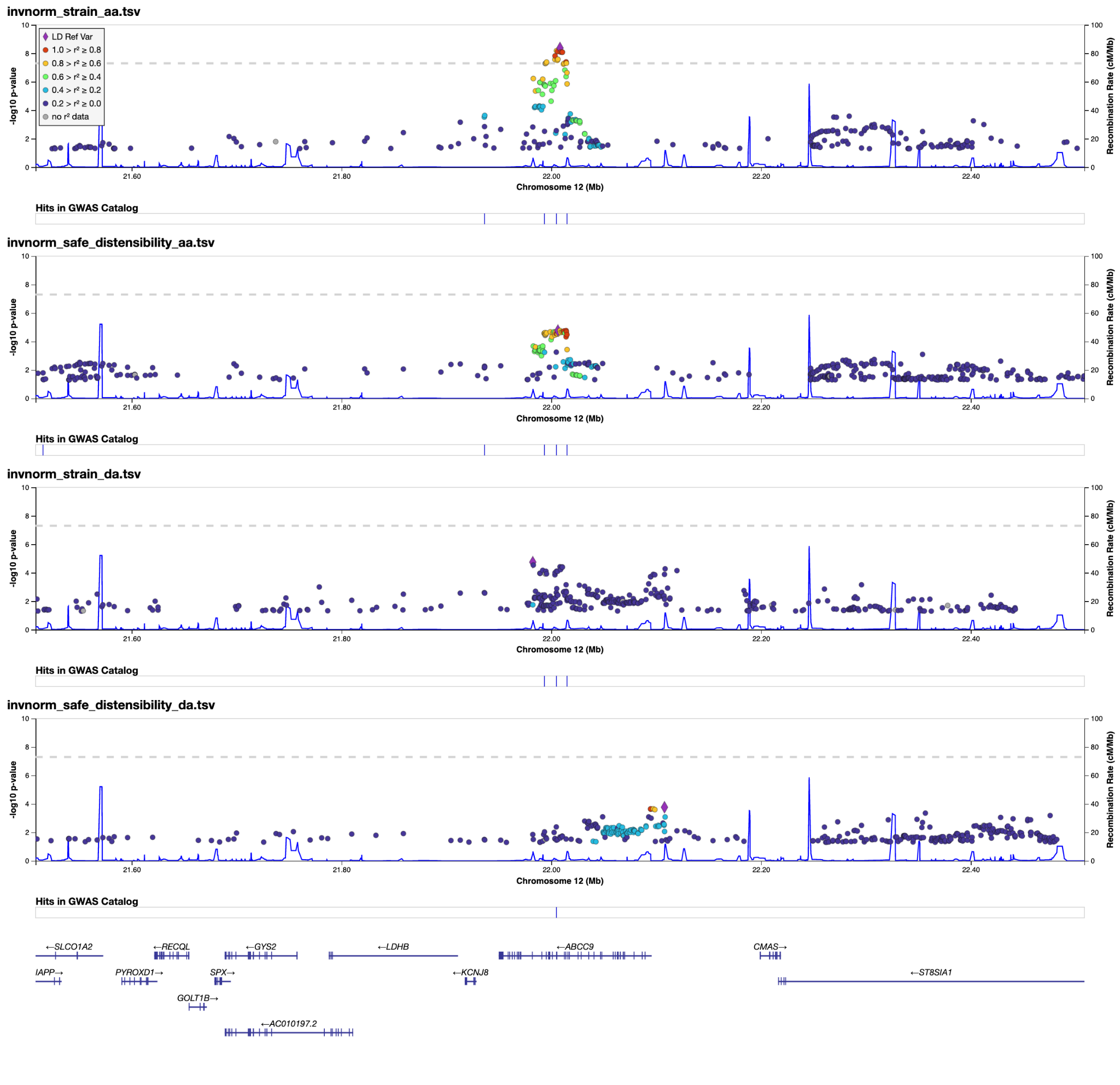

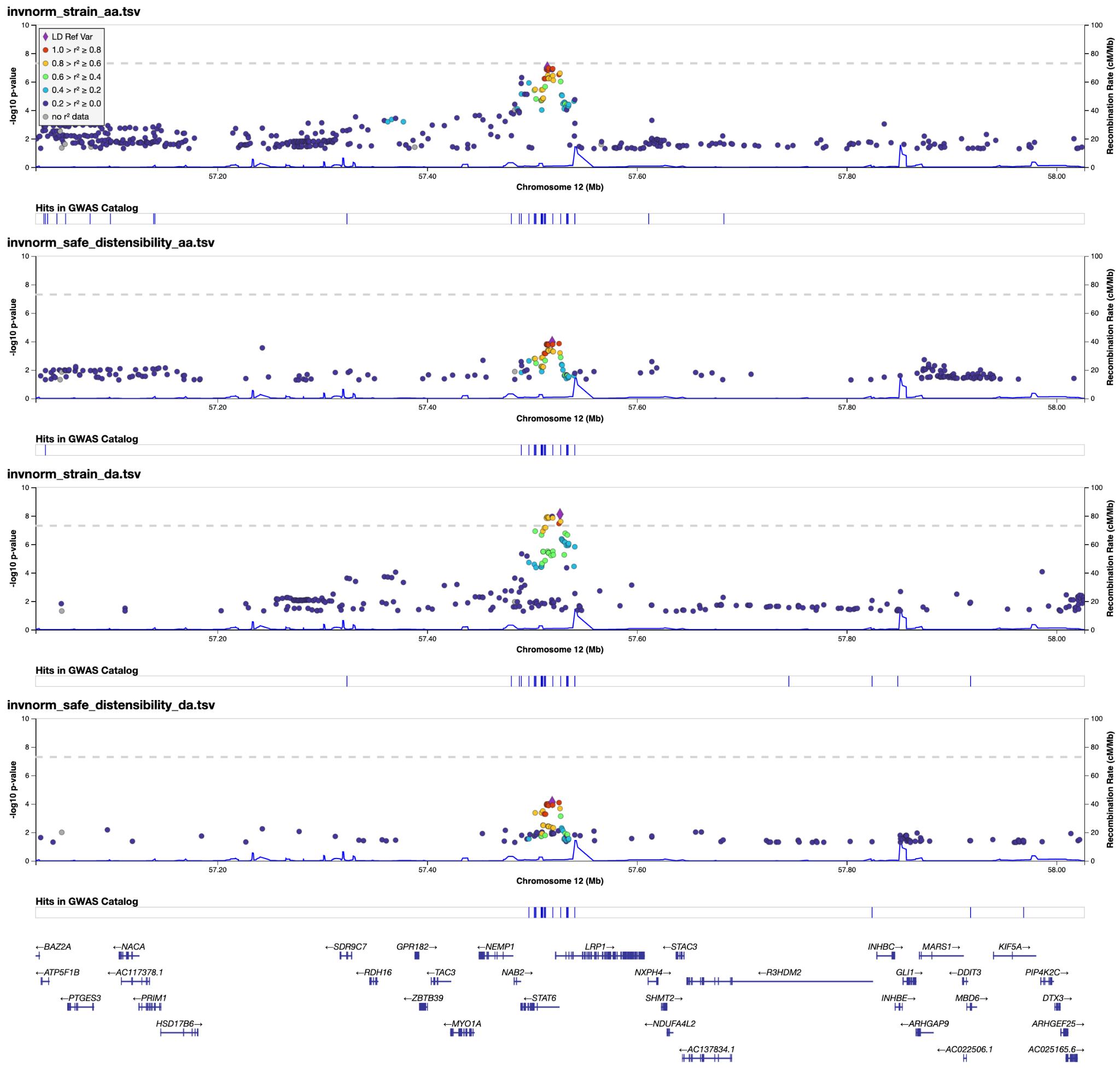

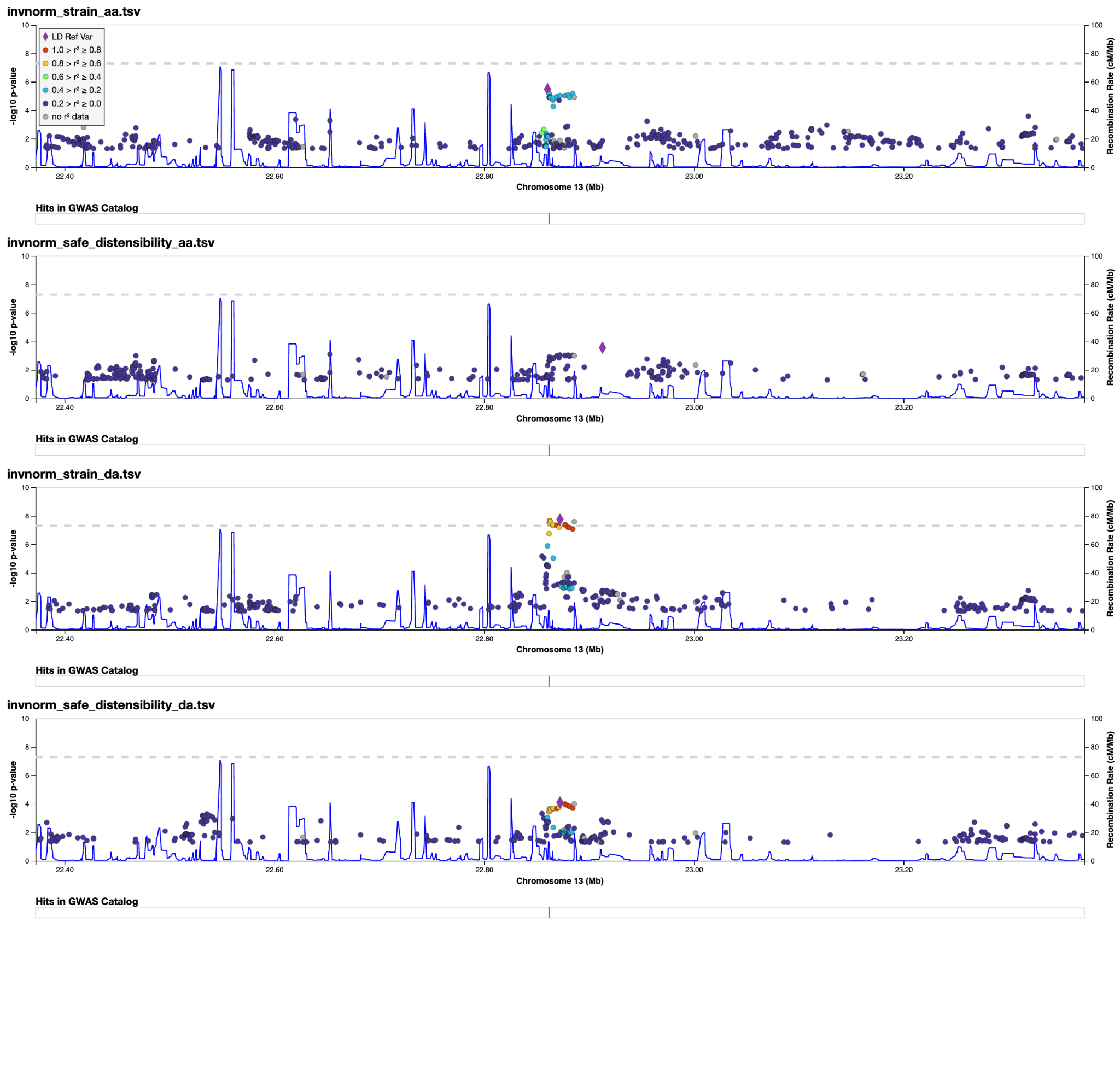

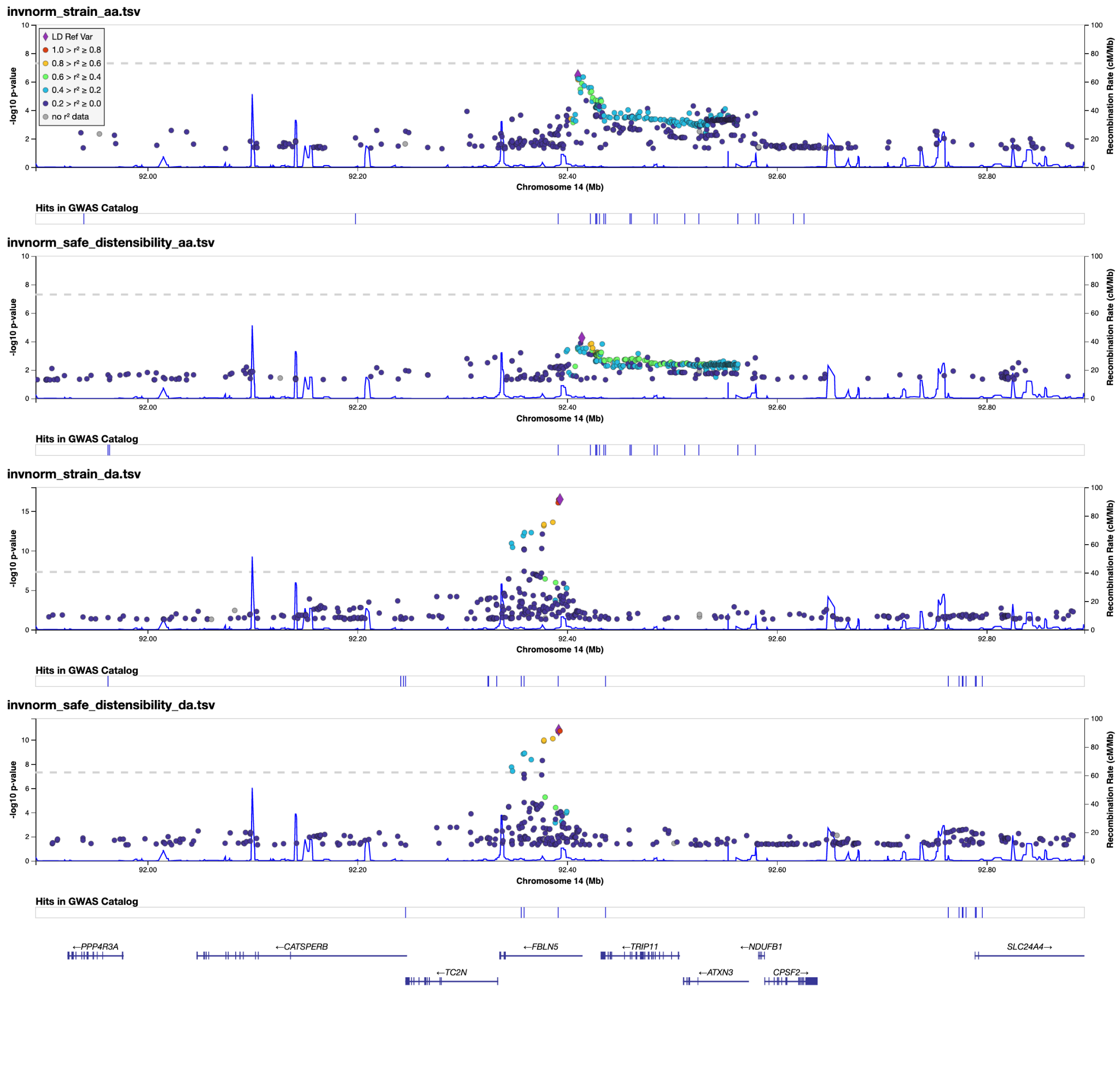

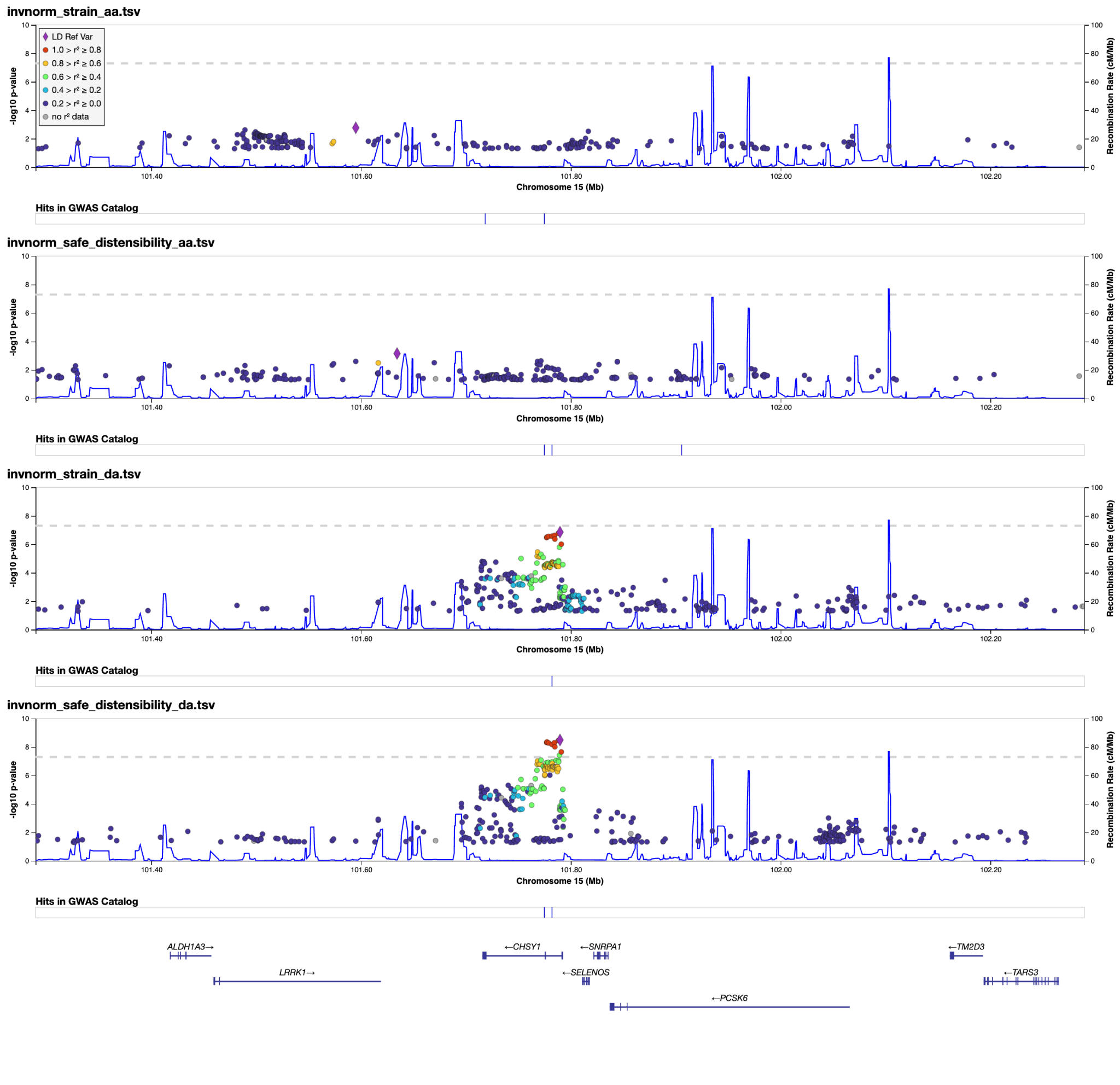

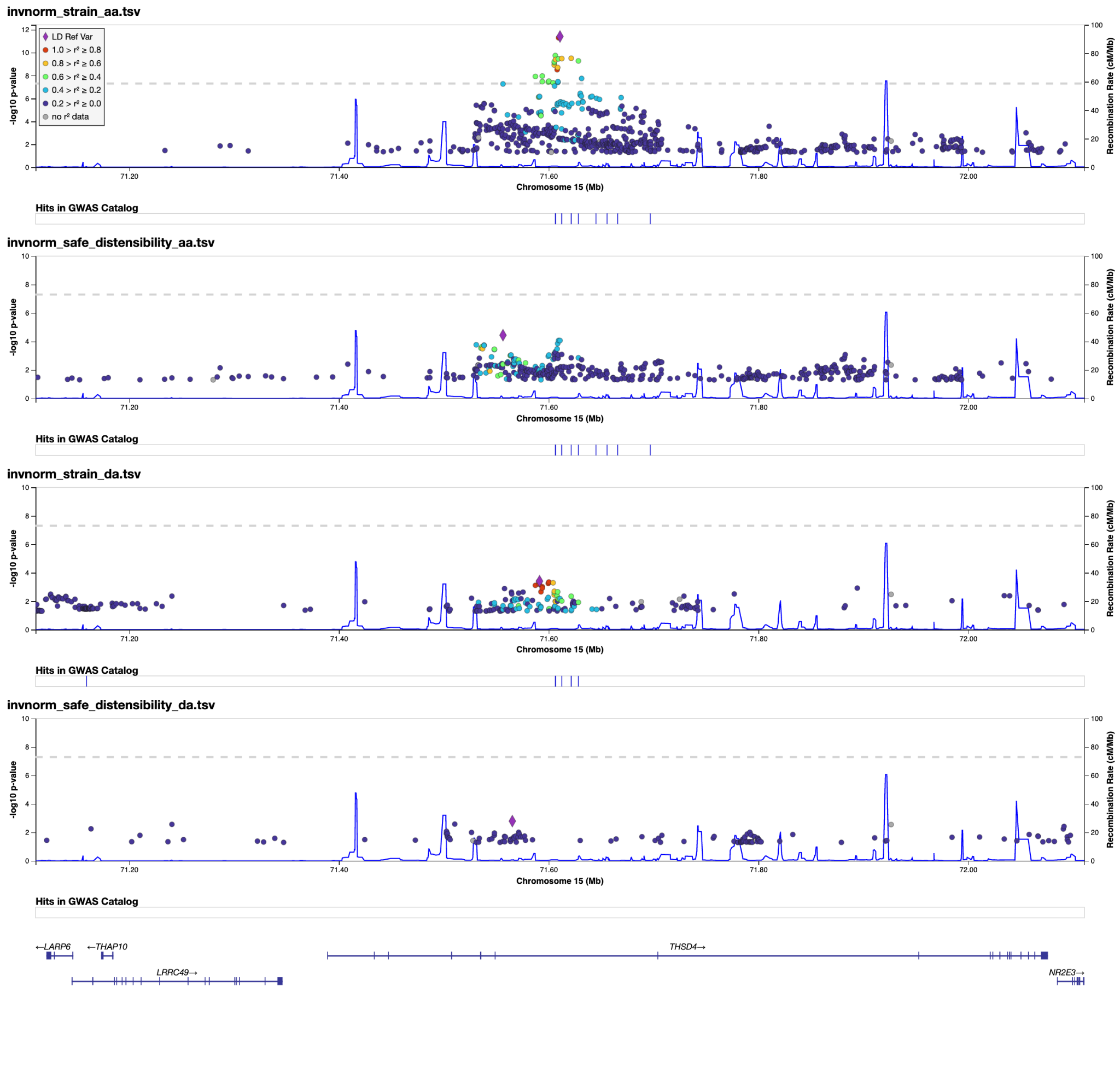

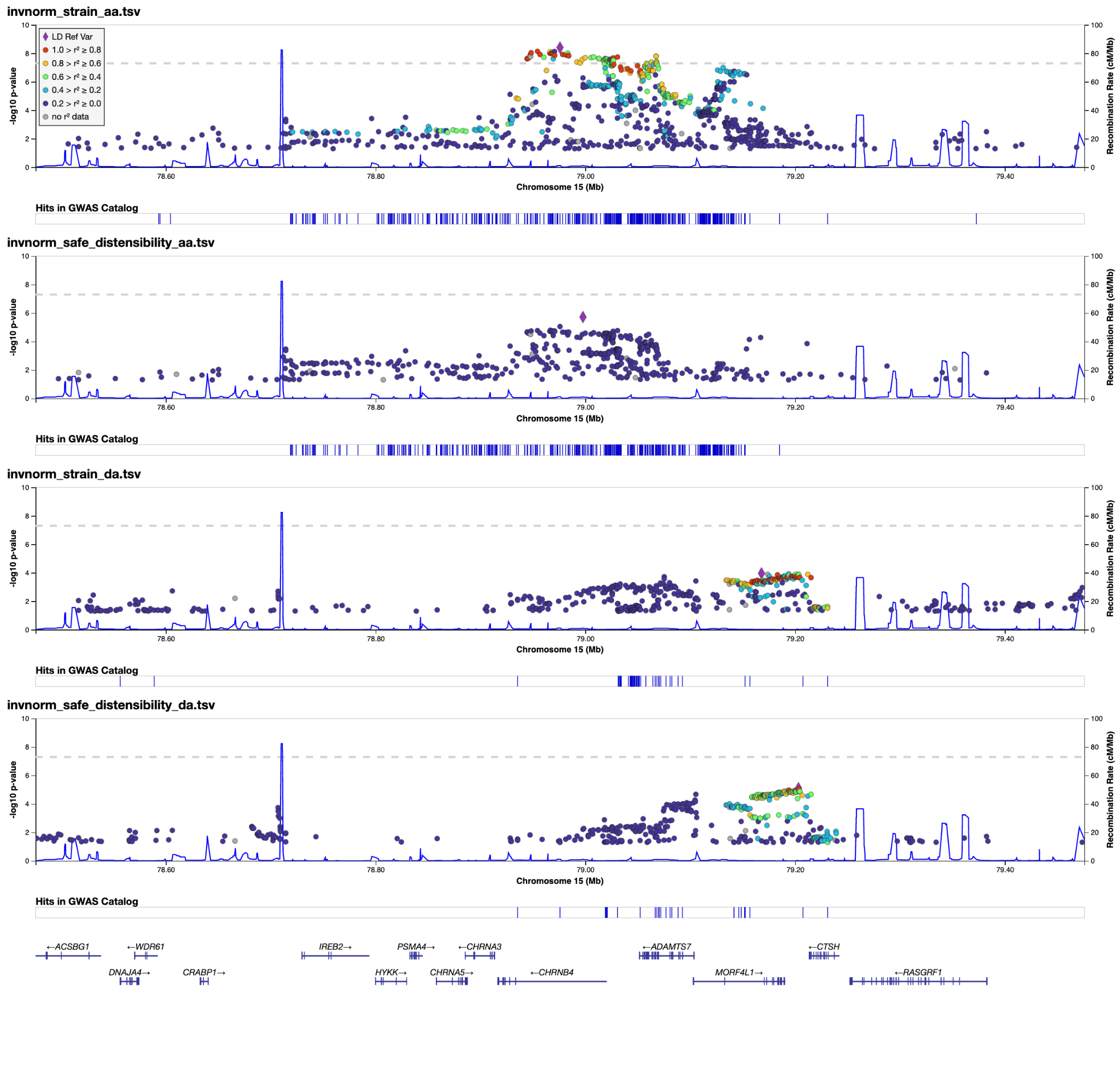

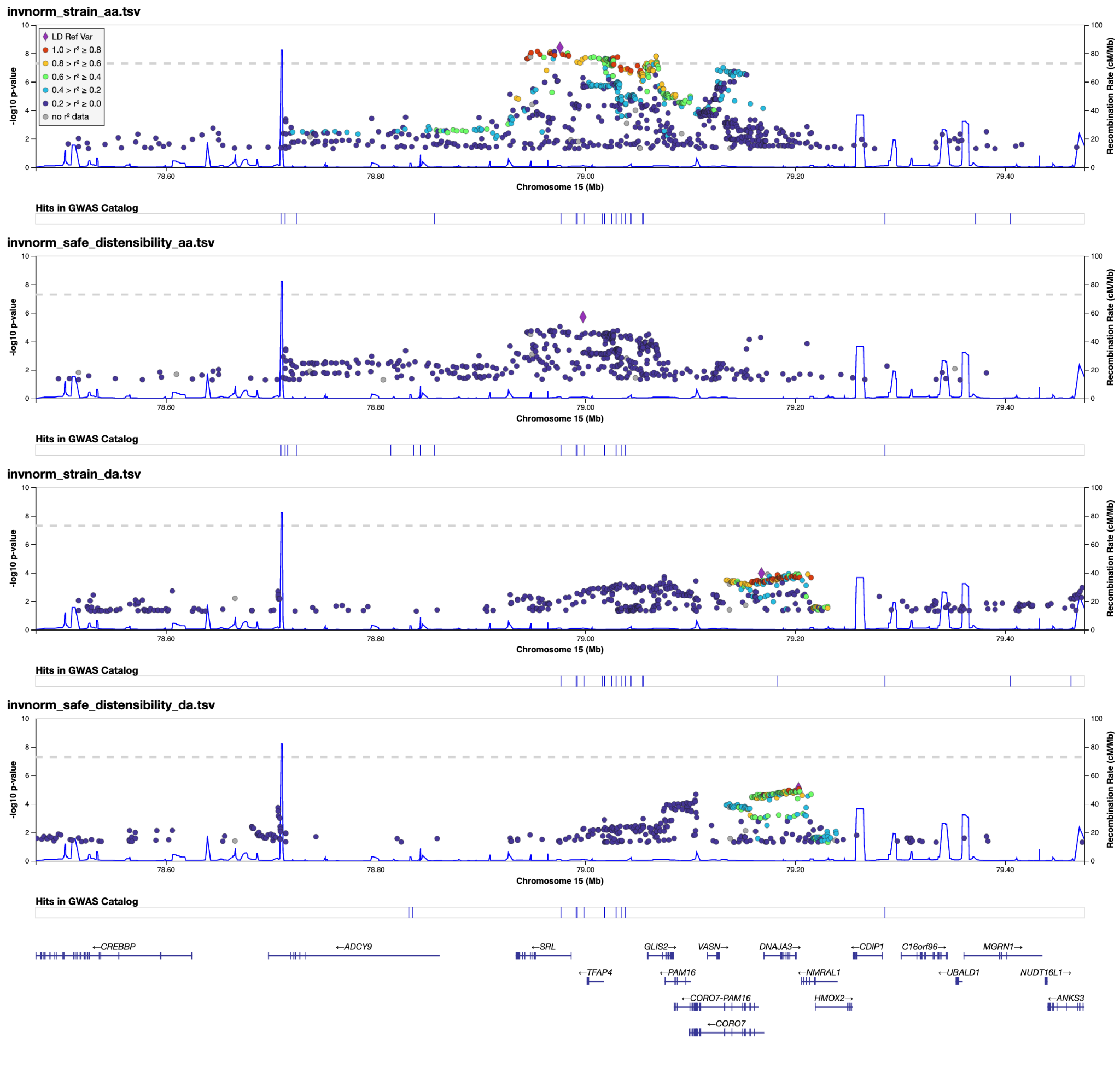

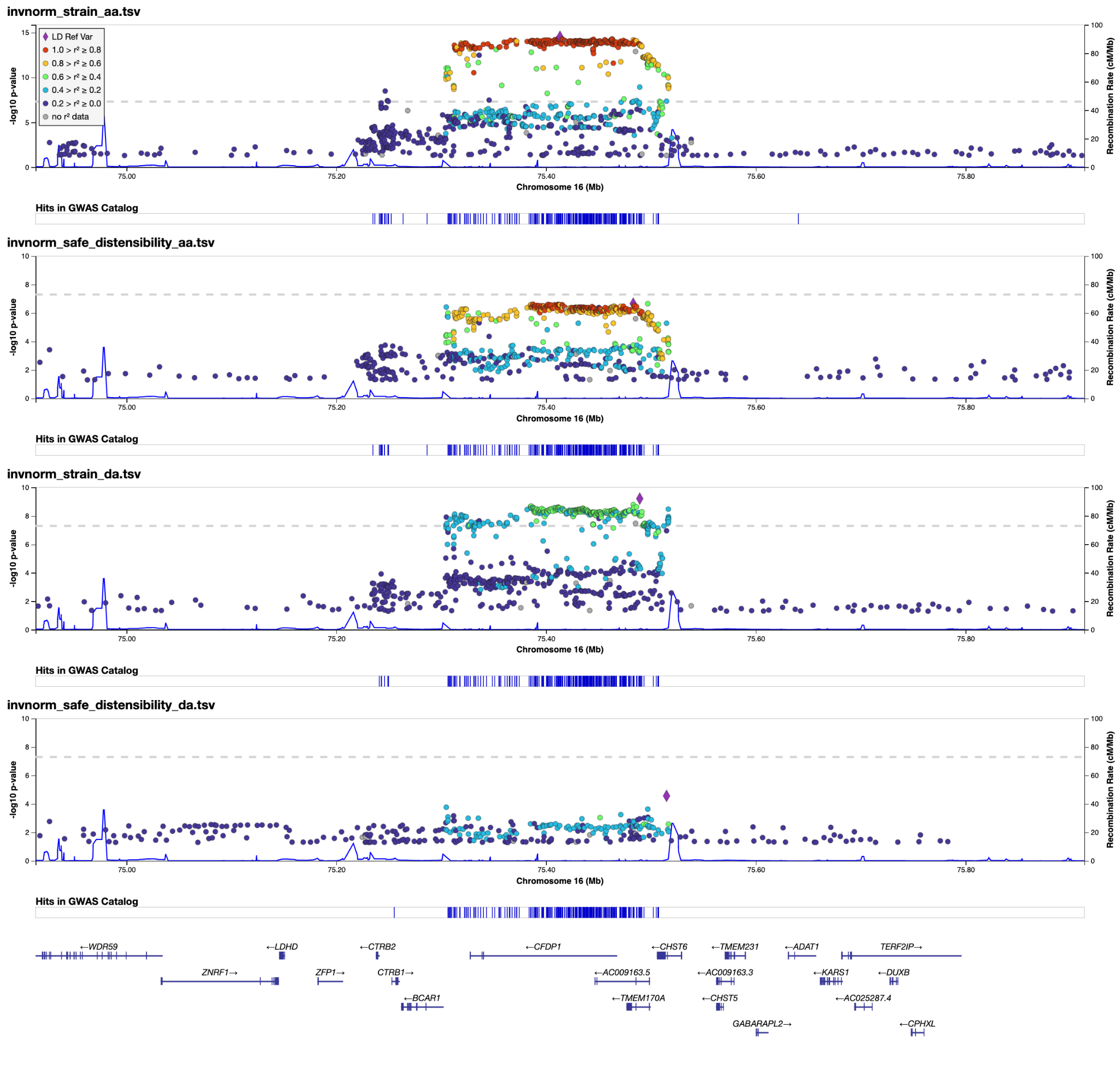

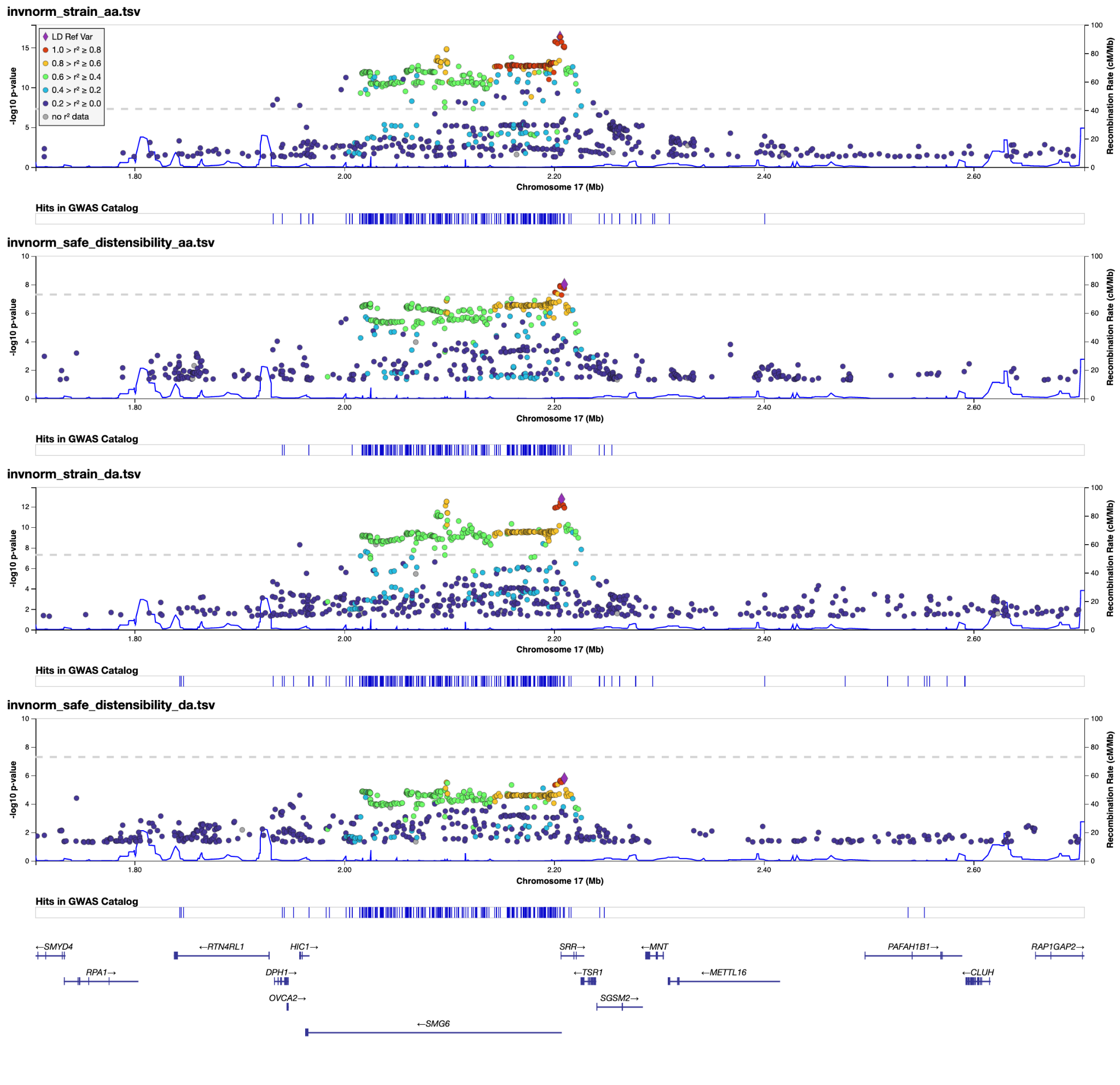

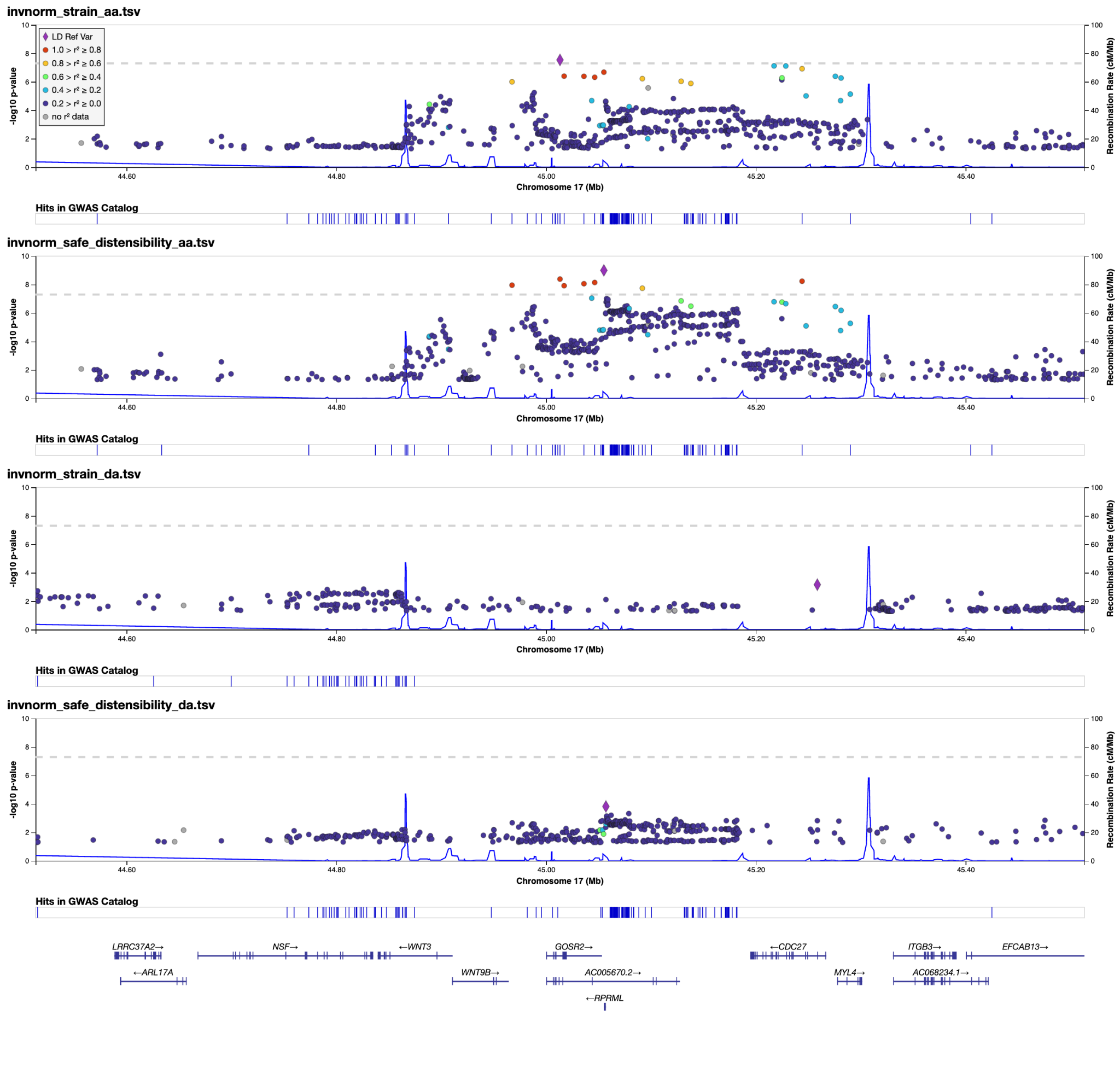

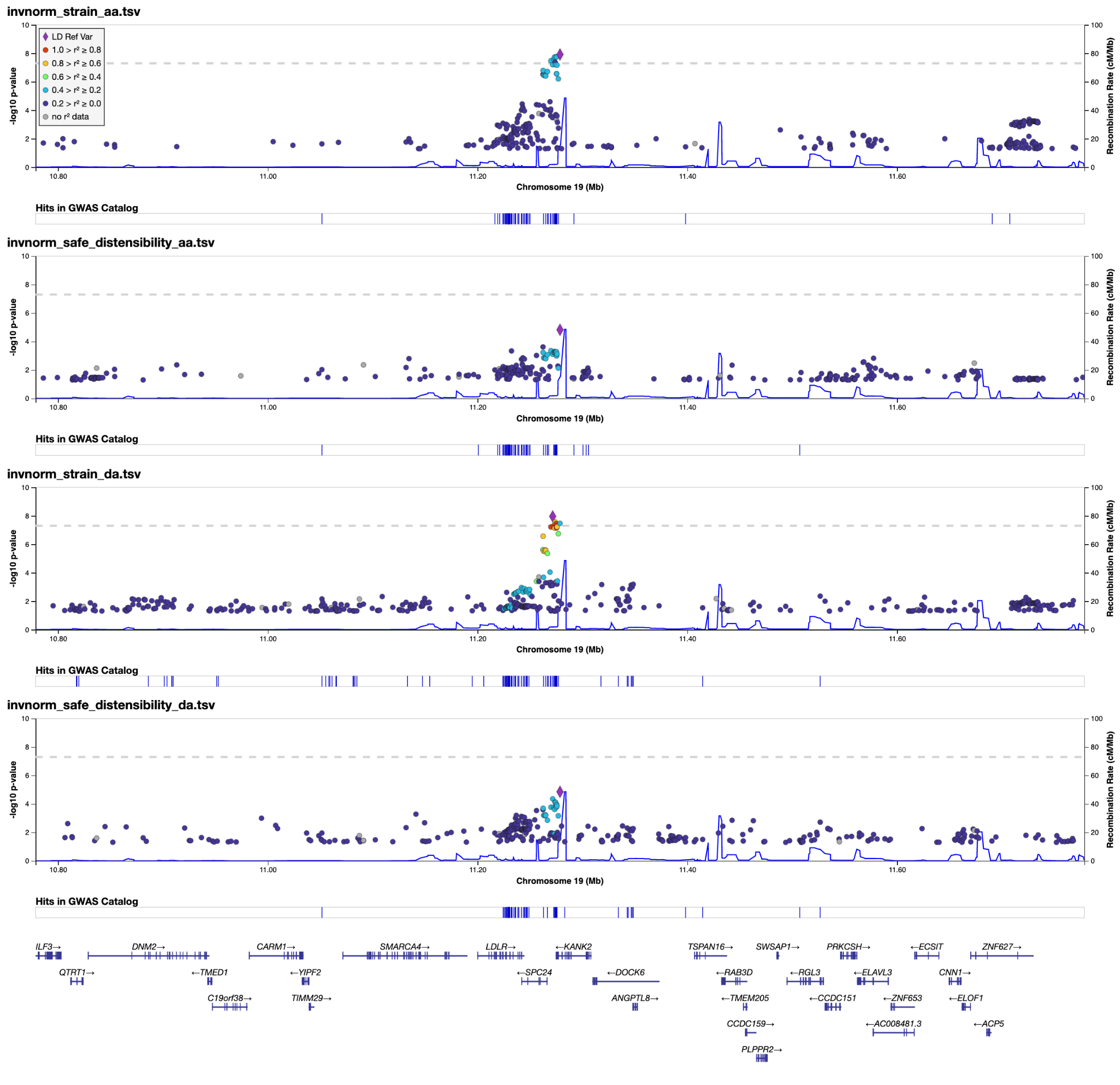

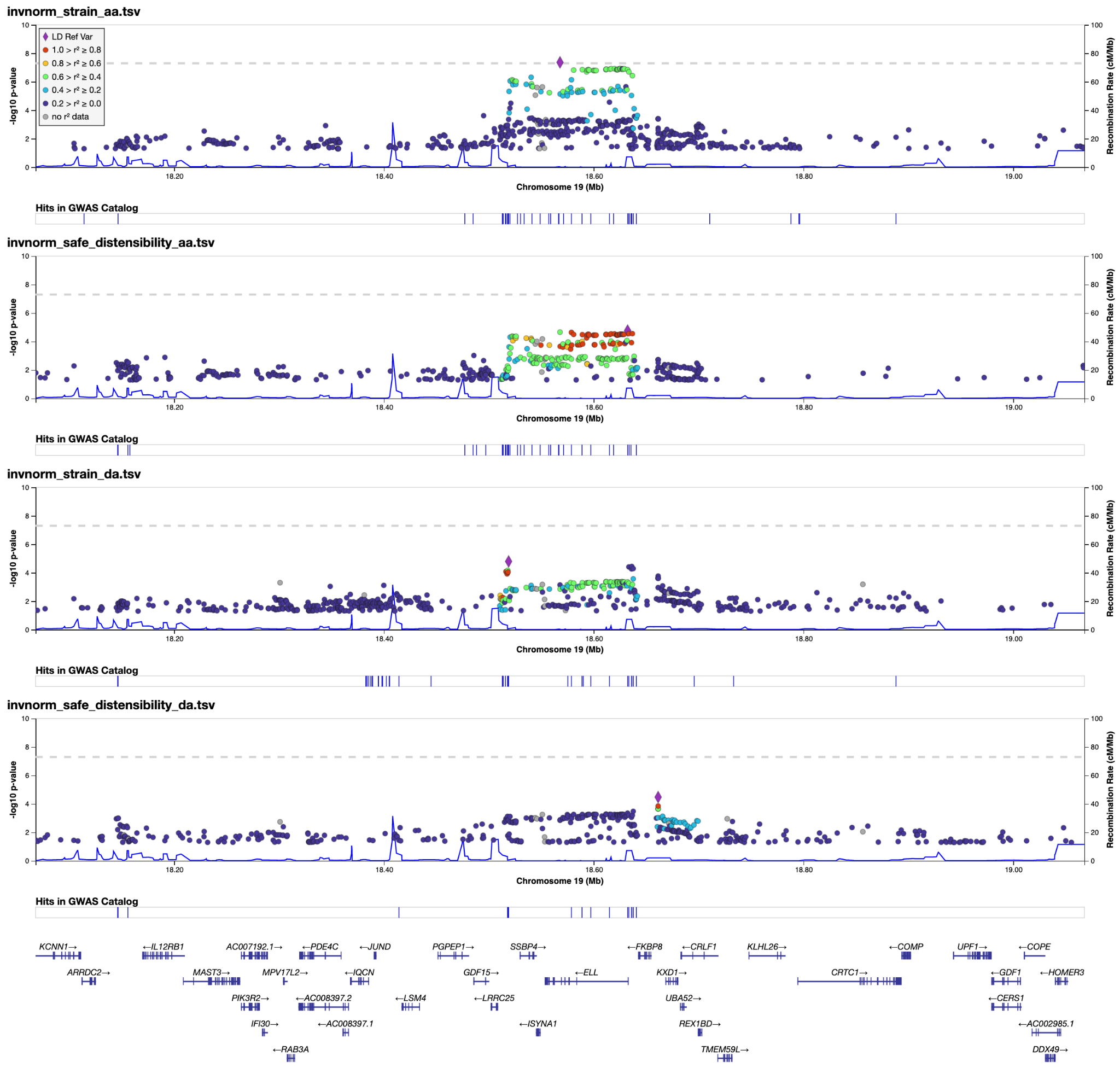

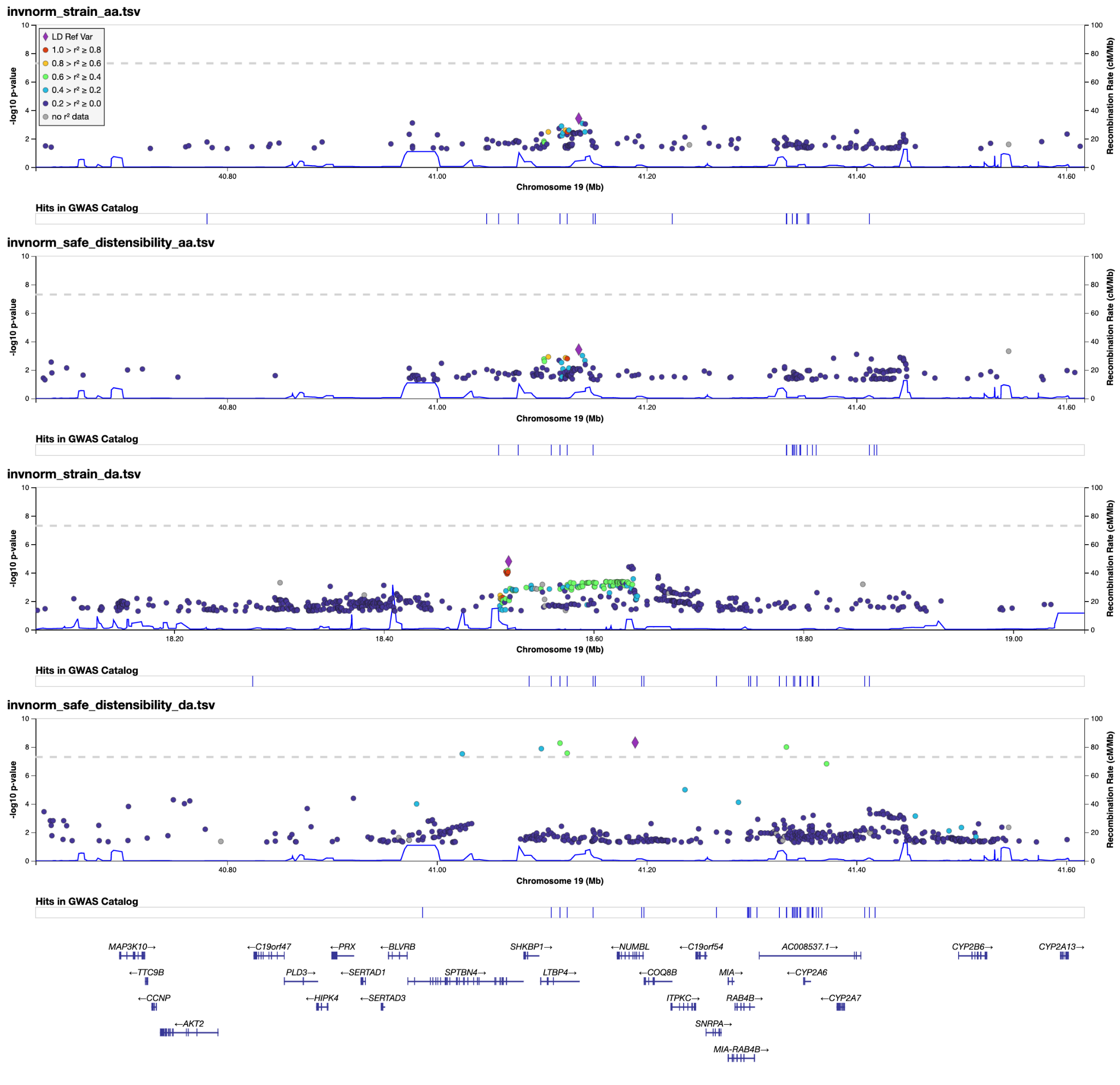

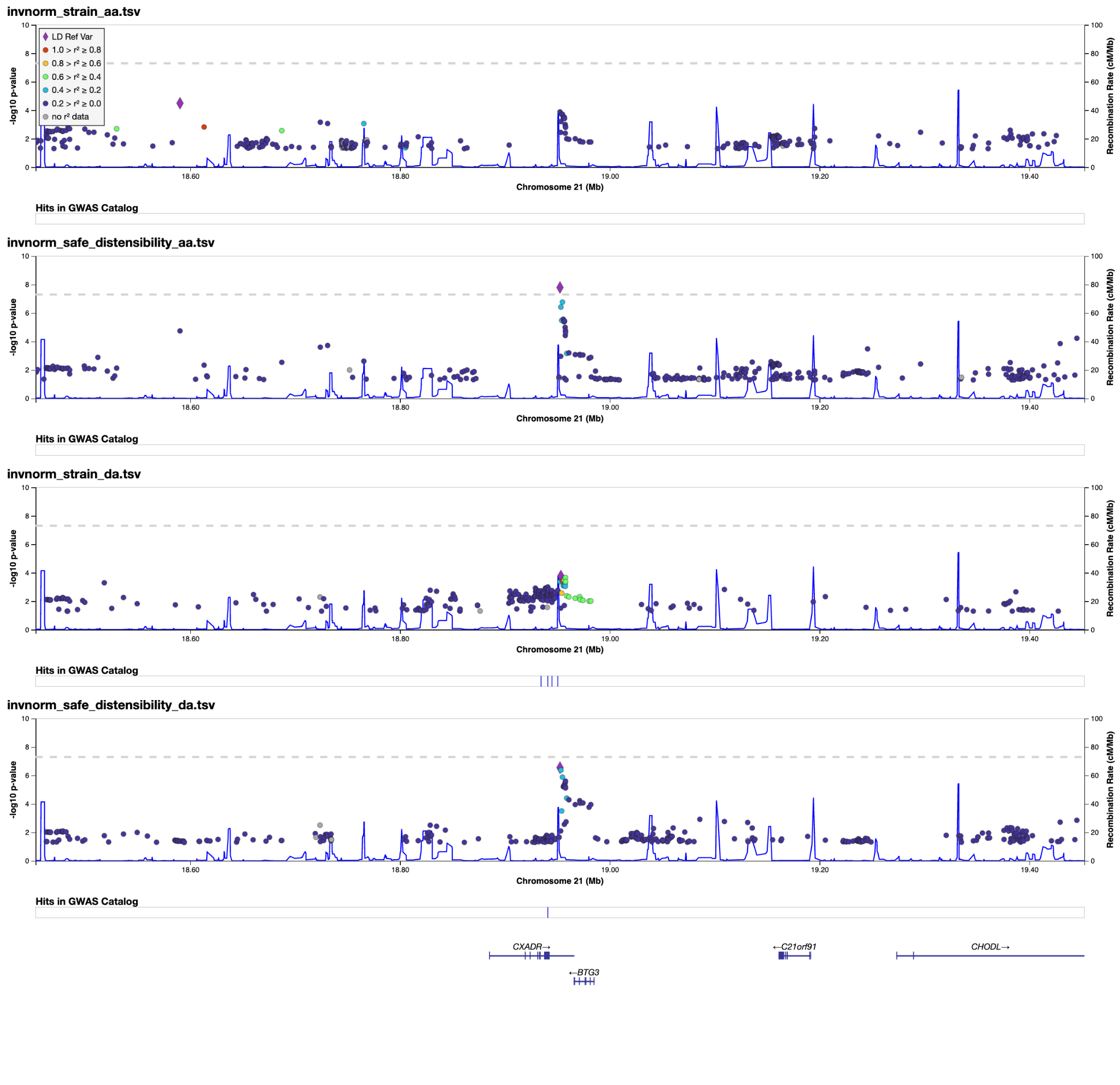

##### Supplementary Figure 10: Single nucleus sequencing

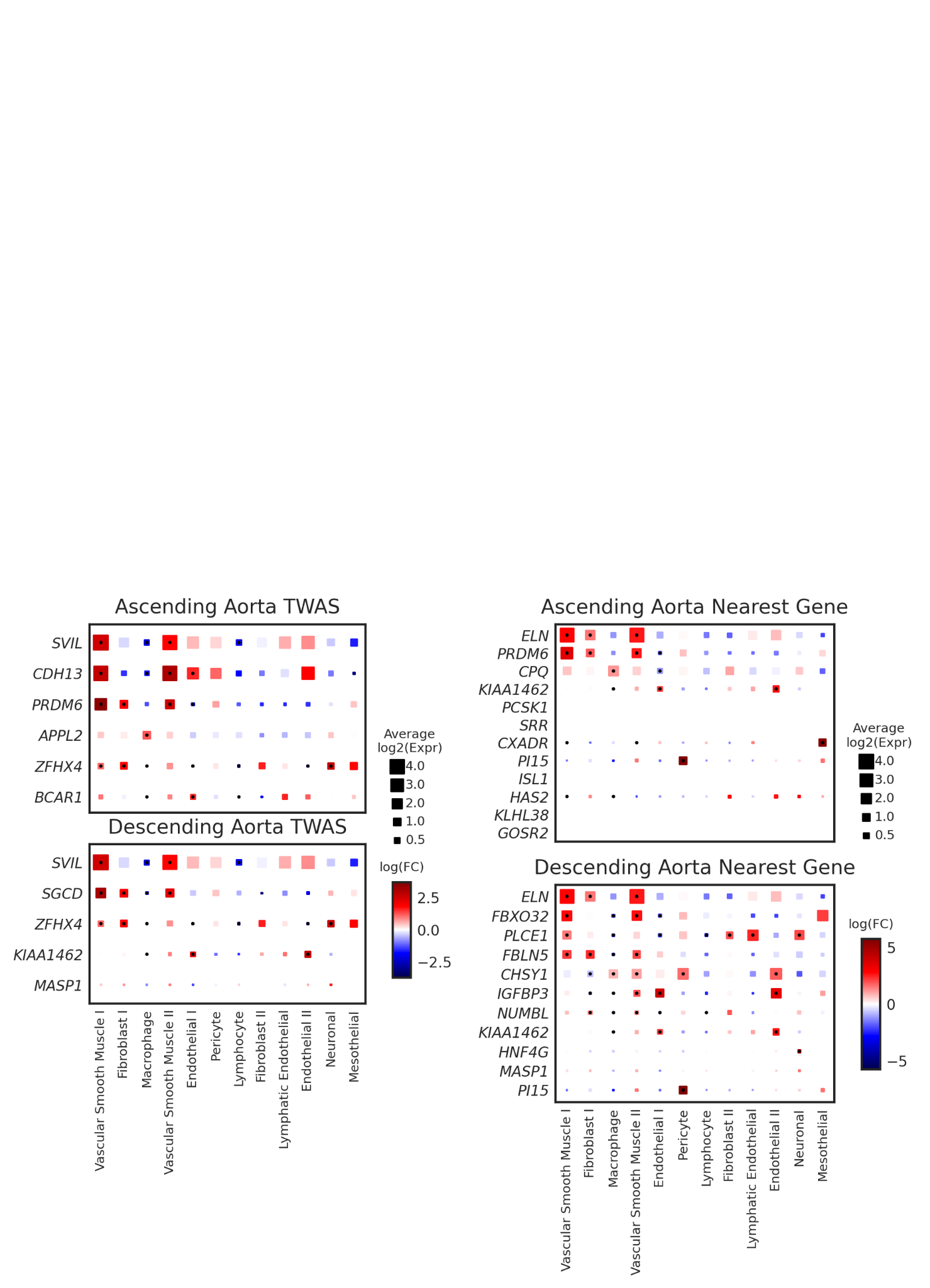

**Left**: Cell-type specificity of genes with expression data supported by the TWAS for the ascending and descending aortic distensibility. **Right**: Cell-type specificity of nearest genes to the ascending and descending aortic distensibility lead SNPs. The size of each square represents the average log2(Expr) for a gene across all nuclei in a given cluster. The color represents the log fold-change comparing the expression of the given gene in each cluster to all other clusters based on a formal differential expression model. A central black dot represents significant up- or down-regulation in the given cluster based on a Benjamini-Hochberg correction for multiple testing at FDR < 0.01. Expr = Normalized nucleus-level expression calculated as the number of counts of a gene divided by the total number of counts in the nucleus and multiplied by 10,000; FC = Fold-change.

##### Supplementary Figure 11: Rare variant association testing

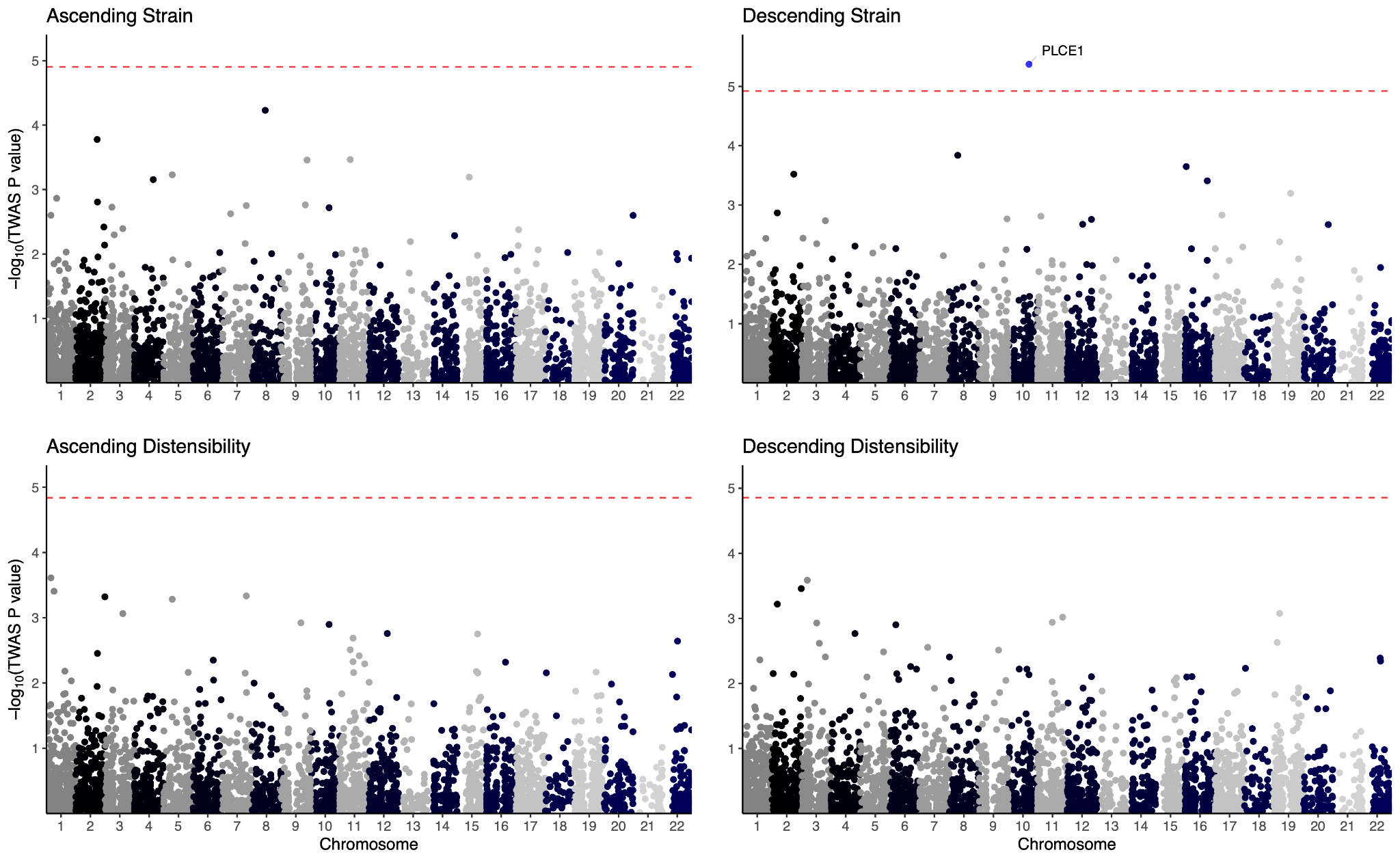

Rare variant association analysis. For each phenotype, the genes’ chromosomal position is on the **x axis** and the -log10(P value) is on the **y axis**. The horizontal dashed line indicates the per-trait Bonferroni significance.

##### Supplementary Figure 12: Polygenic scores

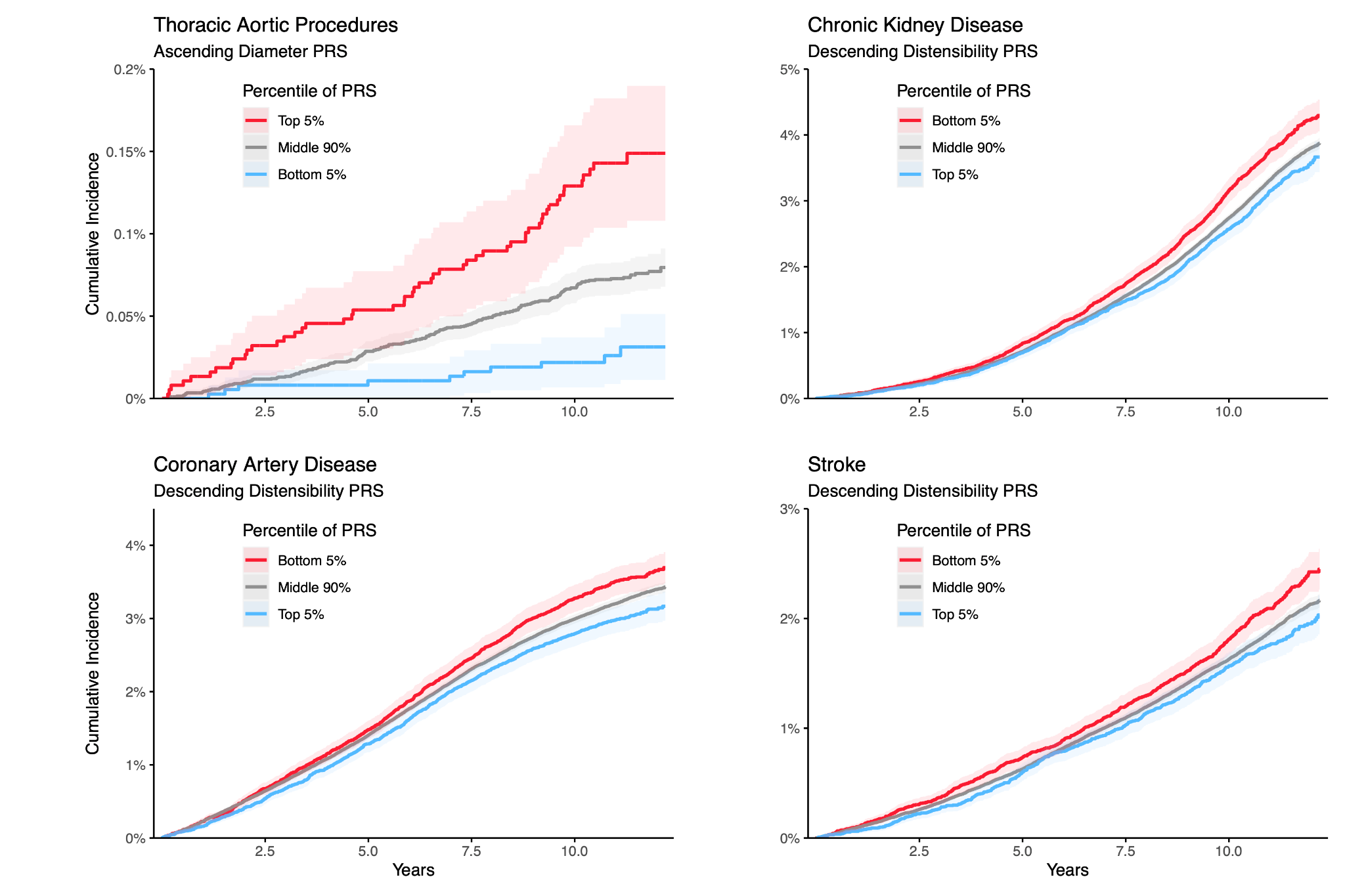

##### Supplementary Figure 13: *PLCE1* ascending aorta locus plots

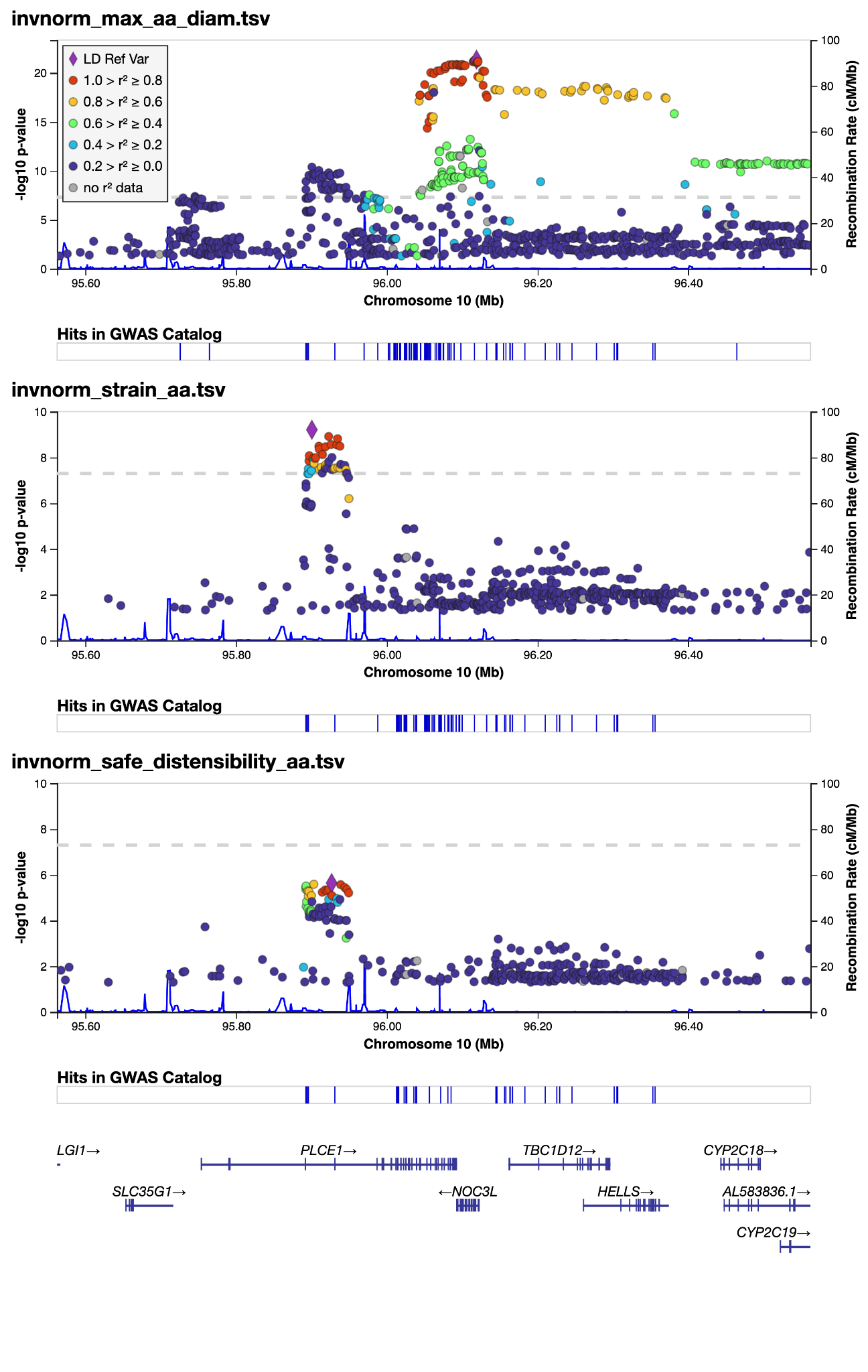

##### Supplementary Figure 14: Gene-based MAGMA for aortic diameter

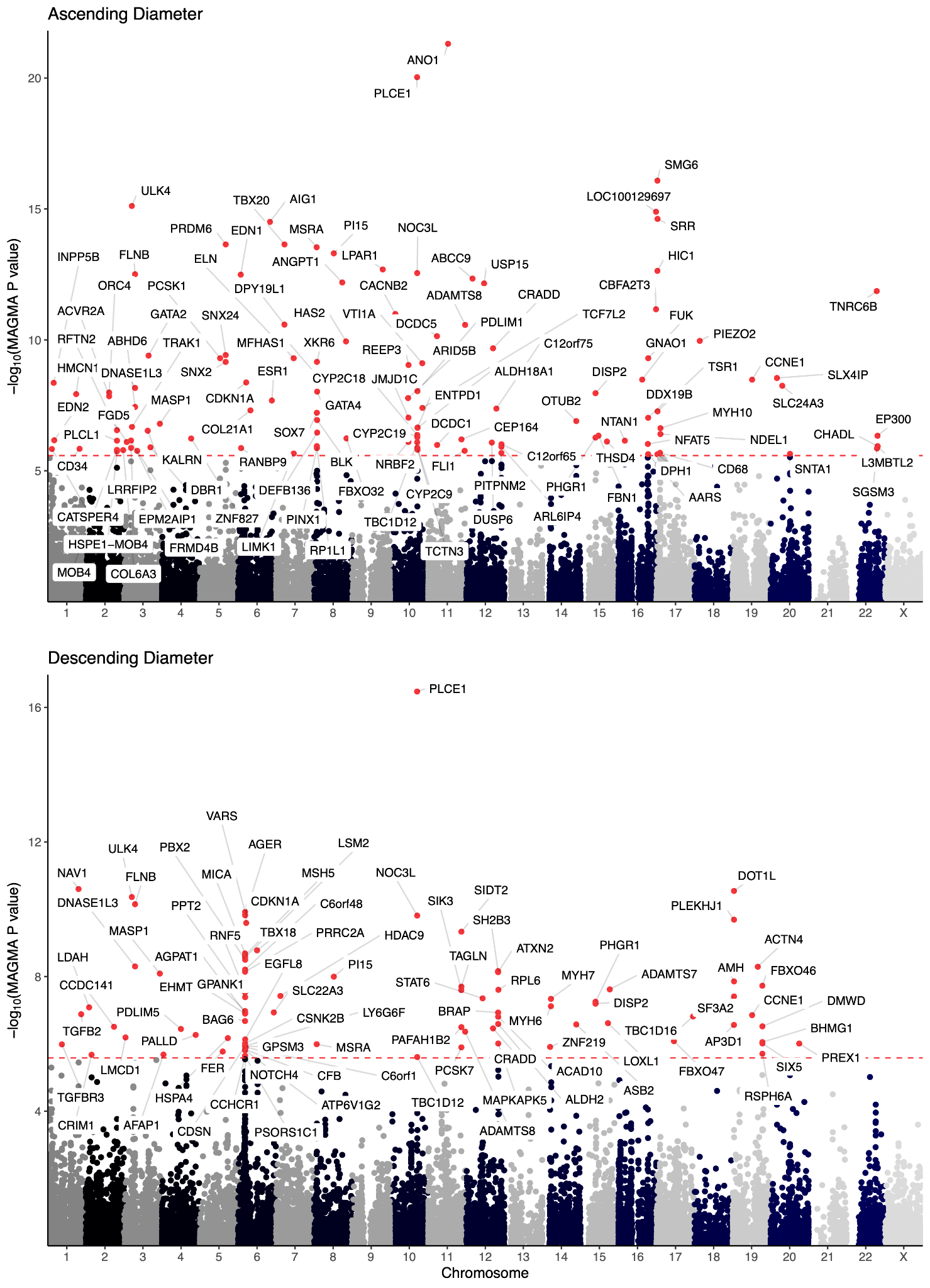

##### Supplementary Figure 15: *PLCE1* locus, common genetic variant data

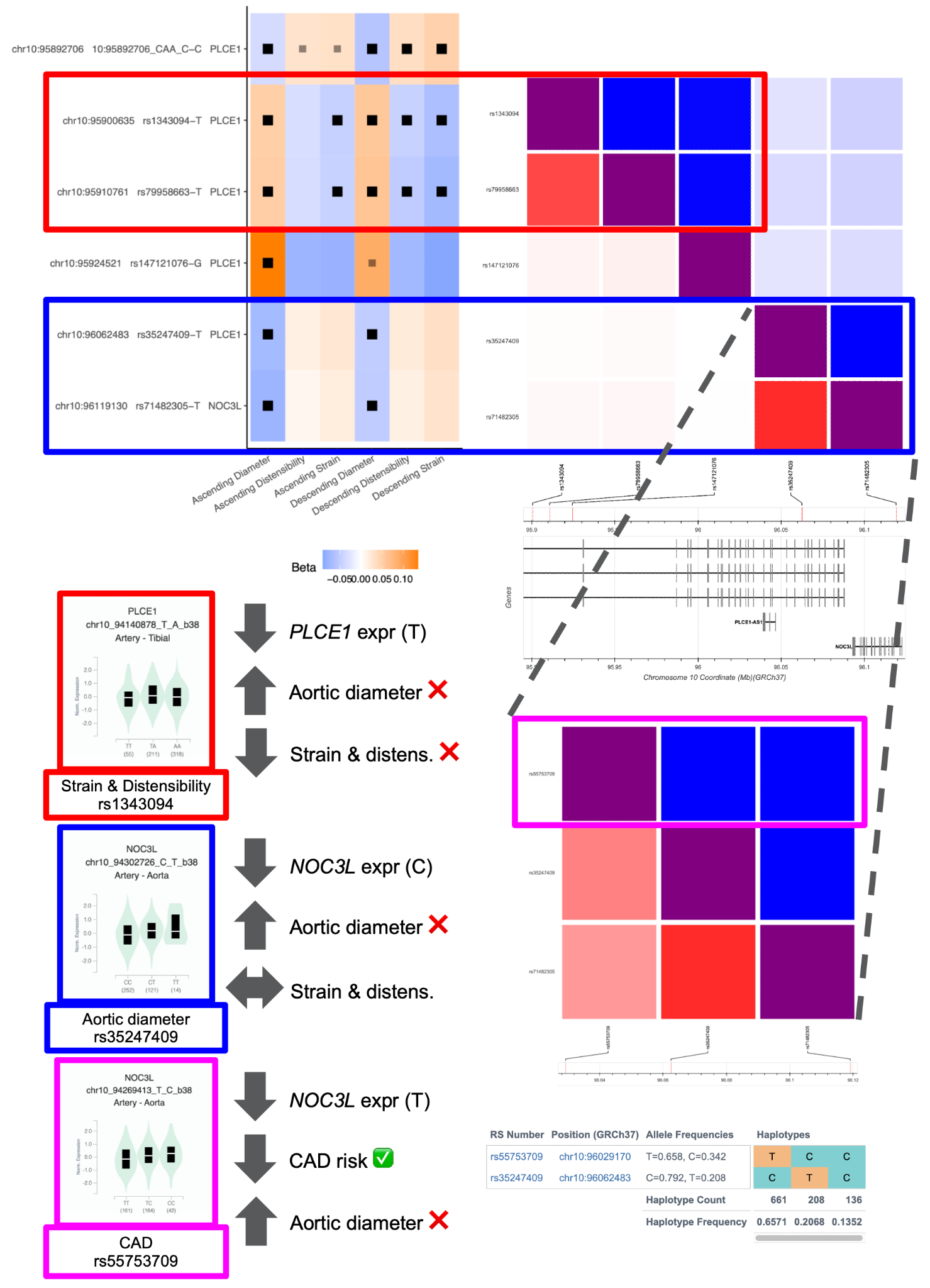

##### Supplementary Figure 16: Distensibility without QC

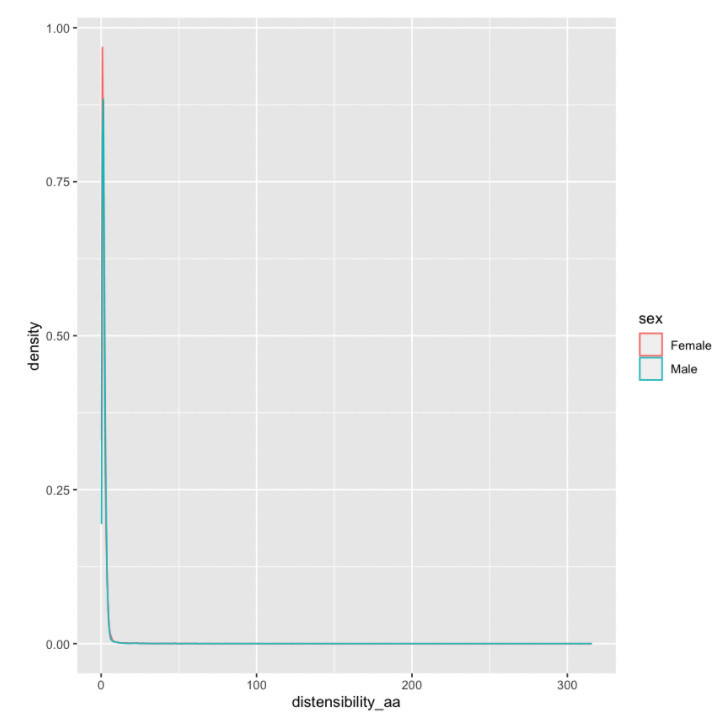

##### Supplementary Figure 17: Strain without QC

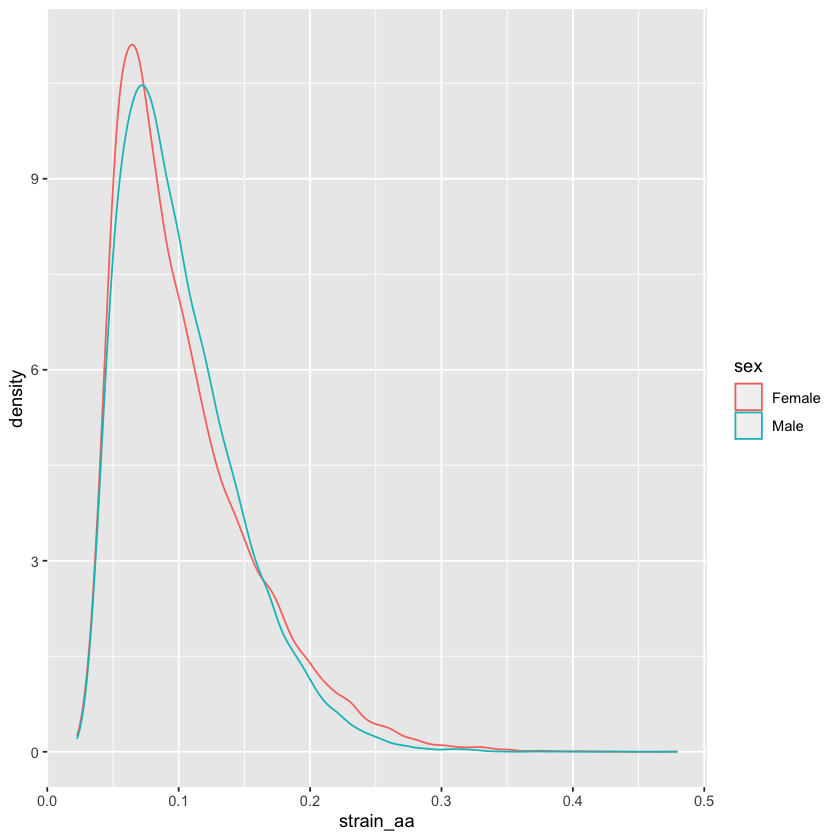

##### Supplementary Figure 18: Central pulse pressure without QC

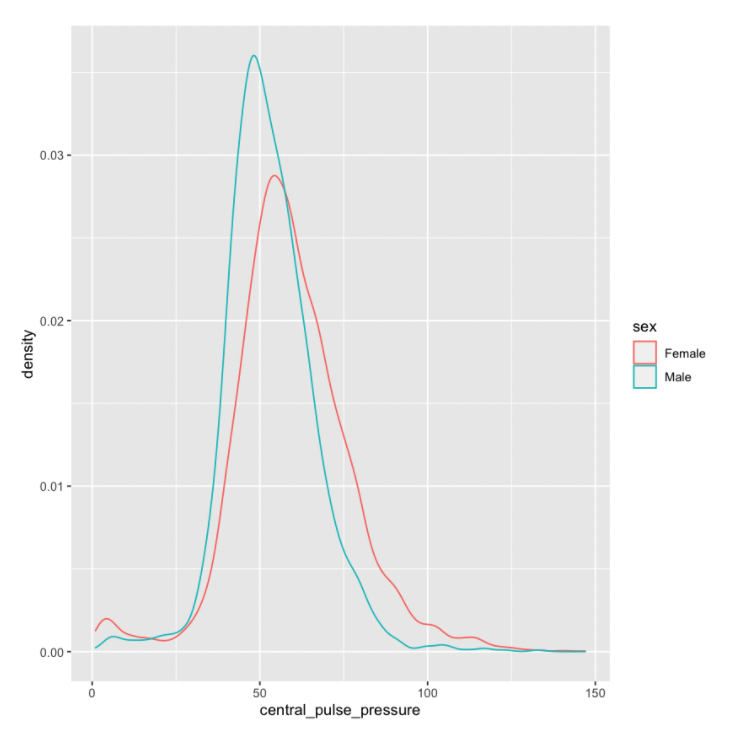

##### Supplementary Figure 19: Central pulse pressure by SBP/ESP

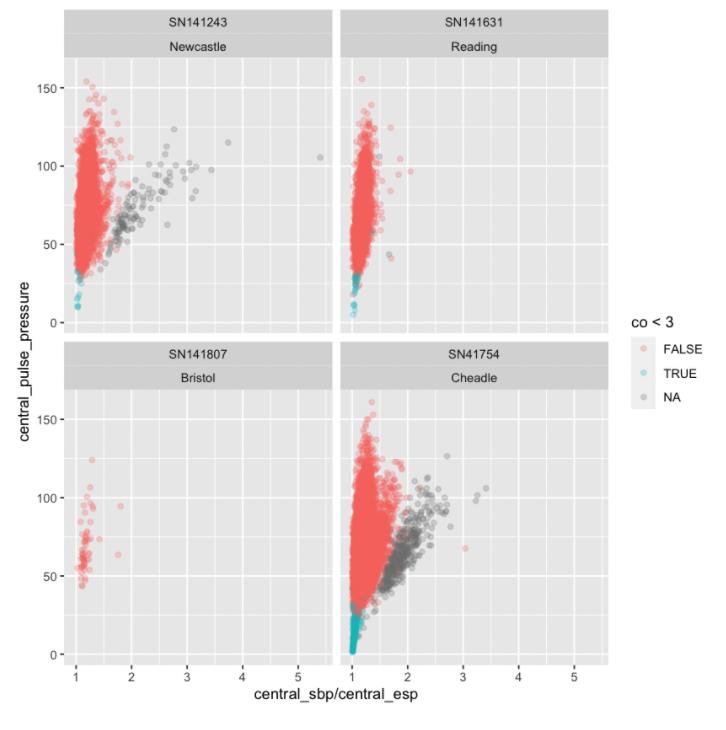

##### Supplementary Figure 20: Central pulse pressure by SBP/ESP over time

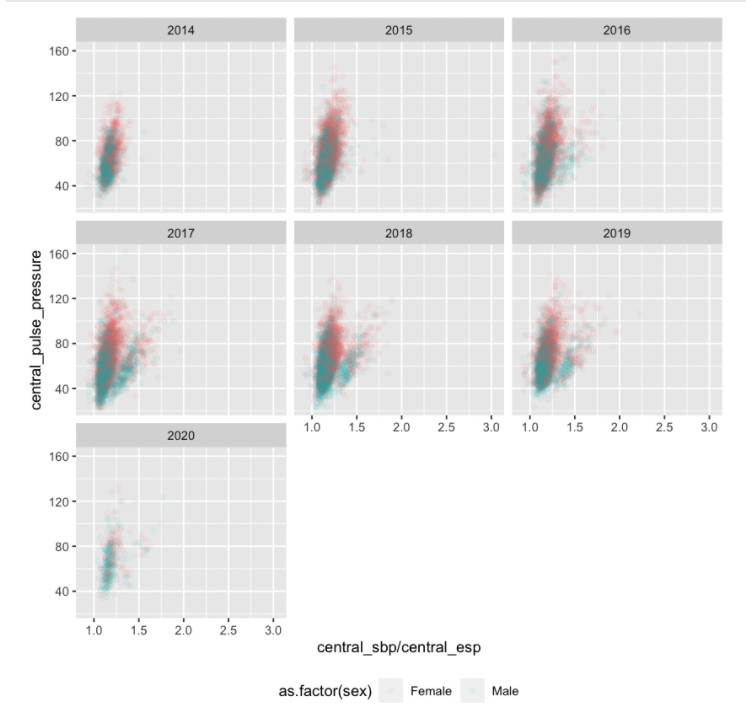

##### Supplementary Figure 21: Distensibility after QC

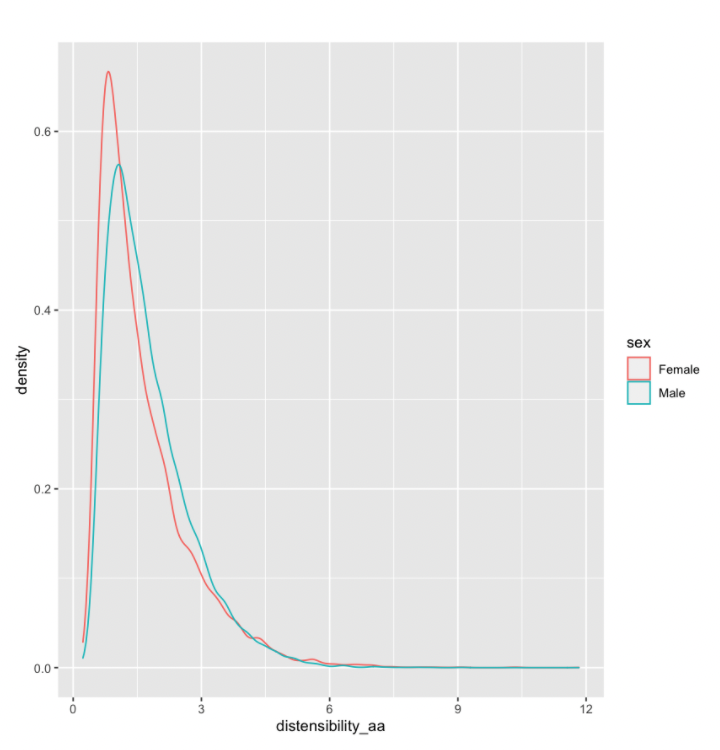
